# A third dose of inactivated vaccine augments the potency, breadth, and duration of anamnestic responses against SARS-CoV-2

**DOI:** 10.1101/2021.09.02.21261735

**Authors:** Kang Wang, Yunlong Cao, Yunjiao Zhou, Jiajing Wu, Zijing Jia, Yaling Hu, Ayijiang Yisimayi, Wangjun Fu, Lei Wang, Pan Liu, Kaiyue Fan, Ruihong Chen, Lin Wang, Jing Li, Yao Wang, Xiaoqin Ge, Qianqian Zhang, Jianbo Wu, Nan Wang, Wei Wu, Yidan Gao, Jingyun Miao, Yinan Jiang, Lili Qin, Ling Zhu, Weijin Huang, Yanjun Zhang, Huan Zhang, Baisheng Li, Qiang Gao, Xiaoliang Sunney Xie, Youchun Wang, Qiao Wang, Xiangxi Wang

## Abstract

Emergence of variants of concern (VOC) with altered antigenic structures and waning humoral immunity to SARS-CoV-2 are harbingers of a long pandemic. Administration of a third dose of an inactivated virus vaccine can boost the immune response. Here, we have dissected the immunogenic profiles of antibodies from 3-dose vaccinees, 2-dose vaccinees and convalescents. Better neutralization breadth to VOCs, expeditious recall and long-lasting humoral response bolster 3-dose vaccinees in warding off COVID-19. Analysis of 171 complex structures of SARS-CoV-2 neutralizing antibodies identified structure-activity correlates, revealing ultrapotent, VOCs-resistant and broad-spectrum antigenic patches. Construction of immunogenic and mutational heat maps revealed a direct relationship between “hot” immunogenic sites and areas with high mutation frequencies. Ongoing antibody somatic mutation, memory B cell clonal turnover and antibody composition changes in B cell repertoire driven by prolonged and repeated antigen stimulation confer development of monoclonal antibodies with enhanced neutralizing potency and breadth. Our findings rationalize the use of 3-dose immunization regimens for inactivated vaccines.

**One sentence summary:** A third booster dose of inactivated vaccine produces a highly sifted humoral immune response *via* a sustained evolution of antibodies capable of effectively neutralizing SARS-CoV-2 variants of concern.

## Main Text

The ongoing coronavirus disease 2019 (COVID-19) pandemic caused by severe acute respiratory syndrome coronavirus-2 (SARS-CoV-2) has lasted for one and a half years, resulting in an unprecedented public health crisis with over 4 million deaths globally. Progress in halting this pandemic seems slow due to the emergence of variants of concern (VOC), such as the B.1.1.7 (Alpha), B.1.351 (Beta), P.1 (Gamma, also known as B.1.1.28.1) and more recent B.1.617.2 (Delta), that appear to be high transmissible and more resistant to neutralizing antibodies (*1-4*). While several types of COVID-19 vaccines are being deployed at a large scale, new variants are thought to be responsible for re-infections, either after natural infection or after vaccination, as observed in Brazil and the United States, respectively (*5, 6*). Closely correlated with these, a general decrease in immune protection against SARS-CoV-2 variants within 6-12 months after the primary infection or vaccination is also observed (*6-8*). The prospect of genetic recombination and antigenic drift in recent SARS-CoV-2 variants together with non-uniform immune protections arising from heterogeneously waning humoral immunity in COVID-19 convalescent or vaccinated individuals, point to the potential risks of a long-term pandemic that could endanger the global human health, diminishing social, economic and outdoor leisure activities. A plausible approach to solving this problem is the administration of a third dose of the vaccine somewhere between 6 and 12 months after the 2nd dose of vaccination for enhancing and prolonging the protection. However, not much is known about the immunogenic features of such a booster dose of a COVID-19 vaccine. In addition, there are large gaps in our understanding about correlating immunogenic findings from surrogate endpoints to gauge vaccine efficacy.

The CoronaVac, a 2-dose β-propiolactone-inactivated vaccine against COVID-19, has been approved for emergency use by the World Health Organization (*9, 10*). In human clinical trials (phase I/II, registration number: NCT04352608), a subgroup with a 3-dose immunization schedule at months 0, 1, 7 was also included. To evaluate immune features, we recruited 22 COVID-19 convalescents, 6 healthy participants (SARS-CoV-2 negative, confirmed by RT-PCR) and 38 volunteers who received either 2 or 3 doses of the Coronavac vaccine for blood donation. The volunteers ranged from 16 to 69 years old (median 33); 30 (45.5%) were men and 36 (54.5%) were women. None of the volunteers recruited for vaccination was infected by SARS-CoV-2 prior to the study. Blood samples from convalescents and vaccinees collected 1.3 months after infection and the indicated times after vaccination were used in this study, respectively, to compare humoral immune responses elicited against circulating SARS-CoV-2 variants.

Neutralizing antibodies (NAbs) are a major correlate of protection for many viruses, including SARS-CoV-2, and have also provided the best correlate of vaccine efficacy. Several types of SARS-CoV-2 neutralization assays have been described using either live SARS-CoV-2 or a pseudo-typed reporter virus carrying SARS-CoV-2 spike protein (S). Both types of assays could yield reproducible neutralizing titers, with the pseudo-typed virus neutralization assay exhibiting higher sensitivity (*11, 12*). Neutralizing activity of plasma samples from 66 participants was measured against WT, B.1.351, P.1 and B.1.617.2 using live SARS-CoV-2 and VSV-pseudoviruses with the S from WT, B.1.1.7, P.1 variants and SARS-CoV (Fig. 1). The geometric mean half-maximal neutralizing titers (GMT NT_50_) against live SARS-CoV-2 in plasma obtained from convalescents and from vaccinees (4 weeks after the final vaccination) suggest an approximately 60% higher neutralizing activity against WT after 3-dose inoculation when compared with 2-dose administration, and 20% higher than those from convalescents (Fig. 1A). Interestingly, for the samples from the convalescents, 2-dose and 3-dose vaccinees, neutralizing titers against B.1.351 were, on average, 7.7-fold, 5.7-fold and 3.0-fold reduced, respectively, compared with WT (Fig. 1A). Similarly, fold decreases in neutralization ID_50_ titers against P.1 and B.1.617.2 for the three cohorts were 5.3, 4.3 and 3.1, and 5.3, 3.7 and 2.3, respectively (Fig. 1A). Overall, plasma of the 3-dose vaccinees displayed minimal reduction in neutralization titers against several authentic VOCs compared to the convalescents and 2-dose vaccinees (Fig. 1A). Remarkably, ∼41% (9/22) and 50% (6/12) samples from the convalescents and 2-dose vaccinees, respectively, failed to reach 50% neutralization at a plasma dilution of 1: 10, with ∼14% (3/22) and 16% (2/12) showing a near ineffectiveness in neutralizing B.1.351 *in vitro* (Fig. 1A). By contrast, only 1 out of 14 samples from the 3-dose vaccinees exhibited a weak neutralizing titer below 10 (Fig. 1A). Importantly, the 3-dose vaccinees showed over 2.5-fold higher neutralizing potency against B.1.617.2 than the convalescents and 2-dose vaccinees (Fig. 1A). The GMT NT_50_ values measured by a VSV-pseudovirus with the WT S were 840, 660 and 1,176 for convalescents, 2-dose and 3-dose vaccinees, respectively, which were 8-10-fold greater than those determined by live WT SARS-CoV-2 (Fig. 1A, 1B), confirming higher sensitivity of pseudovirus-based assays in determining neutralizing titers. In line with the results of live SARS-CoV-2 neutralization assay, the mean fold decrease in the neutralization of B.1.1.7 relative to the WT was 2.8-fold for convalescents, 2.2-fold for 2-dose vaccinees and 1.7-fold for 3-dose vaccinees (Fig. 1B). Similarly, plasma from convalescents, 2-dose and 3-dose vaccinees exhibited a 4.5-fold, 2.9-fold and 2.4-fold reduction, in NAb titers against P.1, respectively, when compared to the WT (Fig. 1B). These results reveal that a third-dose boost of inactivated vaccine leads to enhanced neutralizing breadth to SARS-CoV-2 variants, bolstering the potential to ward off VOCs effectively when compared to convalescent plasma. Of note, neither vaccination nor SARS-CoV-2 infection boosts distinct neutralizing potency against SARS-CoV, presumably due to the relatively far phylogenic relationship (Fig. 1B).

**Fig. 1.**
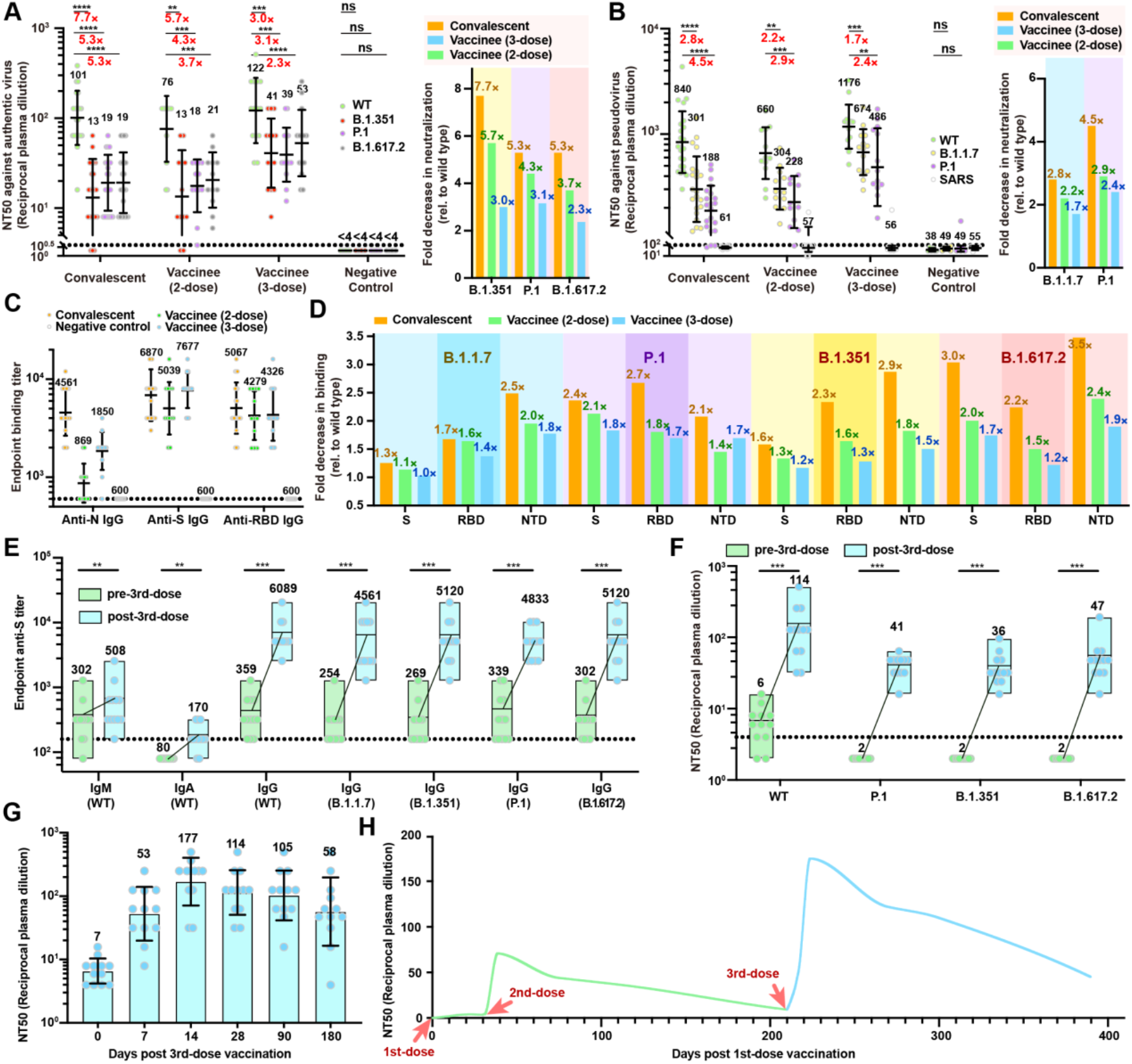
A 3^rd^-dose booster of an inactivated vaccine elicits an expeditious and long-lasting recall antibody response. Plasma neutralizing activity evaluated by authentic SARS-CoV-2 **(A)** and pseudo-typed SARS-CoV-2 neutralization assays **(B)** Left: half-maximal neutralizing titer (NT_50_) values for plasma from COVID-19 convalescents, 2-dose, 3-dose CoronaVac vaccine recipients (at week 4 after the last dose of vaccination) and negative controls (pre-COVID-19 historical control) against live SARS-CoV-2 WT, B.1.351, P.1 and B.1.617.2, and VSV-based SARS-CoV-2 pseudoviruses bearing WT or B.1.1.7 or P.1 S protein. Black bars and indicated values represent geometric mean NT_50_ values. Statistical significance was determined using the two-tailed Wilcoxon matched-pairs test. Experiments were repeated in triplicate. Dotted lines indicate the limit of detection. Right: fold decrease in neutralization for each variant relative to WT for each cohort of plasma samples (calculated from the left datasets) is shown. **(C)** IgG endpoint antibody responses specific to the N, RBD and S of WT SARS-CoV-2 were measured in plasma samples collected from cohorts as described earlier. **(D)** Fold decrease in specific binding to the RBD, NTD and S for each variant over WT for each cohort of plasma samples as described above. **(E)** IgA, IgM and IgG endpoint antibody titers specific to the S of WT SARS-CoV-2 or its variants in plasma samples collected from vaccinees before and 4 weeks after the 3^rd^-dose immunization. **(F)** Neutralizing titers against live SARS-CoV-2 WT, P.1, B.1.351 and B.1.617.2 for plasma from vaccinees before and 4 weeks after the 3^rd^-dose immunization. Black bars and indicated values represent geometric mean NT_50_ values. **(G)** Longitudinal neutralizing titers of plasma from 3-dose vaccinees at days 0, 7, 14, 28, 90 and 180 post the 3^rd^-dose vaccination. The geometric mean NT_50_ values are labeled. **(H)** Kinetics of the 3^rd^-dose booster elicited recall response as indicated during monitoring of NAb titers at different time points. The green and blue curves show the changes in kinetics of NAb titers for pre-3^rd^-dose and post-3^rd^-dose vaccination, respectively.

To seek information on potential binding-neutralization correlates, the abilities of antibodies present in plasma to bind the receptor-binding domain (RBD), N-terminal domain (NTD), S-trimer and nucleoprotein (N) from SARS-CoV-2 and its variants were measured by enzyme-linked immunosorbent assay (ELISA). As expected, all COVID-19 convalescents and vaccinees exhibited high anti-RBD, anti-NTD, anti-S and anti-N titers for SARS-CoV-2 variants, but weak antibody reactivity to SARS-CoV (Fig. 1C and fig. S1). Unexpectedly, the amount of N-specific IgG elicited by 2-dose and 3-dose vaccination schedules was 2-6-fold lower than those of convalescents, and 2-6-fold lower than the antibodies targeting S or RBD in vaccinees, reflecting distinct serological profiles (Fig. 1C and fig. S1). Overall plasma neutralizing activity against the WT was substantially correlated with anti-S and anti-RBD binding titers in ELISA. However, only marginal correlates between binding and neutralization potency were established for VOCs (fig. S2). In spite of this, a 3-dose administration elicits a broader range of antibody binding activities to VOCs with minimal decreasing folds than those of 2-dose vaccination and convalescents (Fig. 1D and fig. S2).

To evaluate the nature of humoral immune response elicited by a booster dose of CoronaVac, the S-specific IgA, IgM and IgG titers and neutralizing activities against SARS-CoV-2 variants were monitored before and 4 weeks after the third immunization. S-specific IgM and IgA titers were generally lower and were not significantly boosted in response to the third-dose vaccination (Fig. 1E). Similar to most convalescents (*2*), approximately 80∼90% of both anti-S IgG and NAb titers against the WT waned 6 months after the second vaccination (*13*), while the third-dose administration of CoronaVac boosted these titers by ∼20-fold at 4 weeks post vaccination (Fig. 1E and F). Significantly, vaccinees 6 months after the second immunization did not have detectable *in vitro* neutralizing activities against B.1.351, P.1 and B.1.617.2, while all vaccinees exhibited a robust recall humoral response to efficiently neutralize circulating variants post the third-dose vaccination (Fig. 1E and F). To further characterize the expeditiousness, longevity and immunological kinetics of recall response stimulated by the third-dose immunization, neutralizing potencies at days 0, 7, 14, 28, 90 and 180 post the third-dose vaccination were determined (Fig. 1G and H). Remarkably, NAb titer surged by ∼8-fold (from 7 to 53) at week 1, peaked by ∼25-fold increase (up to 177) at week 2 after the 3^rd^-booster and slowly decreased over time (Fig. 1G). Notably, NAb titer was maintained at around 60 on 180 days post the 3^rd^-booster, comparable to the high level of NAb titer elicited by the 2-dose administration (Fig. 1H). Taken together, these serological results reveal a third-dose booster can elicit an expeditious, robust and long-lasting recall humoral response.

The molecular mechanism underlying these potent, broad and durative antibody responses elicited by a third-dose booster 6 months after the administration of the second dose of the vaccine, might involve ongoing antibody somatic mutation and evolution of antibody by affinity maturation through prolonged and repeated antigen stimulation (*14, 15*). Although circulating antibodies derived from plasma cells wane over time, long-lived immune memory can persist in expanded clones of memory B cells (*16*). Thereby, we used flow cytometry to sort the SARS-CoV-2 S-trimer-specific memory B cells from the blood of seven selected CoronaVac vaccinees, including four samples from 3-dose vaccinees and three samples from 2-dose vaccinees (Fig. 2A and fig. S3). The averaged percentage of S-binding memory B cells in 3-dose vaccinees was substantially greater than those in 2-dose vaccinees (Fig. 2A and fig. S3). Due to differences in labeling strategies employed for sorting SARS-CoV-2-specific B cells, the above percentage of memory B cells was not directly comparable with those reported in naturally infected individuals and in mRNA vaccinated individuals. The gated double-positive cells were single cell sorted and immunoglobulin heavy (*IGH*; IgG isotype) and light (*IGL* or *IGK*) chain genes were amplified by nested PCR. Overall, we obtained 422 and 132 paired heavy and light chain variable regions from S-binding IgG^+^ memory B cells from four 3-dose and three 2-dose vaccinees, respectively (Fig. 2B and fig. S4). Surprisingly, expanded clones of cells comprised 45-61% of the overall S-binding memory B compartment in 3-dose vaccinees, which is approximately 2-fold higher than those in COVID-19 convalescents and in mRNA or 2-dose vaccinated individuals (Fig. 2B and C). When compared to 2-dose vaccinees, the increase in the number of persistent clones and various clonal compositions in 3-dose vaccinated group suggested an ongoing clonal evolution (Fig. 2B and C). Shared antibodies with the same combination of *IGHV* and *IGLV* genes in 3-dose vaccinees comprised ∼20% of all the clonal sequences. Similar to natural infection and mRNA vaccination (*2, 14, 16*), *IGHV3-30, IGHV3-53* and *IGHV1-69* remained significantly over-represented in 3-dose vaccinees (fig. S5). Meanwhile, notable differences in the frequency of human V genes between 3-dose vaccinated and the other two groups were observed as well (fig. S5). In 3-dose vaccinees, *IGHV3-21, IGHV4-39* and *IGHV7-4-1* were largely abundant, but *IGHV5-51, IGHV3-66* and *IGHV1-2* were significantly scarce when compared to the other two groups (fig. S5), indicative of memory B cell clonal turnover. Notably, large-scale, single-cell sequencing datasets generated from two cohorts of 2-dose, 3-dose vaccinees and a group of convalescents revealed no distinct preference in the frequency of *V* genes at total B cell repertoire level (fig. S6), suggesting that a large abundance of antibodies with low expression or affinities exist in B cells. Additionally, the number of nucleotide mutations in the *V* gene in 3-dose vaccinees is higher than those in both 2-dose vaccinees and naturally infected individuals assayed after 1.3 and 6.2 months, but slightly lower than those in convalescent individuals 1 year after infection (Fig. 2D), revealing ongoing somatic hypermutation of antibody genes. There was no significant difference in the length of the IgG CDR3 between vaccinated (either mRNA or inactivated) and convalescent (after 1.3 or 6.2 or months) groups (fig. S7). These results reveal that a third-dose booster 6 months after the second vaccination elicits an enhanced and anamnestic immune response, which is led by clonal evolution of memory B cell and ongoing antibody somatic mutations, resulting in enhanced neutralizing potency, breadth and longevity of the immune response against SARS-CoV-2.

**Fig. 2.**
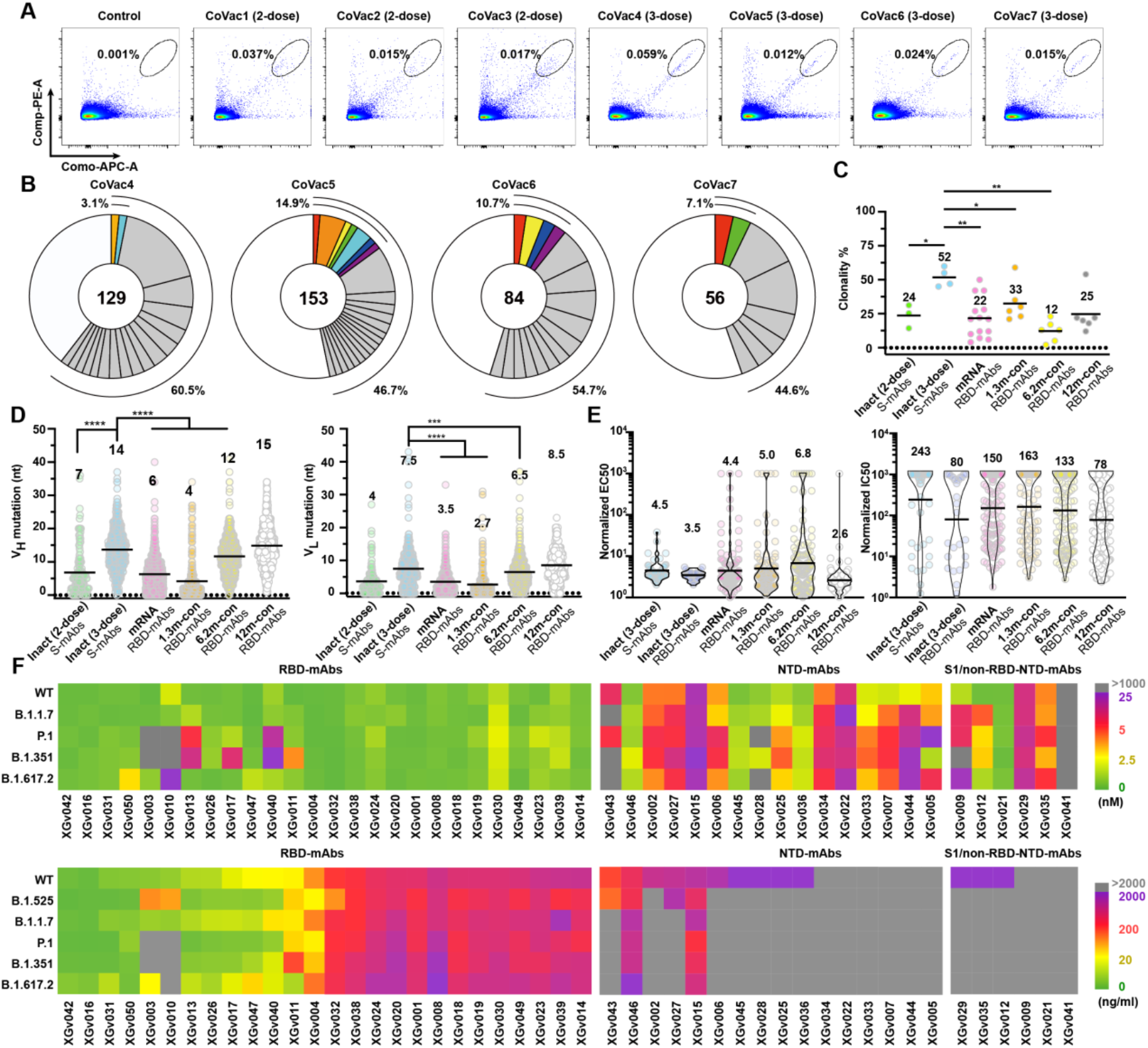
Memory B cell antibodies elicited by a 3^rd^-dose booster of an inactivated vaccine. **(A)** Representative flow cytometry plots showing dual allophycocyanin (APC)-S- and phycoerythrin (PE)-S-binding B cells for vaccinees and control donor. **(B)** Pie charts represent the distribution of antibody sequences from the four 3-dose vaccinees. The number in the inner circle is the number of sequences analyzed here. Pie-slice size is proportional to the number of clonally related sequences. The black outline indicates the frequency of clonally expanded sequences detected individually. Colored slices reveal clones that share the same *IGHV* and *IGLV* genes. **(C)** Graph shows relative clonality among seven individuals who received 2-dose or 3-dose of inactivated vaccines. Relative clonality for COVID-19 convalescents assayed at 1.3, 6.2 and 12 months after infection, as well as 2-dose mRNA vaccine recipients (*2, 14, 18*), previously described by Michel’s group, was compared. Black horizontal bars indicate mean values. Statistical significance was determined using two-tailed t-test. **(D)** Number of somatic nucleotide mutations in the *IGHV* (left) and *IGLV* (right) in antibodies from vaccinees, including 2-dose or 3-dose of inactivated vaccines and 2-dose of mRNA vaccines and COVID-19 convalescents assayed at 1.3, 6.2 and 12 months after infection (*2, 14, 18*). **(E)** Normalized ELISA binding (EC_50_) by antibodies isolated from the 3-dose inactivated and 2-dose mRNA vaccinees (ref) as well as COVID-19 convalescents to SARS-CoV-2 S trimer (left) and normalized pseudovirus neutralization activity (IC_50_) (right) against SARS-CoV-2 assayed at 1.3, 6.2 and 12 months after infection (ref). Among these, eight antibodies reported by Michel’s group were expressed and assessed for both binding by ELISA and pseudovirus neutralization activity for normalized comparison here. Black horizontal bars indicate mean values. **(F)** BLI binding affinities (upper panel) and pseudo-typed virus neutralization (bottom panel) by antibodies isolated from the 3-dose vaccinees to circulating SARS-CoV-2 variants. Color gradient for upper panel indicates *K*_*D*_ values ranging from 0 (green), through 2.5 (yellow) and 5 (red) to 25 nM (purple). Gray suggests no/very limited binding activity (>1000 nM). Color gradient for bottom panel indicates IC_50_ values ranging from 0 (green), through 20 (yellow) and 200 (red) to 2000 ng/ml (purple). Gray suggests no/very limited neutralizing activity (>2000 ng/ml).

To further explore the immunogenic characteristics of the antibodies obtained from memory B cells in 3-dose vaccinees, 48 clonal antibodies, designated as XGv01 to XGv50 (no expression for XGv37 and XGv48) were expressed and their antigen binding abilities verified by ELISA (fig. S8). Biolayer interferometry affinities (BLI) measurements demonstrated that all antibodies bound to WT SARS-CoV-2 at sub-nM levels (fig. S9 and table S1). The normalized geometric mean ELISA half-maximal concentration (EC_50_) revealed that all antibodies (EC_50_=4.5 ng/ml) obtained from 3-dose vaccinees, in particular RBD-specific mAbs (EC_50_=3.5 ng/ml), possessed higher binding activities than RBD-mAbs from early convalescents (at 1.3 and 6.2 months after infection, EC_50_=5.0 and 6.8 ng/ml, respectively) and mRNA (EC_50_=4.4 ng/ml) vaccinated individuals (*2, 14-18*), but were comparable to those from late convalescent individuals (EC_50_=2.6 ng/ml) assessed at 12 months after infection (Fig. 2E). These results indicate the possibility of the loss of antibodies with low binding affinities over time or an ongoing increase in affinity under the repeated exposures of antigen. Among these antibodies tested, 26 bound to RBD, 16 targeted NTD, and 6 interacted with neither RBD nor NTD, but bound S1 (S1/non-RBD-NTD) (fig. S9 and table S1). Pseudovirus neutralization assay revealed that all RBD-specific antibodies, 10 (∼60%) of the 16 NTD-directed antibodies and 3 (∼50%) of the 6 S1/non-RBD-NTD antibodies were neutralizing, presenting a relatively high ratio for NAbs (Fig. 2F, fig. S10 and table S2). Authentic SARS-CoV-2 neutralization assay results largely verified their neutralizing activities, albeit with that higher concentrations were required for some NAbs (fig. S11). Compared to RBD antibodies, many NTD NAbs exhibited very limited neutralizing activities. Notably, approximately 30% of RBD antibodies showed extra potent activities with half-maximal inhibitory concentration values (IC_50_) < 0.1 nM. In line with binding affinity, the normalized geometric mean IC_50_ of the RBD antibodies of 3-dose vaccinees was 80 ng/ml, substantially lower than those from naturally infected individuals (ranging from 1.3 to 6.2 months, IC_50_=130-160 ng/ml) and mRNA vaccinated individuals (IC_50_=150 ng/ml), but similar to those from late convalescents (IC_50_=78 ng/ml) (Fig. 2E) (*2, 14-18*). The overall increased neutralizing potency might have resulted from the ongoing accumulation of clones expressing antibodies with tight binding and potent neutralizing activities. Our experimental observations are consistent with a more recent study where antibodies generated from clonal B cells after 12 months showed enhanced neutralizing activities (*14, 15*).

To examine the cross-reactivity against VOCs and other human coronaviruses, binding responses of these antibodies to WT, B.1.1.7, P.1, B.1.351, B.1.617.2, SARS-CoV, HuCoV NL63, HuCoV 229E and HuCoV HKU1 were measured. All but 2 of the 48 antibodies showed strong cross-binding to SARS-CoV-2 VOCs and about one-third of antibodies exhibited clear cross-reactivity to SARS-CoV, but none of these bound to HuCoV NL63, HuCoV 229E or HuCoV HKU1 (fig. S12). For ∼ 20% and 25% of RBD- and NTD-targeting antibodies, respectively, binding affinities against B.1.351/B.1.617.2 were over 10-fold reduced compared with WT (Fig. 2E). To further determine the neutralization breadth, the neutralizing activity of these antibodies was assayed against five VOCs and SARS-CoV. Out of 26 RBD NAbs, 24 possessed cross-neutralization activity against all five SARS-CoV-2 VOCs (Fig. 2F and fig. S13). Among these, six RBD antibodies could cross-neutralize SARS-CoV, of which 2 exhibited more potent neutralization activity against SARS-CoV with IC_50_ values of 41 and 73 ng/ml. However, most of the NTD and S1/non-RBD-NTD NAbs lost their abilities to inhibit viral infection (Fig. 2F and fig. S13), indicative of higher variations for the NTD in VOCs. In comparison with NAbs from early convalescents, antibodies isolated from 3-dose vaccinees showed overall enhanced neutralizing potency and breadth to VOCs.

RBD is one of the main targets of neutralization in SARS-CoV-2 and other coronaviruses. Due to its inherent conformational flexibility, RBD exists in either an “open” (ACE2 receptor accessible) or “closed” (ACE2 receptor inaccessible) configuration (*19, 20*), bearing antigenic sites with distinct “neutralizing sensitivity”. To dissect the nature of the epitopes of RBD targeted by NAbs, 171 SARS-CoV-2 RBD-targeting NAbs with available structures (*2, 15, 21-82*), including 8 cryo-EM structures determined in this manuscript (fig. S14-S15 and table S3), were examined. By using cluster analysis on epitope structures, the antibodies were primarily classified into six sites (I, II, III, IV, V and VI) (Fig. 3A and fig. S16), that are related to the four or five classes assigned in recent studies (*22, 31*). Additionally, we superimposed structures of RBDs from these complex structures and calculated the clash areas between any 2 NAbs (Fig. 3B). Both strategies yielded identical results. Combining the results of the characterization of binding and neutralization studies reported previously with those determined here, the key structure-activity correlates for the six classes of antibodies were analyzed (Fig. 3). Antibodies with sites I, II and III, most frequently elicited by SARS-CoV-2 early infection, target the receptor-binding motif (RBM), and potently neutralize the virus by blocking the interactions between SARS-CoV-2 and ACE2 (Fig. 3C and D). Class I antibodies, mostly derived from *IGHV3-53/IGHV3-66* with short HCDR3s (generally <15 residues), recognize only the “open” RBD, and make contact with K417 and N501, but not L452/T478/E484 (Fig. 3C and D, and fig. S16-S17). Notably, mutations such as K417N, L452R, T478K, E484K and N501Y, or combinations of these mutations, identified in several VOCs like B.1.1.7, B.1.617.2, P.1 and B.1.351, have been demonstrated to be key determinants for the viral escape of neutralization by many NAbs (fig. S18) (*1, 81*). Approximately ∼75% and 60% of class I NAbs were significantly impaired in binding and neutralizing activities against B.1.351 as well as P.1, respectively, due to the combined mutations of K417N/T and N501Y (Fig. 3D and E, and fig. S18). Contrarily, Class III antibodies that are encoded by *IGHV1-2* and other variable heavy (VH)-genes and bound to RBD either in “open” or “closed” conformation, extensively associate with E484, and partially with L452, but not K417/T478/N501 (Fig. 3D and fig. S17C). Interestingly, *IGHV3-53/IGHV3-66* RBD antibodies with long HCDR3s (>15 residues) switch their epitopes from the site I to site III, indicating a clear antigenic drift during the process of somatic hypermutations (fig. S17C). Disastrously, over 90% class III antibodies showed a complete loss of activity against B.1.351 as well as P.1 largely owing to an E484K mutation (Fig. 3E). Against B.1.617.2, the substantially decreased activity of ∼half of the class III antibodies is presumably mediated by L452R (Fig. 3E). Class II antibodies use more diverse VH-genes and target the patch lying between sites I and III (Fig. 3D and fig. S19). Surprisingly, antibodies binding to site II possess relatively lower specificity in recognition of epitope clusters ranging from K417, L452, S477, E484 to N501 (fig. S16). Like site I, site II can only be accessed when the RBD is in “open” conformation (Fig. 3A). As expected, the effects of mutations on the activity of class II antibodies were severe, two-thirds of these antibodies had >10-fold fall in neutralization activities against VOCs (Fig. 3E). Overall, the above analysis reveals that the RBD mutations identified in several VOCs can significantly reduce and, in some cases, even abolish the binding and neutralization of classes I to III antibodies, albeit being the most potent neutralizing antibodies against WT SARS-CoV-2.

**Fig. 3.**
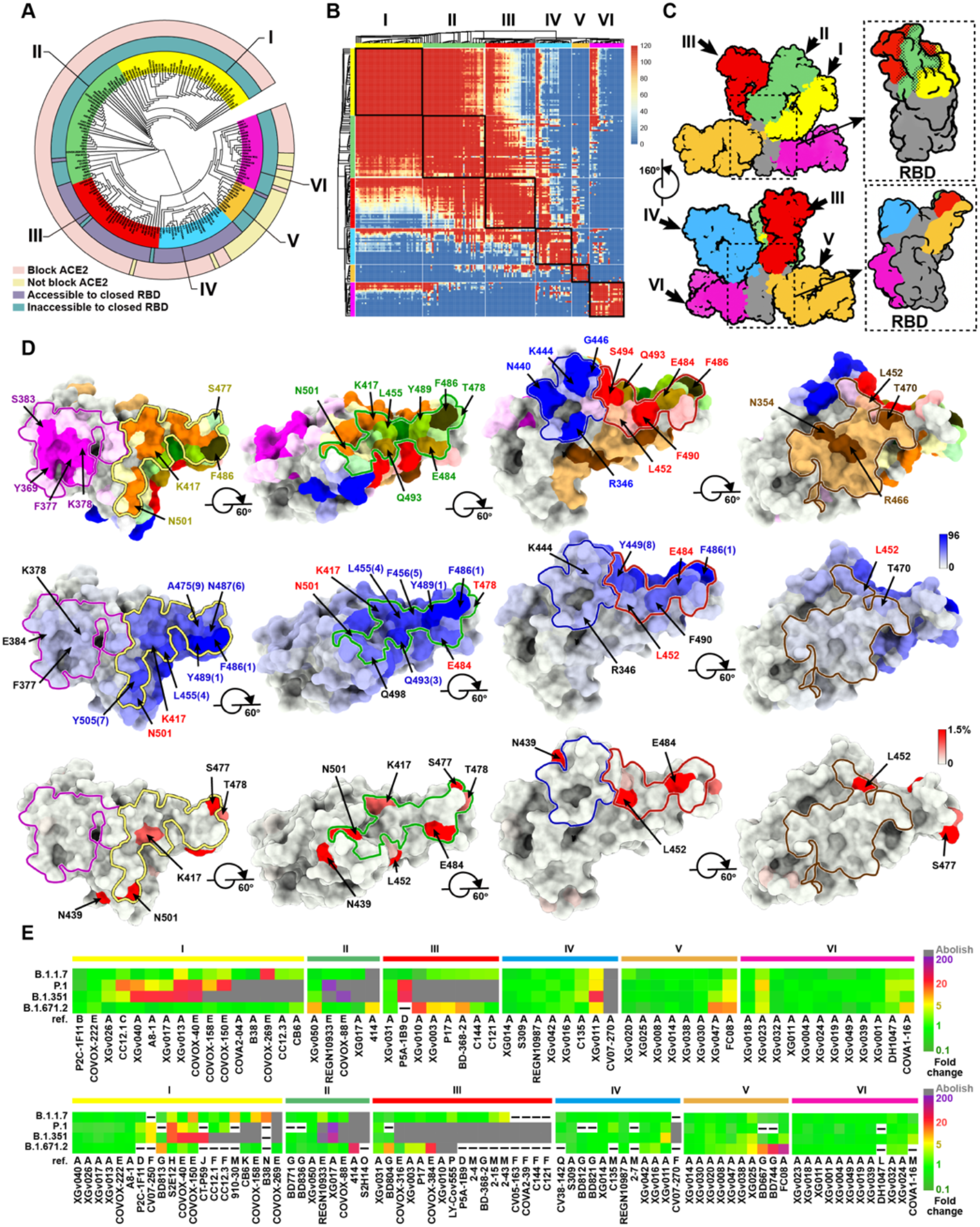
Structural landscape and immunogenic features of RBD NAbs. **(A)** Structure-based antigenic clustering of SARS-CoV-2 RBD NAbs. A total of 171 RBD NAbs with available structures were classified into six clusters (I, II, III, IV, V and VI). NAbs that can block ACE2 binding or not are outlined by light pink and light yellow, respectively. NAbs that can attach to the closed RBD or not are outlined by gray blue and gray green, respectively. **(B)** Superimposition matrix of 171 RBD NAb structures’ output from clashed areas (Å^2^) between variable regions of any two Fab fragments showing the clustering into six antibody classes. **(C)** Surface representative model of six types of NAbs bound to the RBD. Fab fragments of six representative antibodies are shown in different colors and the RBD is colored in gray. Insets illustrate the antigenic patches targeted by six representative antibodies. Dashed dots indicate the overlaps between two adjacent antigenic patches. **(D)** Structural landscapes of the six classes of RBD NAbs (upper panel). Antigenic patches (with targeting frequency >30%) recognized by six classes of NAbs are outlined in the assigned color scheme (same to Fig. 3C), among which residues with “hot targeting frequency” (generally over 65%, but over 85% in class I) are shown in bright colors corresponding to the patches they belong to. Residues involved in two (such as Y489, L452) or three (such as F486) neighboring antigenic patches are presented in a mixed color. Representative “hot” antigenic residues are labeled. Middle: hot map for antigenic residues on the RBD. Per residue frequency recognized by the 171 NAbs were calculated and shown. The top 9 of the hottest antigenic residues and key residues with substitutions in several VOCs are marked and labeled. Bottom: hot map for circulating variants with mutations on the RBD. Mutation frequency for each residue was calculated based on the datasets from GISAID. **(E)** Immunogenic characteristics of six classes of RBD-targeting NAbs. Hot maps show relative fold changes in *K*_*D*_ values (up) and IC_50_ values (down) against several VOCs for the six classes of NAbs, including previously reported (*97-108*) and newly isolated antibodies described in this manuscript. Color gradients for upper and bottom panels indicate relative fold changes and are shown at right side. “-”: no related datasets in the original studies and related references are listed. Ref “A” indicates that the datasets were produced in this manuscript. Other letters in Ref correspond to different reference numbers shown as below. B – 91 and this manuscipt, C – 99 and this manuscript, D – 97, E – 30, 81, 103 and 104, F – 99, G – 98, H – 100 and 108, J – 101, K – 94 and 102, L – 105 and 106, M – 94, N – 105, O – 107, P – 82, Q – 66, respectively.

By contrast, antibodies of the other three classes recognize evolutionarily conserved regions distinct from the RBM and some of these are often cross-reactive with other sarbecoviruses (*65-67, 79*). The binding of class IV antibodies, albeit attached to the apical shoulder of the RBM, is focused on a condensed patch that comprises residues 345-346, 440-441, 444-446, 448-450, which are not related to mutations observed in VOCs (Fig. 3C and fig. S16). Related to the binding position, site IV epitopes, accessible in both “open” and “closed” conformations, exist either as partially overlapped with or outside ACE2 binding sites (Fig. 3A). Interestingly, class IV antibodies can execute their neutralizations via multiple mechanisms, such as (i) direct blockage of RBD-ACE2 associations, (ii) bridging adjacent “closed” RBDs to lock the S-trimer into a completely closed prefusion conformation, (iii) blockage of viral membrane fusion by locking conformational changes of the S-trimer, or (iv) Fc-dependent effector mechanisms (*31, 62, 67*). Class IV antibodies, e.g. 1-57, 2-7, S309 and BD-812, hold the greatest potential for harboring ultra-potent neutralization activity and markedly high tolerance to most VOCs (*63, 67*). Not surprisingly, all class IV antibodies, but CV07-270, exhibited excellent neutralizing breadth and potency to VOCs (Fig. 3E). The probable reason underlying the exception could be that CV07-270 bears an unusually long HCDR3, directly contacting E484, distal to the site IV (*46*). Site V locates beneath the RBM ridge, opposite to the site I, and adjacent to the site III. None of the class V antibodies compete with ACE2 binding (Fig. 3D and fig. S17). Due to ∼40% targeting frequency to L452, B.1.617.2, but not other VOCs, partially decreased the activities of some class V antibodies (Fig. 3E). Class VI antibodies recognize a patch on one side of the RBD, distal from the RBM. Among these, some compete with ACE2 binding, while some do not, and this largely depends on the orientation/pose of the antibodies bound. Both sites V and VI contain cryptic epitopes that are only accessible when at least one RBD is in the open state (Fig. 3A and C). In some cases, e.g. FC08 and CR3022, belonging to class V and VI, respectively, epitopes are only accessible in the prefusion S-trimer under the condition that all RBDs are open, suggesting that binding of these antibodies would facilitate the destruction of the prefusion S-trimer (*83, 84*). In spite of less potency, antibodies targeting sites V to VI are mostly tolerant to the VOCs.

Low levels of NAbs elicited by either natural infection or vaccination during *in vivo* viral propagation may impose strong selection pressure for viral escape, leading to an increase in the number of SARS-CoV-2 variants. To further understand the drivers of viral evolution, we constructed immunogenic and mutational heatmaps for RBD using the 171 NAb complex structures to estimate *in vivo* NAb-targeting frequencies on the RBD and viral mutation frequencies (calculated from the datasets in the GISAID), respectively (Fig. 3D and fig. S19). Briefly, for each antibody, we identified epitope residues and calculated the frequency of each RBD residue being recognized by antibody. Immunogenic heatmap revealed that the epitope residues of sites I to III showed predominantly higher NAb recognition frequencies (about 53.8, 55.0 and 49.2 antibodies per residue on average for site I, II and III, respectively) compared with those of sites IV to VI (about 19.4, 9.1 and 14.3 antibodies per residues on average for site IV, V and VI, respectively), suggesting that class I to III antibody epitopes are “hot” immunogenic sites (Fig. 3D and fig. S19). In line with this, residues within sites I to III exhibited dramatically higher mutation frequencies, as revealed in circulating variants that include mutations of K417, L452, S477, T478, E484 and N501 residues (Fig. 3D and fig. S19). Surprisingly, none of the top 9 hottest immunogenic residues had a high mutation frequency. In particular, residues, such as F486, Y489, Q493, L455, F456, *et*.*al* (top 5, having 96, 96, 81, 73 and 70 antibodies per residue, respectively) with large side chains exhibited extremely low mutation frequencies in circulating SARS-CoV-2 strains (Fig. 3D and fig. S20). It’s worthy to note that all these residues are extensively involved in the recognition of ACE2. The buried surface area (BSA) of these residues upon binding to ACE2 confirmed that extensive interactions would be significantly reduced by amino acid substitutions, thereby affecting ACE2-mediated viral entry. Thus, genetic, structural and immunogenic analysis explains why mutations at these positions would not be selected.

A few studies have reported that a subset of NTD-targeting antibodies can be as potent as best-in-class RBD specific antibodies. They work *via* inhibiting a step post-attachment to cells like blocking fusion of the virus to the host cell membrane (*85-88*). We performed cluster analysis on 26 structures of the NTD-NAb complexes (including 2 structures solved in this manuscript) (fig. S21A) (*54, 85-93*). A dominant site α, defined as the “supersite” in more recent studies (*85-88*), comprising of three flexible loops (N1, N3 and N5), is the largest glycan-free surface of NTD facing away from the viral membrane (facing up). Antibodies targeting site α generally exhibited the most potent neutralizing activity compared to other sites on the NTD (*85, 90*) (fig. S21B and C). The NTD supersite antibodies are primarily derived from a subset of VH-genes with an over-representation of *IGHV1-24*. Sites β and γ, as the left and right flank clusters, construct a shallow groove beneath the supersite and locate at the back of the groove, eliciting less potent antibodies. By contrast, δ antibodies, bound to a patch beneath the groove have their Fab constant domains directed downward toward the virus membrane (facing down) (fig. S21B and C). In line with binding orientation, many of the δ antibodies were shown to present infection enhancing activities *in vitro* (*54, 90*). Perhaps correlated with being a “hot” immunogenic site that is amenable to potent neutralization, highly frequent mutations, including a number of deletions within the NTD supersite were identified in most VOCs under ongoing selective pressure, leading to significant reduction and in some cases even complete loss of neutralization activity for these NTD supersite NAbs (*94*).

More recent studies have reported that SARS-CoV-2 infection can produce a long-lasting memory compartment that continues to evolve over 12 months after infection with ongoing accumulation of somatic mutations, emergence of new clones and increasing affinity of antibodies to antigens (*14, 15*). Consequently, an increase in breadth and overall potency of antibodies produced by memory B cells over time has been revealed (*14*), akin to the experimental observations elicited by a 3-dose vaccination strategy using an inactivated vaccine described in this study. To investigate whether changes in the frequency of distribution of the six types of RBD antibodies is associated with evolution time, we collated and categorized human SARS-CoV-2 NAbs from available literatures. For antibody clustering, we combined structural and square competition matrix analysis for 273 RBD NAbs in total (Fig. 4A and fig. S22). In the earliest documented studies (before Dec 2020), NAbs belonging to classes I to III were predominantly identified in early COVID-19 convalescent and 2-dose vaccinated individuals (defined as early time point), accounting for up to ∼80% of total antibodies. By contrast, a low ratio of NAbs from IV to VI was reported possibly due to their less potent activities at the early time point (Fig. 4A). In recent literatures (after Dec 2020), NAbs with enhanced neutralizing potency and breadth from IV to VI have substantially been enriched in the late convalescents or 3-dose vaccinees, almost equal in frequency to antibodies from I to III and further becoming ascendant in individuals immunized with 3 doses of inactivated vaccine (Fig. 4A). Differential frequency of distribution of antibody types may provide an additional possible explanation for the observed enhanced neutralizing breadth of plasma in late convalescent individuals and 3-dose vaccinees. These results suggest that memory B cells display clonal turnover after about 6 months, subsequently resulting in changes in the composition of antibodies in B cell repertoire and thereby partially contributing to enhanced activities of antibodies secreted in the plasma over time. To explore the underlying mechanism, we measured the binding affinities of 167 type-classified antibodies that are also further categorized into early and late time point groups (table S1 and fig. S9). For the late time group, there was a 10-20 fold increase in binding affinity for individual classes, compared to those in the early time point group (Fig. 4B). In early time point group, antibodies from IV to VI exhibited higher binding affinities to the RBD than those from I to III, in particular, antibodies from V and VI despite limited numbers (Fig. 4B). Possibly higher affinities for these antibodies are required to accomplish neutralization successfully. Thus, most antibodies from V and VI with low affinities and activities might be screened out in the early time point. In the late point group, sub-*nM* binding affinities for individual class antibodies with no distinct variations were observed, reflecting ongoing affinity maturation over time. This might also explain the observation that some antibodies, from I to III isolated in the late time point possess potent cross-neutralization activities (Fig. 3E). Our antibody clustering and *V* gene usage analysis suggests that individual class antibodies can be derived from multiple *V* genes and the shared *V* gene antibodies belong to different classes. To decipher the intrinsic trends in the relationship between binding affinity and somatic hypermutation (SHM) rate, we determined the relative affinity (*K*_*D*_) and calculated the SHM rate of antibodies that are encoded by the same *V* gene and belong to the same class. The measured *K*_*D*_–SHM plots and *K*_*D*_–SHM log-log plots of class I antibodies (n=61), including 32 NAbs derived from *IGHV3-53*, show least squares fitting of data to a power law with a strong correlation of −0.81 for *IGHV3-53* antibodies (−0.55 for all class I antibodies) (Fig. 4C). The absolute value of its slope corresponding to a free energy change per logarithm (base e) *SHM* of cal nmol^−1^, where free energy change is 4.98*RT* + 1.48*RT* In(*SHM*) (*R* = 2.0 cal K^−1^ nmol^−1^ and *T* = 298 K). Antibodies with adequate numbers tested from II and III exhibited similar trends by following a power law, among which *IGHV3-66* antibodies in class II yielded a compelling correlation of −0.94 despite 6 plots involved in the fitting (Fig. 4C). These trends indicate that as the SHM increase, the binding energy increases and *K*_*D*_ value decreases.

**Fig. 4.**
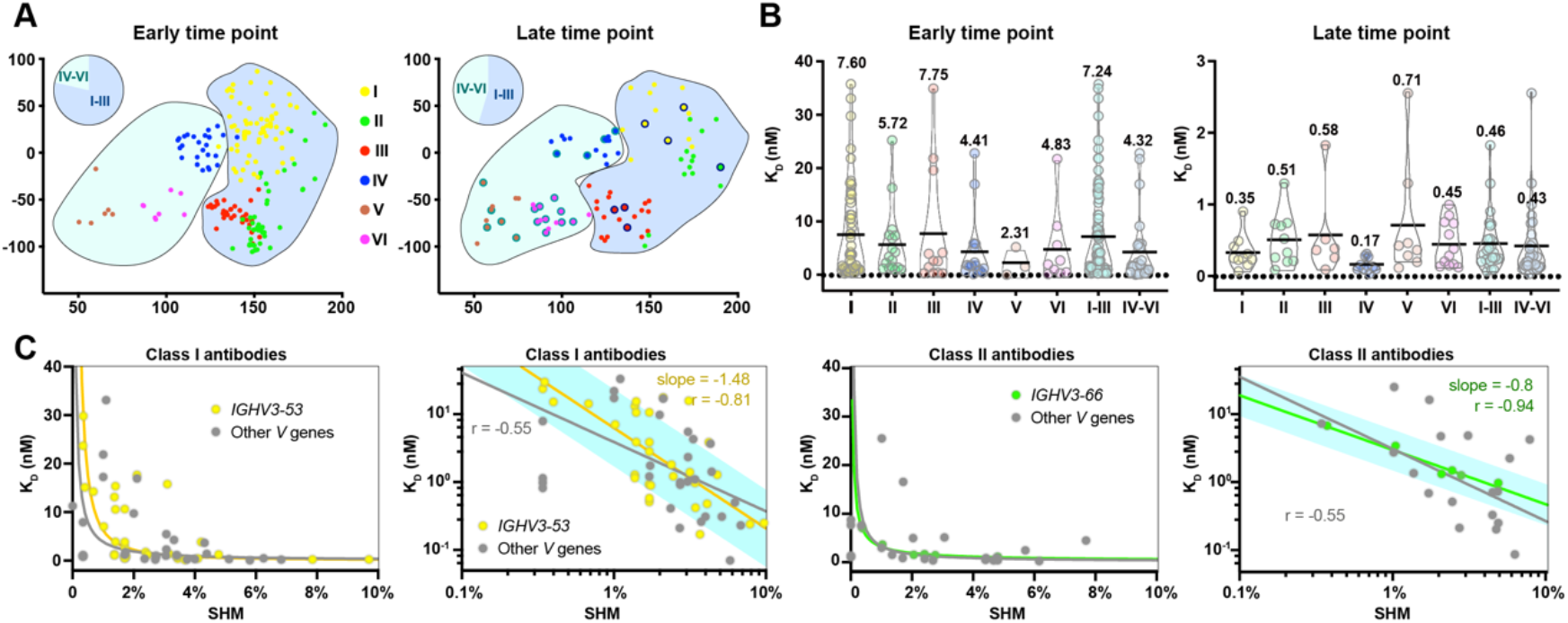
Antibody evolution and affinity maturation. **(A)** Uniform manifold approximation and projection (UMAP) plot displaying the antibodies defined as the early time point group (left) and late time point group (right). The antibodies are colored based on their cluster assignments by the hierarchical clustering algorithm. Antibodies from I to III and IV to VI are highlighted in cyan and gray blue background, respectively. Pie charts represent the frequency distribution of antibodies belonging to I to III and IV to VI. Antibodies isolated from 3-dose vaccinees are outlined by black lines. **(B)** Dissociation constants (*K*_D_) of the antibodies from I to VI. Individual class antibodies are represented in colors corresponding to the classes they belong to. The color scheme is same as Fig. 4A. BLI traces are shown in fig. S9. **(C)** The measured *K*_*D*_–SHM plots (left) and *K*_*D*_–SHM log-log plots (right) of antibodies from I and II are shown. *IGHV3-53* and *IGHV3-66* antibodies belonging to class I and II are colored in yellow and green, respectively. The straight curves and lines are the least squares fits of the data to the power law with the values of the slope for *IGHV3-53* and *IGHV3-66* antibodies. The black curves and lines indicate the fitting of antibodies from I or II; the yellow and green ones suggest the fitting of *IGHV3-53* and *IGHV3-66* antibodies, respectively. The cyan lines are the 90% predicted interval.

More recently, the B.1.617.2 variant has contributed to another surge in COVID-19 cases worldwide, accounting for ∼90% of new cases in the UK and >40% in the US, despite the fact that increasing number of people have been vaccinated. Evaluation of the effectiveness of several vaccines performed recently suggests that the efficacy for VOCs correlates with full vaccination status and the time that has passed since vaccination (*95, 96*). These may indicate that the effectiveness of the vaccines has started to decline as months pass after vaccination due to fading immunity. Our results demonstrate that a third-dose booster of inactivated vaccine can elicit an expeditious, robust and long-lasting recall humoral response which continues to evolve with ongoing accumulation of somatic mutations, emergence of new clones and increasing affinities of antibodies to antigens, conferring enhanced neutralizing potency and breadth. Collectively, our findings rationalize the use of 3-dose vaccination regimens.

## Data Availability

Cryo-EM density maps of the SARS-CoV-2 S trimer in complex with XGv013 or XGv043, the SARS-CoV-2 S trimer in complex with XGv004, XGv030 and XGv016; the SARS-CoV-2 S trimer in complex with XGv026 and XGv046, and the SARS-CoV-2 S trimer in complex with XGv018, XGv038 and XGv42 have been deposited at the Electron Microscopy Data Bank with accession codes EMD-UUUU, EMD-WWWW, EMD-XXXX, EMD-YYYY and EMD-ZZZZ, respectively

## Acknowledgments

We thank Dr. Xiaojun Huang, Dr. Boling Zhu, Dr. Lihong Chen, Dr. Xujing Li and Dr. Gang Ji for cryo-EM data collection, the Center for Biological Imaging (CBI) in Institute of Biophysics for EM work and thank Dr. Yuanyuan Chen, Zhenwei Yang for technical help with BLI experiments. We also thank Prof. David Ho and Prof. Barton Ford Haynes for generous gift of the sequences of two reference mAbs. Work was supported by the Strategic Priority Research Program (XDB29010000, XDB37030000), CAS (YSBR-010), National Key Research and Development Program (2020YFA0707500, 2018YFA0900801), Emergency Key Program of Guangzhou Laboratory, Grant No. EKPG21-09 and Beijing Municipal Science and Technology Project (Z201100005420017). Xiangxi Wang was supported by Ten Thousand Talent Program and the NSFS Innovative Research Group (No. 81921005).

## Author contributions

X.W., Q.W., Y.W., X.S.X and Y.C. conceived, designed and coordinated the experiments. Y.Z., A.Y., Y.W., Q.Z. and J-B.W. cloned and produced antibodies. K.W., Z.J., K.F., J.M., Y.J. and L.Q. expressed and purified all recombinant antigen proteins used in this manuscript. J-J.W., Y.H., L.W., J.L., X.G., Y-J.Z., H.Z. and B-S.L performed pseudovirus and authentic virus neutralization experiments. K.W., L.W., P. L., WJ.F, N.W and L.Z performed structural study. Y.C., Y.Z., W.W. and Y.G. prepared PBMCs and flow cytometry sorting. Y.C. and A.Y. performed 10X sequencing library construction. Z.J. and K.F performed BLI assay. Y.H. and Q.G recruited volunteers and coordinated the collection of blood samples. All authors analyzed data; X.W wrote the manuscript with input from all authors.

