## Supplemental Materials for "A third dose of inactivated vaccine augments the potency, breadth, and duration of anamnestic responses against SARS-CoV-2"

**This PDF file includes:**

Materials and Methods

Figs. S1 to S22

Tables. S1 to S3

### Materials and Methods

#### Facility and ethics statements

All experiments with live SARS-CoV-2 viruses were performed in the enhanced biosafety level 3 (P3+) facilities in the Sinovac Biotech Ltd approved by the National Health Commission of the People's Republic of China.

#### Cell lines

Vero cells (ATCC, CCL-81) and HEK293T cells (ATCC, CRL-3216) were cultured in Dulbecco's Modified Eagle's Medium (DMEM) supplemented with 10% fetal bovine serum (FBS). The cultures were maintained at 37 °C in an incubator supplied with 5% $CO_2$.

#### Viral stocks

The SARS-CoV-2 wild type strain CN01 was isolated from a patient in China during the early phase of the COVID-19 endemic. The SARS-CoV-2 variants of concern (VOC) beta (B.1.351 lineage), was isolated from a patient traveling back from South Africa; VOC gamma (P.1 lineage) was isolated from a person in Brazil; and the newly emerged VOC delta (B.1.617.1 lineage), was isolated from a traveller with PCR-confirmed infection from India in early June. All the viruses were subjected to three rounds of plaque-purifications with clones from each passage sequenced for verification as described previously (*9, 10*). The final clonal of the virus was inoculated into Vero cells and grown to 95% confluence at a MOI of 0.01. When 98% of the cells developed visible cytopathic effect (CPE) after 72 hours incubation at 37 °C, the cultures were harvested and centrifuged at 10,000 × g at 4 °C for 20 mins to remove the debris and preserved at -80 °C as viral stocks. Then, a small fraction of the stock was thawed at room temperature for viral titration.

#### Human sera

All the convalescent sera were taken from individuals recovered from COVID-19 30 – 45 days after confirmation by RT-PCR. All the vaccine sera were collected from volunteers who received two doses or three doses of the WHO-approved inactivated SARS-COV-2 vaccine (CorovaVac, Sinovac, China).

**Protein expression and purification**

The plasmids encoding the full-length spike (S) protein (residues 1-1208), receptor-binding domain (RBD) (residues 319-541) and N-terminal domain (NTD) (residues 1-304) of wild-type SARS-COV-2 (GenBank: MN908947) were constructed as described previously (*71*). These plasmids were used as templates for the construction of S, RBD and NTD of the variants of concern - B.1.1.7 (with mutations of 69-70 del, 145 del, N501Y, A570D, D614G, P681H, T716I, S982A and D1118H), B.1.351 (with mutations of 242-244 del, L18F, D80A, D215G, K417N, E484K, N501Y, D614G and A701V), P.1 (with mutations of L18F, T20N, P26S, D138Y, R190S, K417T, E484K, N501Y, D614G, H655Y and T1027I) and B.1.617.2 (with mutations of T19R, G142D, 156del, 157del, R158G, L452R, T478K, D614G, P681R, D950N) by overlapping PCR. All the full-length S gene constructs have two proline substitutions at residues 986 and 987, a 'GSAS' substitution at the furin cleavage site and a C-terminal T4 fibritin foldon domain to facilitate the protein expression and stabilization of the trimer conformation. All the constructs described above were attached with a C-terminal twin-strep-tag II for protein purification. To obtain the S, RBD or NTD of interest, the plasmid constructed above was transiently transfected into HEK293 F cells grown in suspension at 37 °C in a rotating, humidified incubator supplied with 8% $CO_2$, maintained at 130 rpm. After incubation for 72 hours, the supernatant was harvested, concentrated and exchanged into the binding buffer by tangential flow filtration cassette. The protein of interest was separated by affinity chromatography using resin attached with streptavidin and further dialyzed into a buffer containing 20 mM Tris pH 8.0 and 200 mM NaCl.

**Collection of Human Peripheral Blood Mononuclear Cells**

Volunteer recruitment and blood draw were approved by the institutional Ethics Committee of the School of Basic Medical Sciences, Fudan University (2020-C007). Study participants, four donors receiving three doses of CoronaVac inactivated vaccines and three donors receiving two doses, donated blood one month after vaccination. All donors ranged in age from 23-52, and the female:male ratio was 3:4. After collection of peripheral blood, human peripheral blood mononuclear cells (PBMCs) were isolated, aliquoted, and stored in liquid nitrogen.

**Single Memory B Cell Sorting and Antibody Cloning**

Single memory B cell sorting and antibody cloning from the sorted single B cells were performed as described previously (*17*). Briefly, stored PBMCs were thawed and incubated with CD19 MicroBeads (Miltenyi Biotec). CD19+ B lymphocytes were then incubated sequentially with human Fc block (BD Biosciences), anti-CD20-PECy7 (BD Biosciences), S-ECD-PE, and S-ECD-APC. The single memory B cells (CD20-PECy7+ S-ECD-PE+ S-ECD-APC+) were then sorted into 96-well plates using a FACSAria II (BD Biosciences), and used for antibody cloning as previously reported (*17*). Amplified PCR products of Fab regions of immunoglobulin heavy and kappa/lambda light chains were subjected to electrophoresis and Sanger sequencing. Their nucleotide sequences were analyzed by IMGT/V-QUEST and IgBlast, and the V(D)J gene segment and CDR3 sequences of each antibody were determined.

**Antibody Expression**

The selected antibodies were subjected to vector construction and antibody expression as reported previously (*17*). Briefly, all the cloned human monoclonal antibodies were prepared by transient transfection in mammalian HEK293F cells, which were cultured using serum-free OPM-293-CD05 medium (OPM Biosciences) at 37°C under 5% CO2 with shaking at 100 rpm.

**Production of Fab fragment**

To generate the Fab fragments, the purified Mabs XGv013, XGv043, XGv004, XGv030, XGv016, XGv026, XGv046, XGv018, XGv038 and XGv042 were processed using the Pierce FAB preparation kit (Thermo Scientific) as described previously (*71*). Briefly, the samples were first applied to desalination columns to remove the salt. After centrifugation, the flow through was collected and incubated with beads attached with papain to cleave Fab fragments from the whole antibodies. Then the mixtures were transferred to protein A affinity column which specifically binds the Fc fragments of antibodies. After centrifugation, the Fab fragments were obtained and dialyzed into Phosphate Buffered Saline (PBS) (ThermoFisher, catalog #10010023).

**Authentic virus neutralization assay**

The plasma samples collected from both the convalescents and vaccinated volunteers were inactivated first at 56 °C for 0.5 h. The inactivated serum samples or purified mAbs were diluted by serial dilution from 1: 4 or 50,000 ng/mL with cell culture medium in two-fold steps and mixed with a virus suspension containing 100 TCID50 and incubated at 36.5 °C for 2 h. Afterwards, the mixtures were added to the 96-well plates seeded with confluent Vero cells and incubated for another 5 days at 36.5 °C in an incubator supplied with 5% $CO_2$. Cytopathic effect (CPE) of each well was observed and recorded under microscopes by three different individuals, and then used for the calculation of neutralizing titers by the Reed–Muench method.

**Pseudovirus neutralization assay**

The pseudotyped viruses were produced, aliquoted, and stored as described previously (*71*). The generation of pseudotyped SARS-CoV-2 variants of concern was performed similarly except using the plasmids with the corresponding S protein mutations. In vitro neutralization assay using pseudoviruses was performed as described previously (*71*). Briefly, the Huh-7 cells were seeded and then subjected to incubation with pseudovirus/antibody mixture for 12 hours. The antibodies were serially diluted 1: 3 in PBS for nine dilutions in total, with the beginning concentration of 10 μg/ml. Fresh DMEM medium was used to replace the mixture for further culture. After 24 or 48-hour, the Huh-7 cells were collected and luminescence was measured as described previously (*17*).

**ELISA and Competition ELISA**

The ELISA and competitive ELISA assays were performed as reported previously (*17*). For ELISA, 96-well plates were coated with antigen proteins (10 μg/ml) and then blocked with 2% BSA in PBS. The first antibody was serially diluted 1: 3 in PBS for eight dilutions in total, with the beginning concentration of 10 μg/ml, for incubation. Following the incubation and visualization with HRP-conjugated second antibody (Thermo Fisher Scientific), the area under the curve (AUC) was then calculated (PRISM) to evaluate the antigen-binding capacity. For competitive ELISAs, 96-well plates were coated with antigen proteins (2 μg/ml) and then blocked with 2% BSA in PBS. Incubation with first blocking antibody (15 μg/ml) was followed by directly adding biotinylated second antibodies (0.25 μg/ml). Streptavidin-HRP (BD Biosciences) was then added for detection. Samples with no first antibody were used as a negative control for normalization.

**Bio-layer interferometry**

Bio-layer interferometry **(**BLI) experiments were carried out on an Octet Red 96e machine (Fortebio). To measure the binding affinities of monoclonal antibodies with native and variant RBDs, antibodies were immobilized onto protein A biosensors (Fortebio), while serial dilutions of RBD or its variants were used as analytes. Data were recorded using software Data Acquisition 11.1 (Fortebio) and analyzed using software Data Analysis HT 11.1 (Fortebio) with a 1: 1 fitting model.

**Cryo-EM sample preparation, data collection and processing**

The purified S protein was mixed and incubated with the Fab fragments of XGv013 and XGv043, XGv004, XGv030 and XGv016, XGv026 and XGv046, as well as XGv018, XGv038 and XGv42 with a molar ratio of 1 : 1.5 (S protein to Fab) to obtain the S-Fab-complexes. Then, three-microliter aliquot of each complex was deposited onto the glow-discharged holey carbon-coated gold grid (C-flat, 300-mesh, 1.2/1.3, Protochips In.), blotted for 7 seconds in 100% relative humidity and plunged into the liquid ethane using Vitrobot (FEI) (3,4). Cryo-EM data sets were collected at 300 kV with a Titan Krios microscope (FEI). Movies (32 frames, each 0.2 s, total dose of 60 $e^{-}$ $Å^{-2}$) were recorded using a K2 Summit direct detector with a defocus range between 1.5-2.7 μm. Automated single particle data acquisition was performed by SerialEM, with a calibrated magnification of 22,500 yielding a final pixel size of 1.04 Å.

**Model fitting and refinement**

Coordinates for initial complexes were generated by docking individual chains from reference structures into cryo-EM density using UCSF Chimera. (S trimer, PDB number 6VXX; Fab, PDB number 7CAK). Models were then adjusted manually using Coot and automatically refined into cryo-EM maps by rigid-body and real-space refinement in Phenix.

**PISA and clustering**

All the published structures of SARS-CoV-2 neutralizing antibodies in complex with S trimer or its subdomain (RBD or NTD) were obtained from the Protein Data Bank and were superimposed on RBD individually. The buried surface areas (BSA) of each pairwise antibody were recorded as the distance between them. Afterwards, the clustering of BSA matrix was performed with pheatmap in R.

**Epitope identification and Clustering**

All the published structures of SARS-CoV-2 neutralizing antibodies in complex with S trimer or its subdomain (RBD or NTD) were obtained from the Protein Data Bank. All residues on RBDs and NTDs with distances within 4 Å from their corresponding antibodies were recorded as the epitope residues. The binding frequency of each residue was calculated for the generation of epitope heatmap. The binary binding situation of each residue (aa 319-541 for RBD and aa 2-310 for NTD) were used to calculate the distance between pairwise sequences with an in-house python script. The clustering of sequence distance matrix was performed by fetch in PHYLIP.

**Prediction of NAbs' class from ELISA assay.**

K-means algorithm was used to classify NAbs. Briefly, we first calculated the ratio of NAbs that didn't clash with each other for all the 6 classes. The results were stored in the matrix *S*. Then, the data of 12 NAbs belonging to 6 known classes was utilized to generate initial seeds of the following K-means clustering. This process was achieved by MATLAB script *four_template.m.* In consideration of no such significant differences for templates data for Classes I to III, antibodies from these three classes yielded from the first round of clustering needs to be further analyzed. After that, Sum of Squares for Error (SSE) between a certain NAb and the template in the same class was calculated. NAbs corresponding to large SSE (>0.5) had a low confidence level of right clustering validity and would be reclassified. The consistency of matrix $S$ and data of ELISA assay were the basis for the second round of clustering. We found the best class for the reclassified NAbs by the MATLAB script *class_abnormal.m.*

**Calculation of positional mutation frequency**

The frequencies of positional mutations were calculated based on the SARS-CoV-2 spike sequences deposited in GISAID by April 6th, 2021 (Elbe and Buckland-Merrett, 2017). All the sequences were aligned pairwise with the wide-type strain, the mutation frequencies for each position were calculated as the proportion of total mutations in all deposited sequences. The low quality 'X' residues were not counted as mutations.

**High-throughput single-cell mRNA, VDJ, and feature barcode sequencing**

As previously described, SARS-CoV-2 antigen-specific B cells from 2-dose and 3-dose CoronaVac vaccinees' PBMC were isolated through MACS and FACS, and subjected to microfluidic-based single-cell sequencing (10x Genomics, Libra-seq) to obtain 5' mRNA, VDJ, and feature barcode information *(72)*. Briefly, B cells were first enriched from frozen PBMCs by negative selection using the EasySep™ Human B Cell Enrichment Kit (STEMCELL, 17954). Purified B cells were stained with antibody cocktails containing oligo-barcoded antigens (RBD and NTD, PE/APC dual labeled for each antigen) on ice for 30 minutes *(72)*. Stained B cells were collected on an Astrios EQ (BeckMan Coulter) for single CD14–, CD16–, 7-AAD–, CD19+, antigen+ cells. Collected cells were subjected to 10× Chromium System (10× Genomics) according to the manufacturer's instructions for Libra-seq library construction. Sequencing was performed on the Illumina xten platform for 2x150bp dual index sequencing.

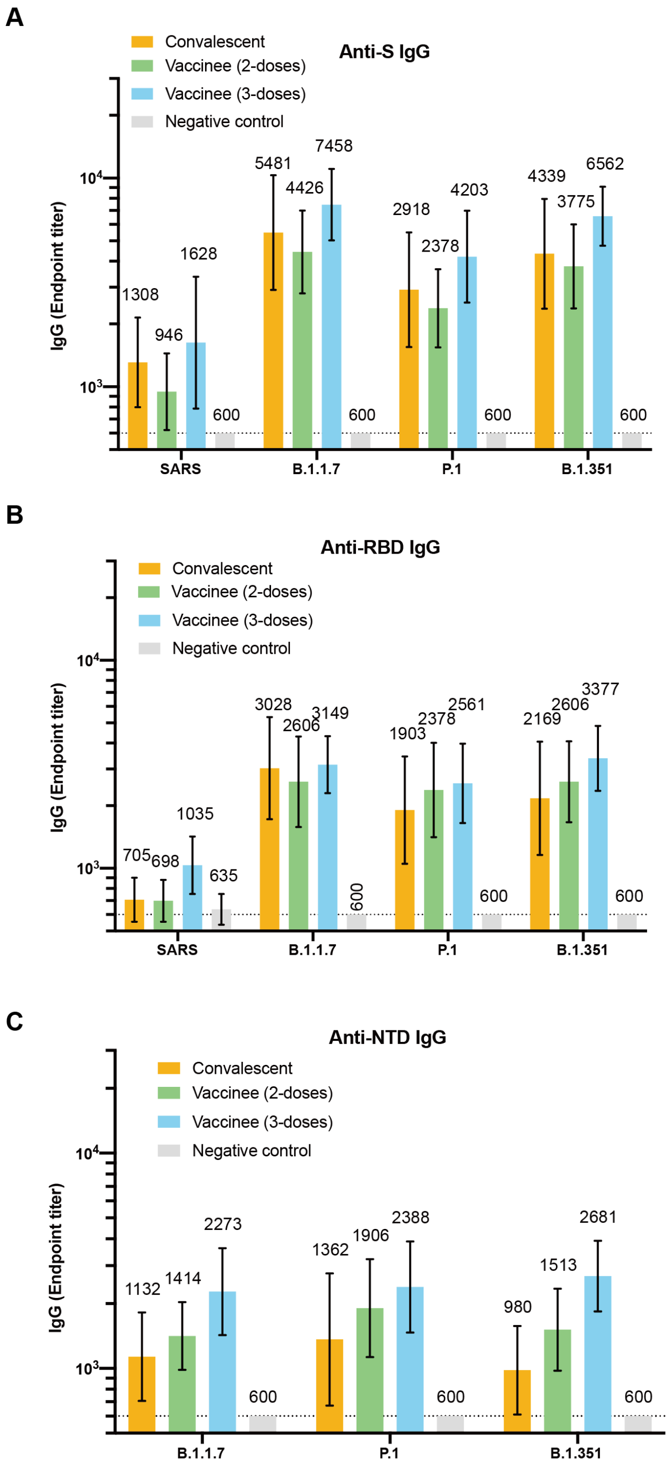

**Fig. S1 Cross-binding activity of plasma samples**

Results of ELISAs measuring the cross-binding activity of plasma against S, RBD and NTD proteins of control individuals (grey), convalescent individuals (orange), 2-dose (green) and 3-dose (blue) vaccinees. (A) Anti-S IgG, (B) Anti-RBD IgG, (C) Anti-NTD IgG.

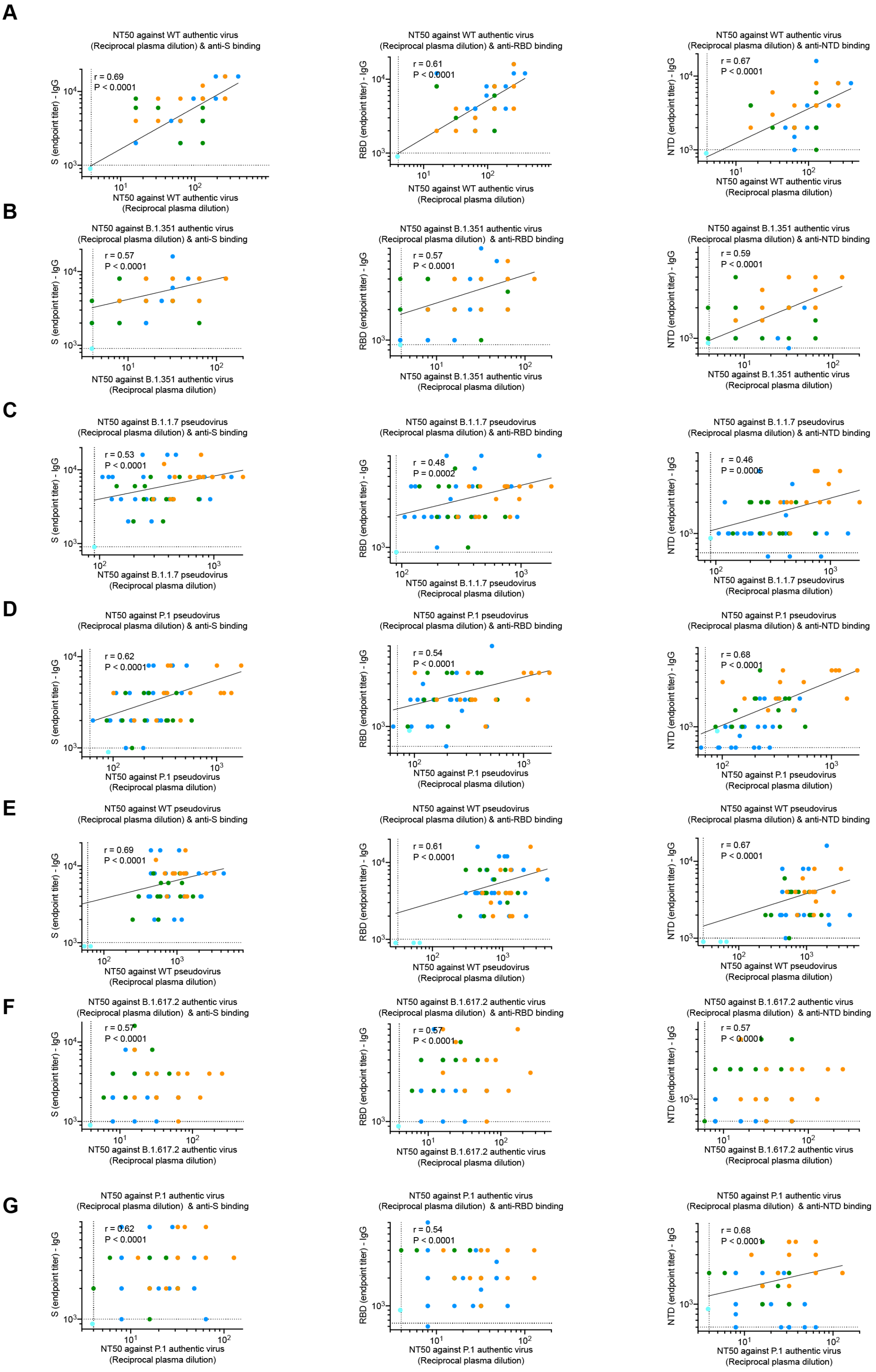

A
NTD50 against WT authentic virus (Reciprocal plasma dilution) & anti-S binding
r = 0.69
P < 0.0001
S (endpoint titer) - IgG
NT50 against WT authentic virus (Reciprocal plasma dilution)
NT50 against WT authentic virus (Reciprocal plasma dilution) & anti-RBD binding
r = 0.61
P < 0.0001
RBD (endpoint titer) - IgG
NT50 against WT authentic virus (Reciprocal plasma dilution) & anti-NTD binding
r = 0.67
P < 0.0001
NTD (endpoint titer) - IgG
B
NT50 against B.1.351 authentic virus (Reciprocal plasma dilution) & anti-S binding
r = 0.57
P < 0.0001
NT50 against B.1.351 authentic virus (Reciprocal plasma dilution) & anti-RBD binding
r = 0.57
P < 0.0001
NT50 against B.1.351 authentic virus (Reciprocal plasma dilution) & anti-NTD binding
r = 0.59
P < 0.0001
NT50 against B.1.351 authentic virus (Reciprocal plasma dilution)
C
NT50 against B.1.1.7 pseudovirus (Reciprocal plasma dilution) & anti-S binding
r = 0.53
P < 0.0001
NT50 against B.1.1.7 pseudovirus (Reciprocal plasma dilution) & anti-RBD binding
r = 0.48
P = 0.0002
NT50 against B.1.1.7 pseudovirus (Reciprocal plasma dilution) & anti-NTD binding
r = 0.46
P = 0.0006
NT50 against B.1.1.7 pseudovirus (Reciprocal plasma dilution)
D
NT50 against P.1 pseudovirus (Reciprocal plasma dilution) & anti-S binding
r = 0.62
P < 0.0001
NT50 against P.1 pseudovirus (Reciprocal plasma dilution) & anti-RBD binding
r = 0.54
P < 0.0001
NT50 against P.1 pseudovirus (Reciprocal plasma dilution) & anti-NTD binding
r = 0.68
P < 0.0001
NT50 against P.1 pseudovirus (Reciprocal plasma dilution)
E
NT50 against WT pseudovirus (Reciprocal plasma dilution) & anti-S binding
r = 0.69
P < 0.0001
NT50 against WT pseudovirus (Reciprocal plasma dilution) & anti-RBD binding
r = 0.61
P < 0.0001
NT50 against WT pseudovirus (Reciprocal plasma dilution) & anti-NTD binding
r = 0.67
P < 0.0001
NT50 against WT pseudovirus (Reciprocal plasma dilution)
F
NT50 against B.1.617.2 authentic virus (Reciprocal plasma dilution) & anti-S binding
r = 0.57
P < 0.0001
NT50 against B.1.617.2 authentic virus (Reciprocal plasma dilution) & anti-RBD binding
r = 0.57
P < 0.0001
NT50 against B.1.617.2 authentic virus (Reciprocal plasma dilution) & anti-NTD binding
r = 0.57
P < 0.0001
NT50 against B.1.617.2 authentic virus (Reciprocal plasma dilution)
G
NT50 against P.1 authentic virus (Reciprocal plasma dilution) & anti-S binding
r = 0.62
P < 0.0001
NT50 against P.1 authentic virus (Reciprocal plasma dilution) & anti-RBD binding
r = 0.54
P < 0.0001
NT50 against P.1 authentic virus (Reciprocal plasma dilution) & anti-NTD binding
r = 0.68
P < 0.0001
NT50 against P.1 authentic virus (Reciprocal plasma dilution)

**Fig. S2 IgG endpoint titer plotted against $NT_{50}$.**

Anti-S (left), anti-RBD (middle), and anti-NTD (right) IgG endpoint titer (y-axis) plotted against $NT_{50}$ (x-axis) for WT authentic virus (A), B.1.351 authentic virus (B), B.1.1.7 pseudovirus (C), P.1 pseudovirus (D), WT pseudovirus (E), B.1.617.2 authentic virus (F), and P.1 authentic virus (G). R- and p-value are label on the top left corner. Data from 3-dose vaccinees, 2-dose vaccinees, convalescent individuals, and control individuals are colored as blue, green, orange and grey, respectively.

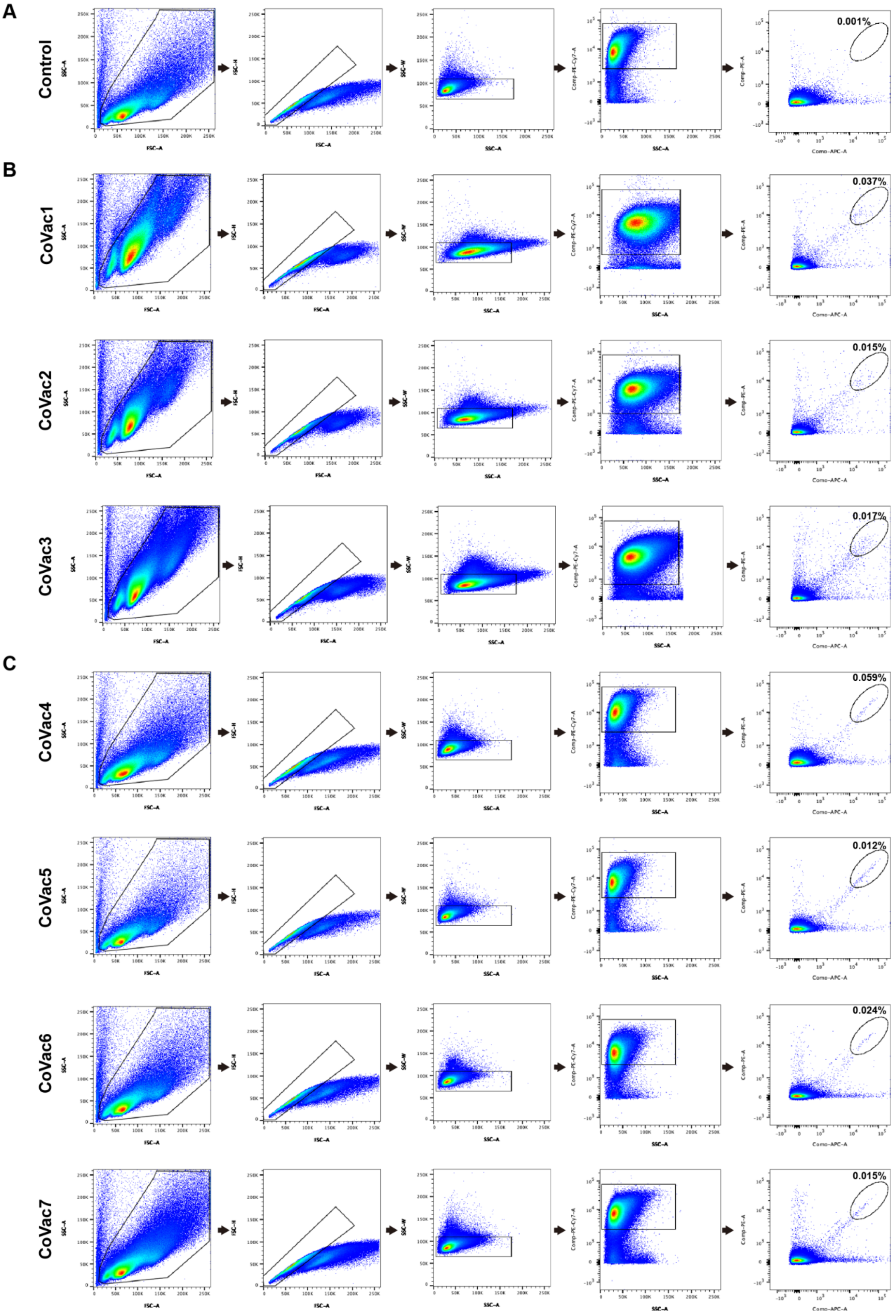

**Fig S3 Gating strategy used for cell sorting and representative data for flow cytometry.**

(A-C) Flow cytometry showing the percentage of S-ECD-double positive B cells from an uninfected unvaccinated control donor (A), three 2-dose vaccinees (B), and four 3-dose vaccinees. Each panel from left to right corresponds to a sequential gating strategy, from live cells (X axis, FSC-A; Y axis, SSA-A in panel 1) and singlets (X axis, FSC-A; Y axis, FSC-H in panel 2; and X axis, SSC-A; Y axis, SSA-W in panel 3) to CD20$^{+}$ B cells (X axis, SSC-A; Y axis, CD20-PE-Cy7 in panel 4) and S-ECD-double positive B cells (X axis, S-ECD-APC; Y axis, S-ECD-PE in panel 5). Sorted cells were S-ECD-APC$^{+}$ and S-ECD-PE$^{+}$, with their frequencies labeled in the top right corner in panel 5. FSC, forward scatter; SSC, side scatter (-A, area; -H, height; -W, width).

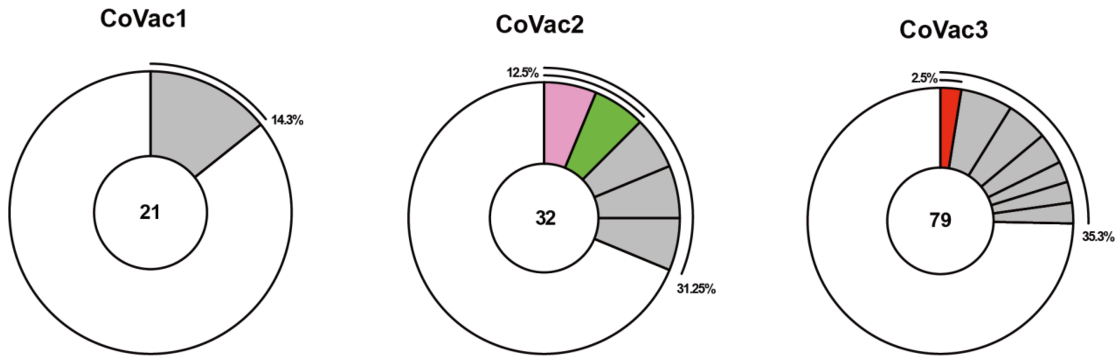

**Fig. S4 Antibody pie charts for the three 2-dose vaccinees.**

There are 21, 32 and 79 sequenced antibodies with naturally paired Ig heavy and light chains, respectively for the three 2-dose vaccinees. Antibodies with the same IGHV/IGLV genes and closely related CDR3 sequences were grouped together and represented as a slice (grey). Antibody singlets are in one big slice (white). Colored slices reveal expanded antibody clones sharing the same *IGHV* and *IGLV* genes with the four 3-dose vaccinees (see Fig. 2).

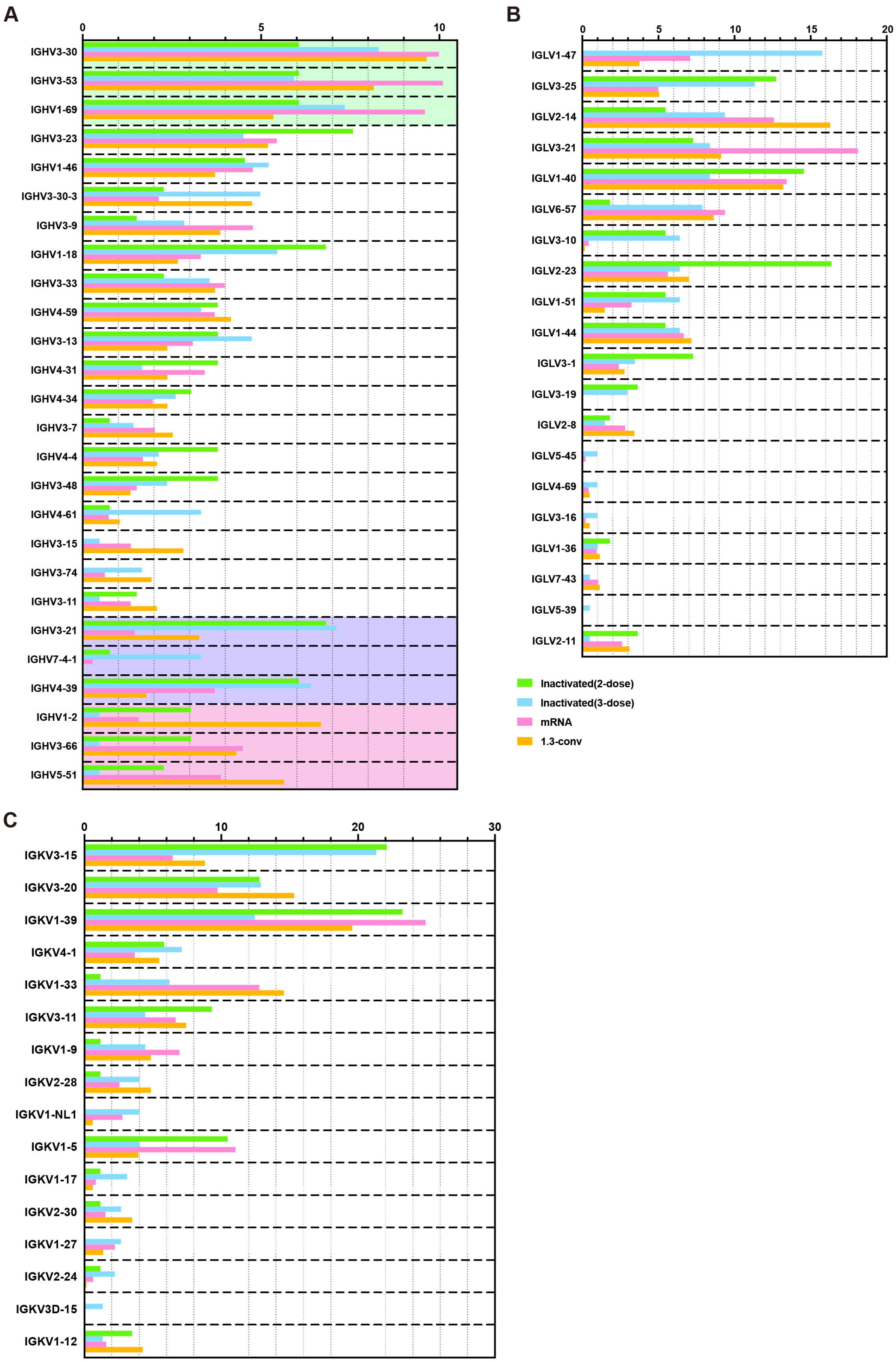
A
B
C
IGHV3-30
IGHV3-53
IGHV1-69
IGHV3-23
IGHV1-46
IGHV3-30-3
IGHV3-9
IGHV1-18
IGHV3-33
IGHV4-59
IGHV3-13
IGHV4-31
IGHV4-34
IGHV3-7
IGHV4-4
IGHV3-48
IGHV4-61
IGHV3-15
IGHV3-74
IGHV3-11
IGHV3-21
IGHV7-4-1
IGHV4-39
IGHV1-2
IGHV3-66
IGHV5-51
IGLV1-47
IGLV3-25
IGLV2-14
IGLV3-21
IGLV1-40
IGLV6-57
IGLV3-10
IGLV2-23
IGLV1-51
IGLV1-44
IGLV3-1
IGLV3-19
IGLV2-8
IGLV5-45
IGLV4-69
IGLV3-16
IGLV1-36
IGLV7-43
IGLV5-39
IGLV2-11
Inactivated(2-dose)
Inactivated(3-dose)
mRNA
1.3-conv
IGKV3-15
IGKV3-20
IGKV1-39
IGKV4-1
IGKV1-33
IGKV3-11
IGKV1-9
IGKV2-28
IGKV1-NL1
IGKV1-5
IGKV1-17
IGKV2-30
IGKV1-27
IGKV2-24
IGKV3D-15
IGKV1-12

**Fig. S5 Frequency distributions of human *V* genes.**

(A-C) Graphs show relative percentage (X axis) of human *IGHV* (A), *IGLV* (B) and *IGKV* (C) genes in data from convalescent (1.3 month) individuals (orange), 2-doses inactivated vaccinees (green), 3-dose inactivated vaccinees (blue), and mRNA vaccinees (pink). The top 3 popular *IGVH* genes for 3-dose inactivated, 2-dose inactivated, and mRNA vaccinees are highlighted with purple, green, and pink box, respectively.

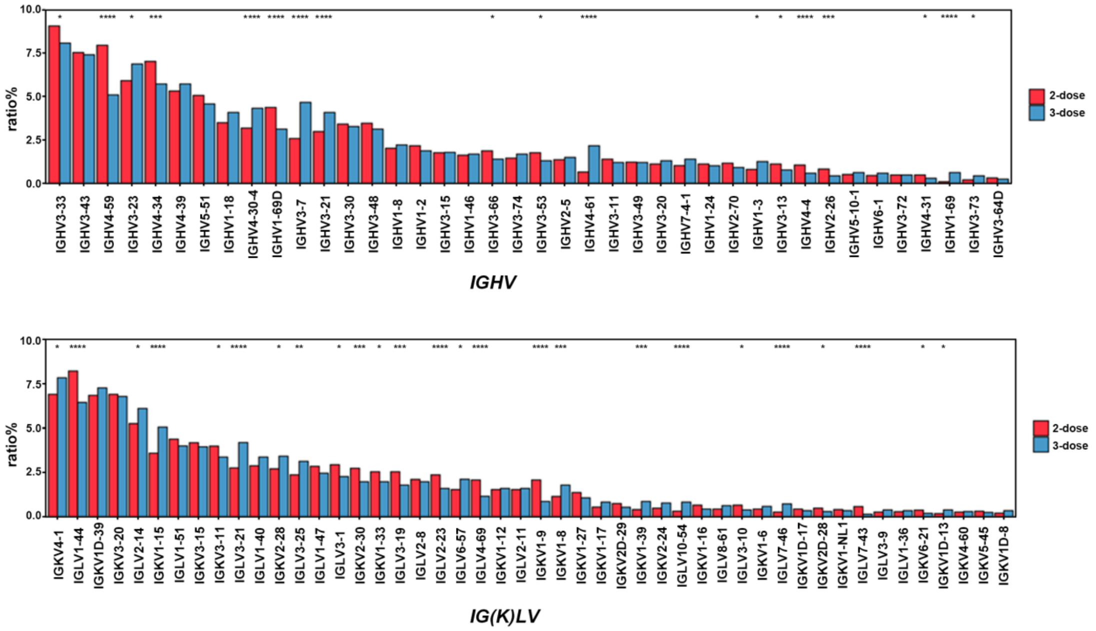

**Fig. S6 Large-scale, single-cell sequencing datasets generated from two cohorts consisting of 2-dose and 3-dose vaccinees.**

Graph shows relative abundance of human *IGHV* and *IGK(L)V* genes of two cohorts of 2-dose (red, n=10) and 3-dose vaccinees (blue, n=10). Two-sided binomial tests with unequal variance were used to compare the frequency distributions. *P<0.05, **P<0.01, ***P<0.001, ****P<0.0001.

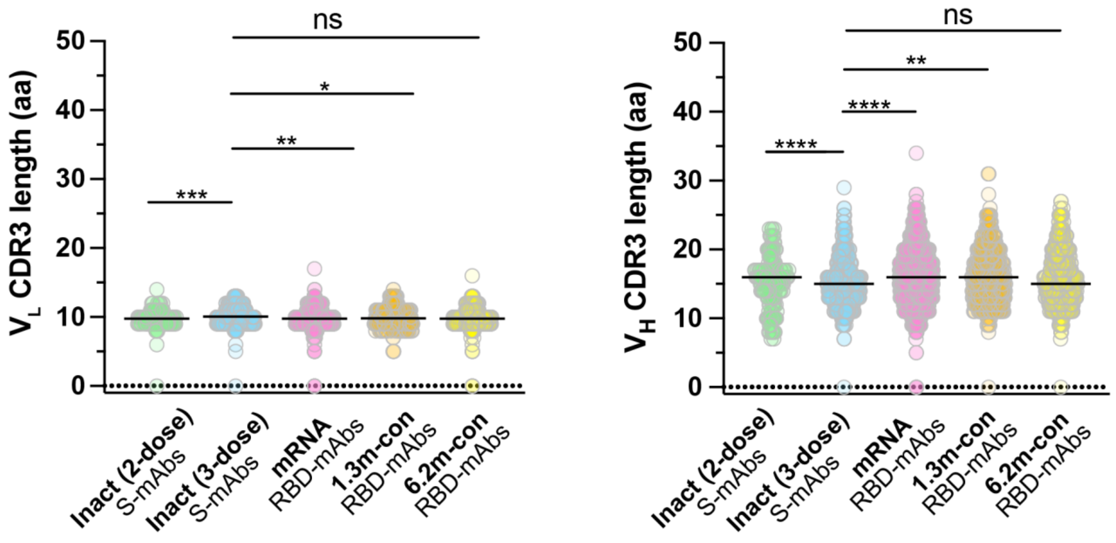

**Fig. S7 CDR3 length distribution.**

The amino acid length of CDR3 of light chain and heavy chain V genes from 2-dose (green) and 3-dose (blue) inactivated vaccine vaccinees, mRNA vaccine (pink) vaccinated, 1.3-month (orange) and 6.2-month (yellow) convalescent individuals.

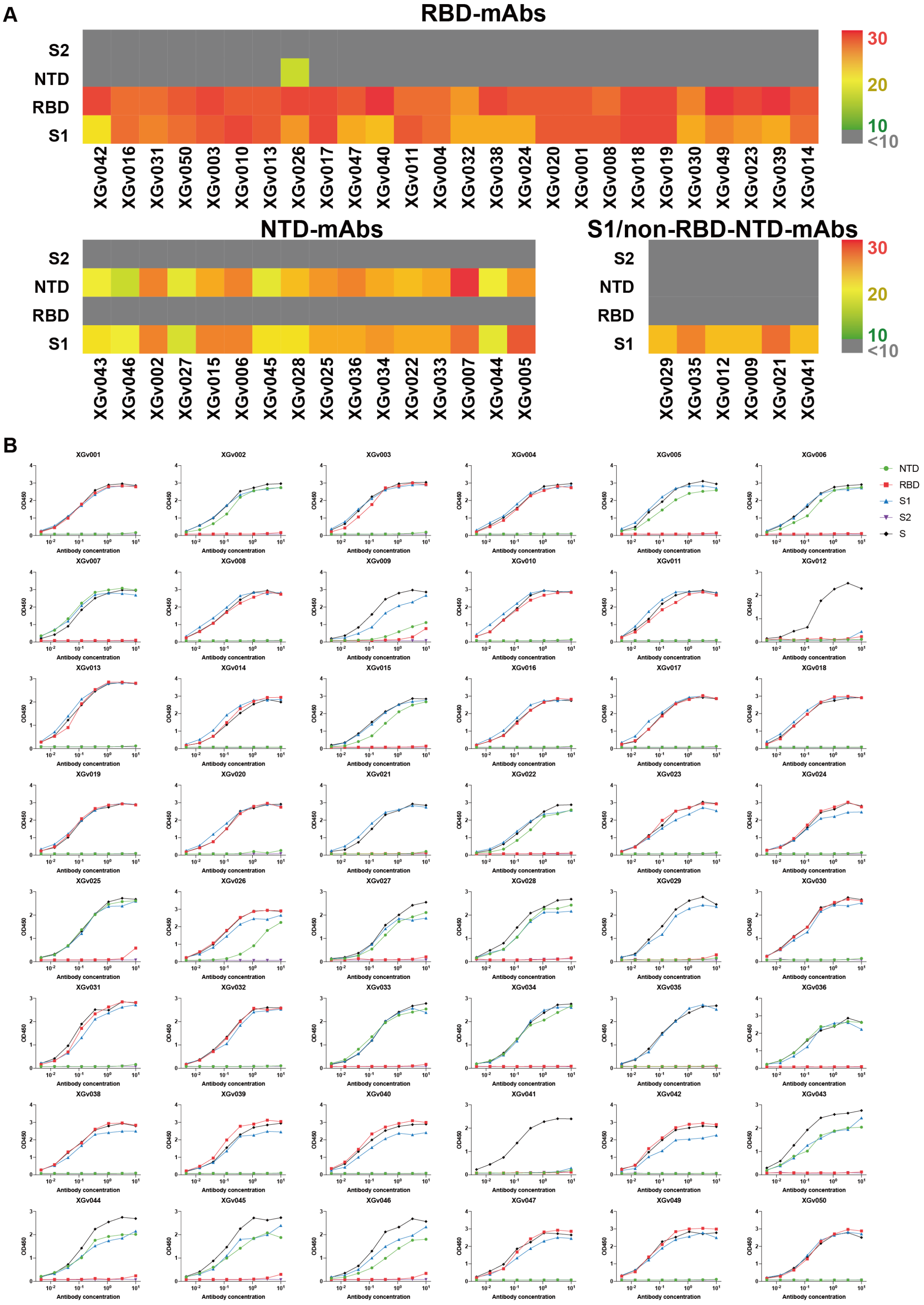
A
RBD-mAbs
S2
NTD
RBD
S1
30
20
10
<10
XGv042
XGv016
XGv031
XGv050
XGv003
XGv010
XGv013
XGv026
XGv017
XGv047
XGv040
XGv011
XGv004
XGv032
XGv038
XGv024
XGv020
XGv001
XGv008
XGv018
XGv019
XGv030
XGv049
XGv023
XGv039
XGv014
NTD-mAbs
XGv043
XGv046
XGv002
XGv027
XGv015
XGv006
XGv045
XGv028
XGv025
XGv036
XGv034
XGv022
XGv033
XGv007
XGv044
XGv005
S1/non-RBD-NTD-mAbs
XGv029
XGv035
XGv012
XGv009
XGv021
XGv041
B
NTD
RBD
S1
S2
S
OD450
Antibody concentration
XGv001
XGv002
XGv003
XGv004
XGv005
XGv006
XGv007
XGv008
XGv009
XGv010
XGv011
XGv012
XGv013
XGv014
XGv015
XGv016
XGv017
XGv018
XGv019
XGv020
XGv021
XGv022
XGv023
XGv024
XGv025
XGv026
XGv027
XGv028
XGv029
XGv030
XGv031
XGv032
XGv033
XGv034
XGv035
XGv036
XGv038
XGv039
XGv040
XGv041
XGv042
XGv043
XGv044
XGv045
XGv046
XGv047
XGv049
XGv050

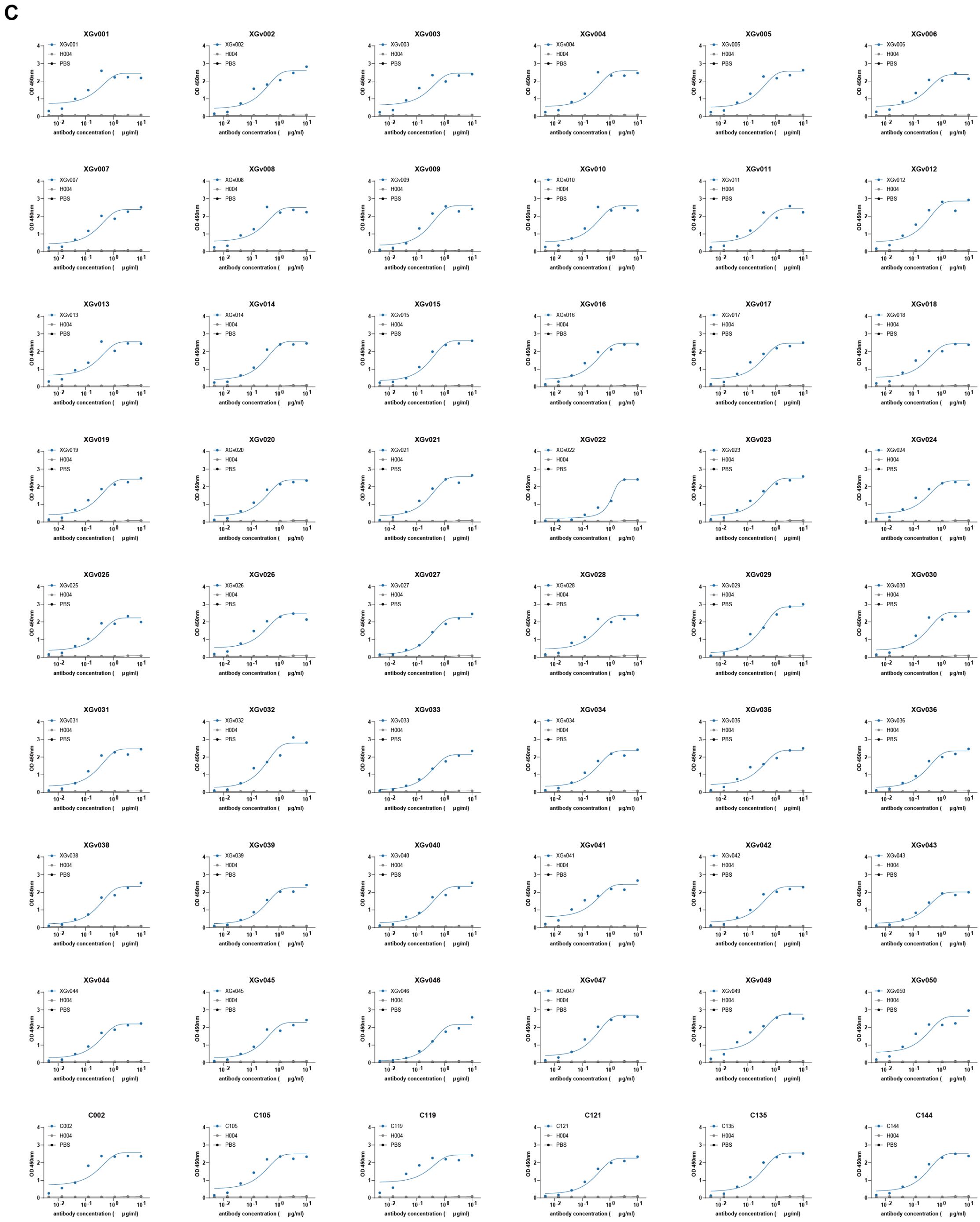

**Fig. S8 Antibody binding domain scanning and $EC_{50}$ calculation by ELISA**.

(A) Heatmap representation of AUC values of the 48 XGv mAbs by ELISA against the subdomain S2, NTD, RBD or S1 of SARS-CoV-2 S, respectively was shown with a color bar indicated on the right.

(B) Curves corresponding to each value in the heatmap were generated based on all the ELISA values at different concentrations.

(C) $EC_{50}$ values of the 48 XGv mAbs and reference mAbs of C002, C105, C119, C121, C135 and C144 were calculated from the curves fitted from ELISA results; then, each $EC_{50}$ of the XGv mAbs was normalized against the $EC_{50}$ values of the reference mAbs obtained previously to compare with those of antibodies reported from Michel's group.

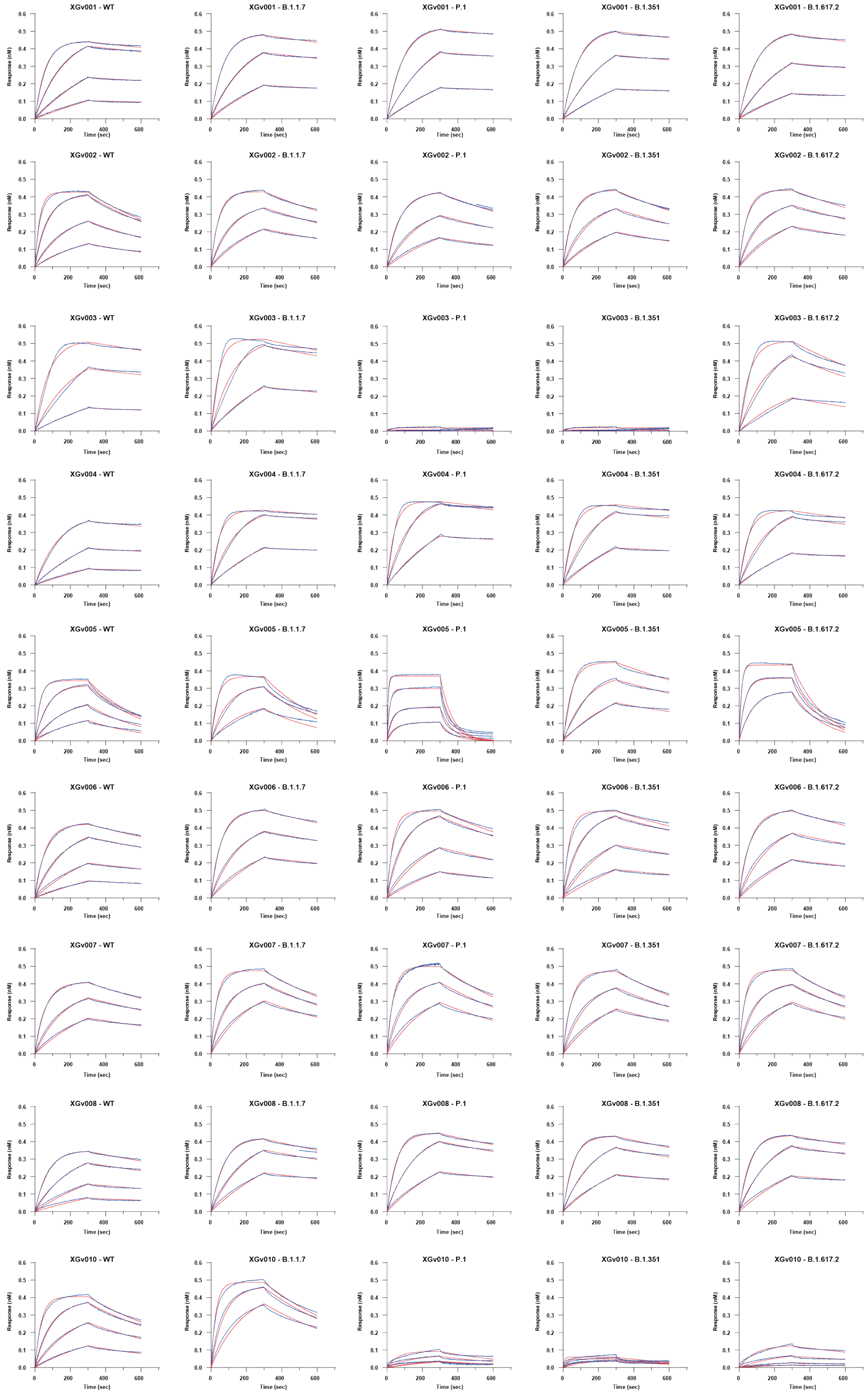
XGv001 - WT
XGv001 - B.1.1.7
XGv001 - P.1
XGv001 - B.1.351
XGv001 - B.1.617.2
XGv002 - WT
XGv002 - B.1.1.7
XGv002 - P.1
XGv002 - B.1.351
XGv002 - B.1.617.2
XGv003 - WT
XGv003 - B.1.1.7
XGv003 - P.1
XGv003 - B.1.351
XGv003 - B.1.617.2
XGv004 - WT
XGv004 - B.1.1.7
XGv004 - P.1
XGv004 - B.1.351
XGv004 - B.1.617.2
XGv005 - WT
XGv005 - B.1.1.7
XGv005 - P.1
XGv005 - B.1.351
XGv005 - B.1.617.2
XGv006 - WT
XGv006 - B.1.1.7
XGv006 - P.1
XGv006 - B.1.351
XGv006 - B.1.617.2
XGv007 - WT
XGv007 - B.1.1.7
XGv007 - P.1
XGv007 - B.1.351
XGv007 - B.1.617.2
XGv008 - WT
XGv008 - B.1.1.7
XGv008 - P.1
XGv008 - B.1.351
XGv008 - B.1.617.2
XGv010 - WT
XGv010 - B.1.1.7
XGv010 - P.1
XGv010 - B.1.351
XGv010 - B.1.617.2
Response (nM)
Time (sec)
0.0
0.1
0.2
0.3
0.4
0.5
0.6
0
200
400
600

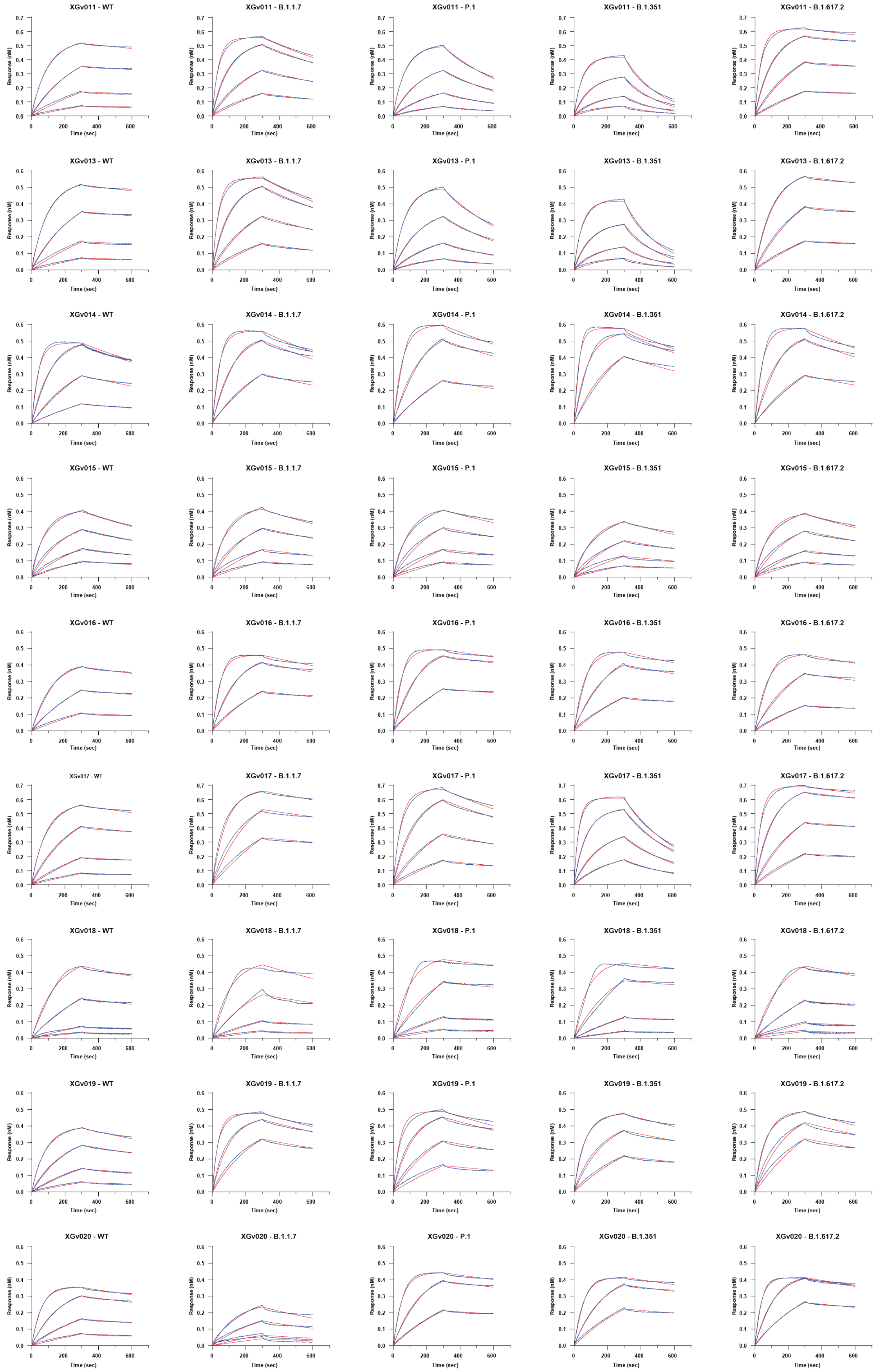
XGv011 - WT
XGv011 - B.1.1.7
XGv011 - P.1
XGv011 - B.1.351
XGv011 - B.1.617.2
XGv013 - WT
XGv013 - B.1.1.7
XGv013 - P.1
XGv013 - B.1.351
XGv013 - B.1.617.2
XGv014 - WT
XGv014 - B.1.1.7
XGv014 - P.1
XGv014 - B.1.351
XGv014 - B.1.617.2
XGv015 - WT
XGv015 - B.1.1.7
XGv015 - P.1
XGv015 - B.1.351
XGv015 - B.1.617.2
XGv016 - WT
XGv016 - B.1.1.7
XGv016 - P.1
XGv016 - B.1.351
XGv016 - B.1.617.2
XGv017 - WT
XGv017 - B.1.1.7
XGv017 - P.1
XGv017 - B.1.351
XGv017 - B.1.617.2
XGv018 - WT
XGv018 - B.1.1.7
XGv018 - P.1
XGv018 - B.1.351
XGv018 - B.1.617.2
XGv019 - WT
XGv019 - B.1.1.7
XGv019 - P.1
XGv019 - B.1.351
XGv019 - B.1.617.2
XGv020 - WT
XGv020 - B.1.1.7
XGv020 - P.1
XGv020 - B.1.351
XGv020 - B.1.617.2
Response (nM)
Time (sec)

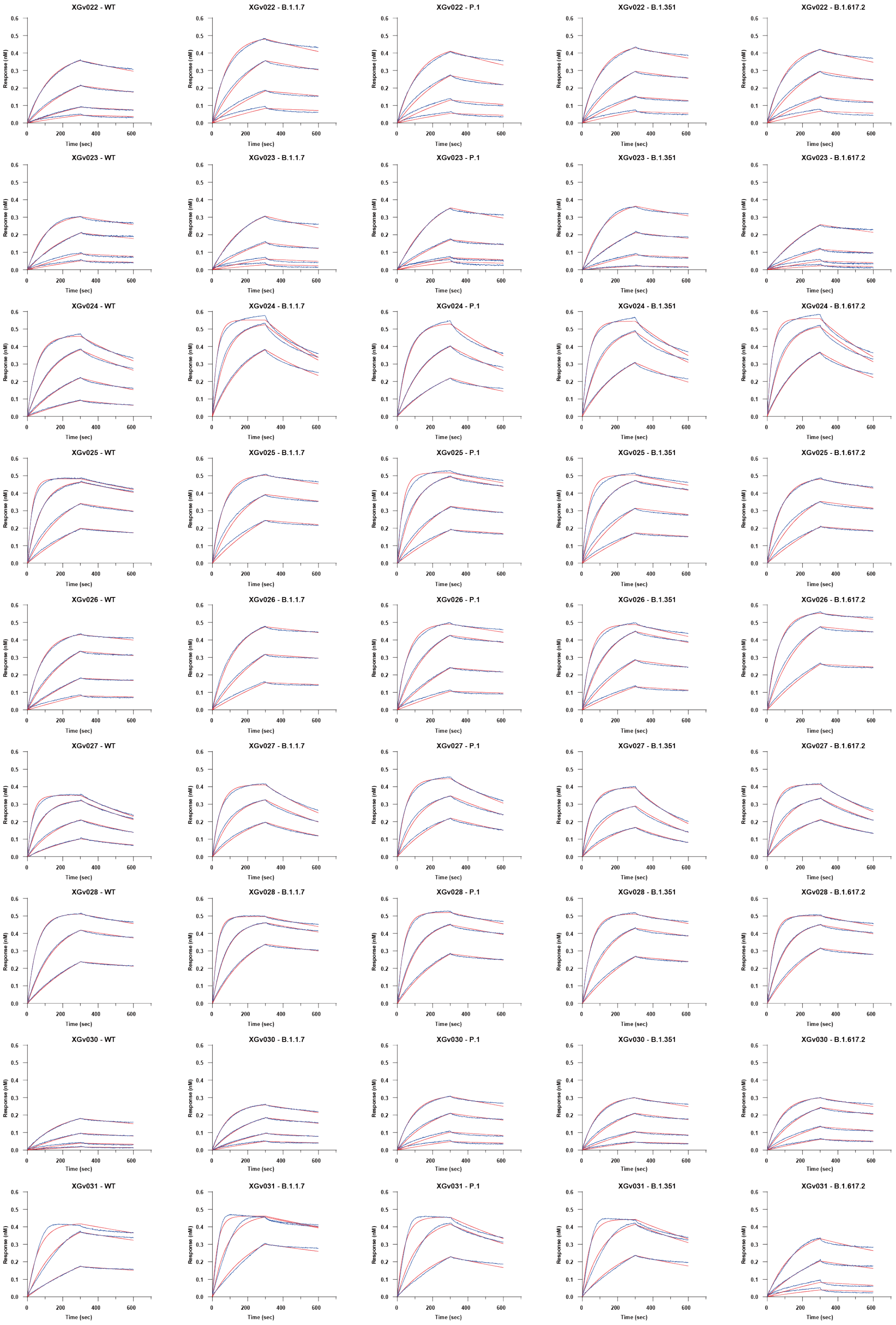
XGv022 - WT
XGv022 - B.1.1.7
XGv022 - P.1
XGv022 - B.1.351
XGv022 - B.1.617.2
XGv023 - WT
XGv023 - B.1.1.7
XGv023 - P.1
XGv023 - B.1.351
XGv023 - B.1.617.2
XGv024 - WT
XGv024 - B.1.1.7
XGv024 - P.1
XGv024 - B.1.351
XGv024 - B.1.617.2
XGv025 - WT
XGv025 - B.1.1.7
XGv025 - P.1
XGv025 - B.1.351
XGv025 - B.1.617.2
XGv026 - WT
XGv026 - B.1.1.7
XGv026 - P.1
XGv026 - B.1.351
XGv026 - B.1.617.2
XGv027 - WT
XGv027 - B.1.1.7
XGv027 - P.1
XGv027 - B.1.351
XGv027 - B.1.617.2
XGv028 - WT
XGv028 - B.1.1.7
XGv028 - P.1
XGv028 - B.1.351
XGv028 - B.1.617.2
XGv030 - WT
XGv030 - B.1.1.7
XGv030 - P.1
XGv030 - B.1.351
XGv030 - B.1.617.2
XGv031 - WT
XGv031 - B.1.1.7
XGv031 - P.1
XGv031 - B.1.351
XGv031 - B.1.617.2
Response (nM)
Time (sec)

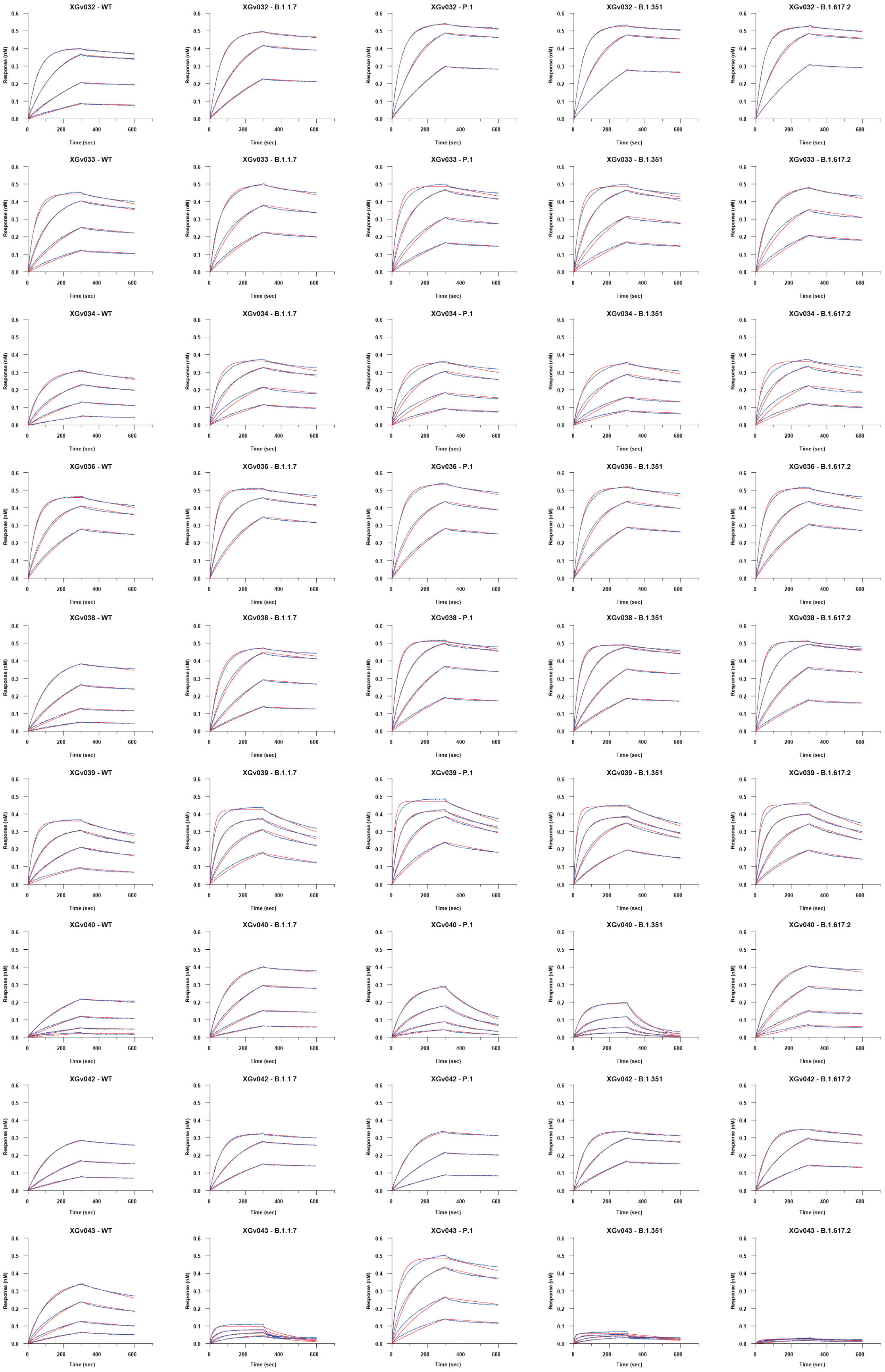
XGv032 - WT
XGv032 - B.1.1.7
XGv032 - P.1
XGv032 - B.1.351
XGv032 - B.1.617.2
XGv033 - WT
XGv033 - B.1.1.7
XGv033 - P.1
XGv033 - B.1.351
XGv033 - B.1.617.2
XGv034 - WT
XGv034 - B.1.1.7
XGv034 - P.1
XGv034 - B.1.351
XGv034 - B.1.617.2
XGv036 - WT
XGv036 - B.1.1.7
XGv036 - P.1
XGv036 - B.1.351
XGv036 - B.1.617.2
XGv038 - WT
XGv038 - B.1.1.7
XGv038 - P.1
XGv038 - B.1.351
XGv038 - B.1.617.2
XGv039 - WT
XGv039 - B.1.1.7
XGv039 - P.1
XGv039 - B.1.351
XGv039 - B.1.617.2
XGv040 - WT
XGv040 - B.1.1.7
XGv040 - P.1
XGv040 - B.1.351
XGv040 - B.1.617.2
XGv042 - WT
XGv042 - B.1.1.7
XGv042 - P.1
XGv042 - B.1.351
XGv042 - B.1.617.2
XGv043 - WT
XGv043 - B.1.1.7
XGv043 - P.1
XGv043 - B.1.351
XGv043 - B.1.617.2
Response (nM)
Time (sec)

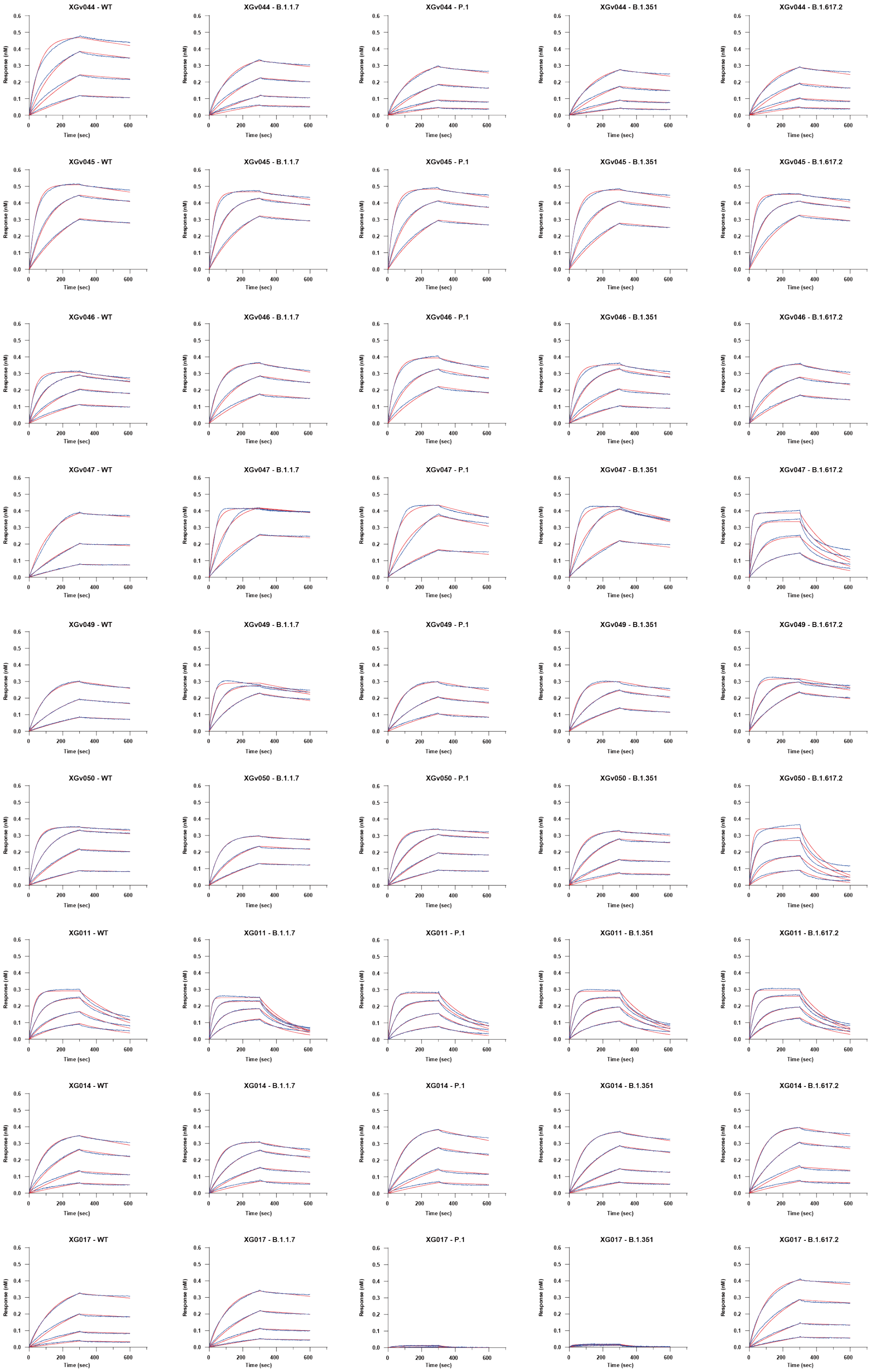

XGv044 - WT
XGv044 - B.1.1.7
XGv044 - P.1
XGv044 - B.1.351
XGv044 - B.1.617.2
XGv045 - WT
XGv045 - B.1.1.7
XGv045 - P.1
XGv045 - B.1.351
XGv045 - B.1.617.2
XGv046 - WT
XGv046 - B.1.1.7
XGv046 - P.1
XGv046 - B.1.351
XGv046 - B.1.617.2
XGv047 - WT
XGv047 - B.1.1.7
XGv047 - P.1
XGv047 - B.1.351
XGv047 - B.1.617.2
XGv049 - WT
XGv049 - B.1.1.7
XGv049 - P.1
XGv049 - B.1.351
XGv049 - B.1.617.2
XGv050 - WT
XGv050 - B.1.1.7
XGv050 - P.1
XGv050 - B.1.351
XGv050 - B.1.617.2
XG011 - WT
XG011 - B.1.1.7
XG011 - P.1
XG011 - B.1.351
XG011 - B.1.617.2
XG014 - WT
XG014 - B.1.1.7
XG014 - P.1
XG014 - B.1.351
XG014 - B.1.617.2
XG017 - WT
XG017 - B.1.1.7
XG017 - P.1
XG017 - B.1.351
XG017 - B.1.617.2
Response (nM)
Time (sec)

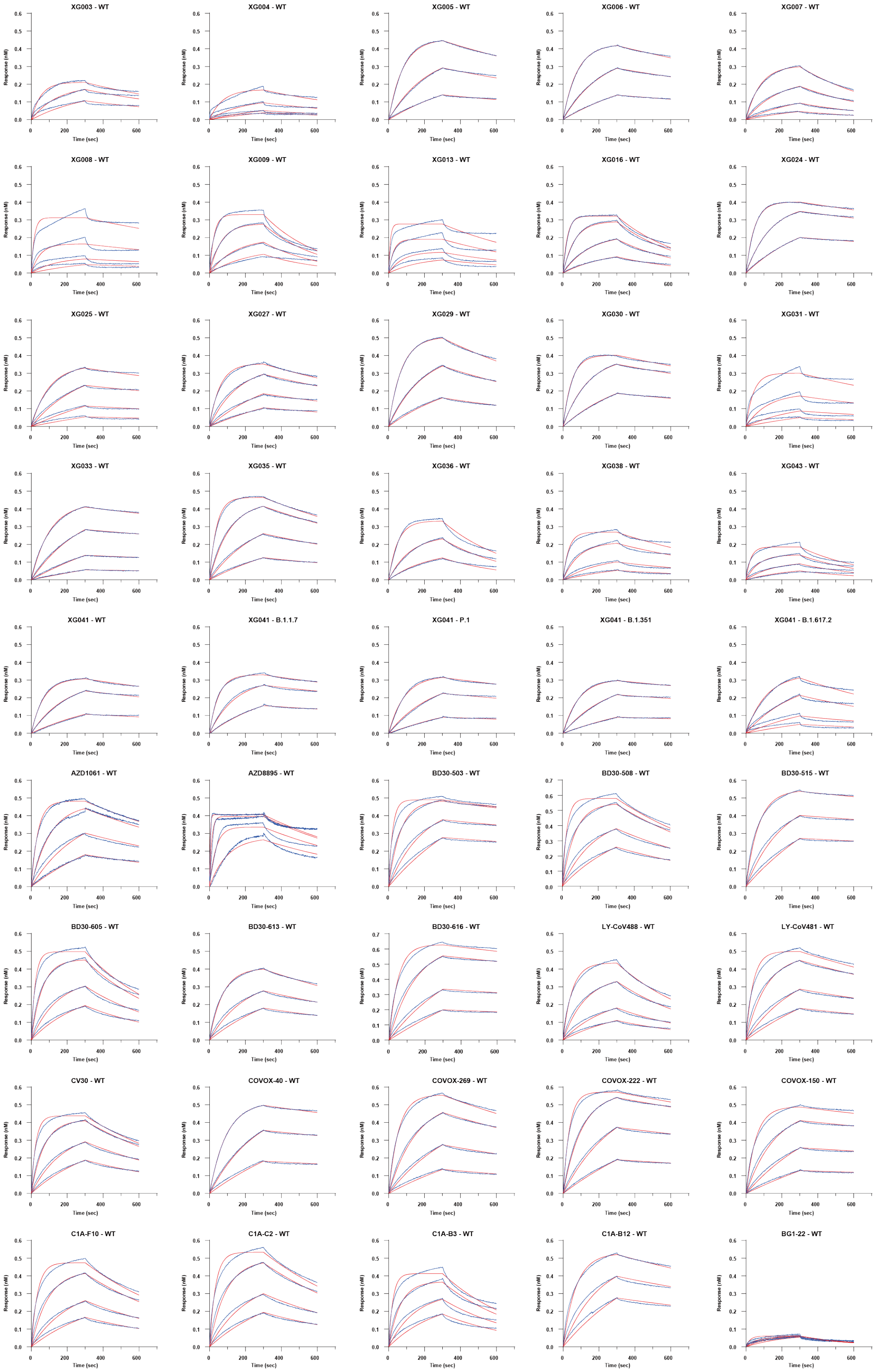

XG003 - WT
XG004 - WT
XG005 - WT
XG006 - WT
XG007 - WT
XG008 - WT
XG009 - WT
XG013 - WT
XG016 - WT
XG024 - WT
XG025 - WT
XG027 - WT
XG029 - WT
XG030 - WT
XG031 - WT
XG033 - WT
XG035 - WT
XG036 - WT
XG038 - WT
XG043 - WT
XG041 - WT
XG041 - B.1.1.7
XG041 - P.1
XG041 - B.1.351
XG041 - B.1.617.2
AZD1061 - WT
AZD8895 - WT
BD30-503 - WT
BD30-508 - WT
BD30-515 - WT
BD30-605 - WT
BD30-613 - WT
BD30-616 - WT
LY-CoV488 - WT
LY-CoV481 - WT
CV30 - WT
COVOX-40 - WT
COVOX-269 - WT
COVOX-222 - WT
COVOX-150 - WT
C1A-F10 - WT
C1A-C2 - WT
C1A-B3 - WT
C1A-B12 - WT
BG1-22 - WT
Response (nM)
Time (sec)

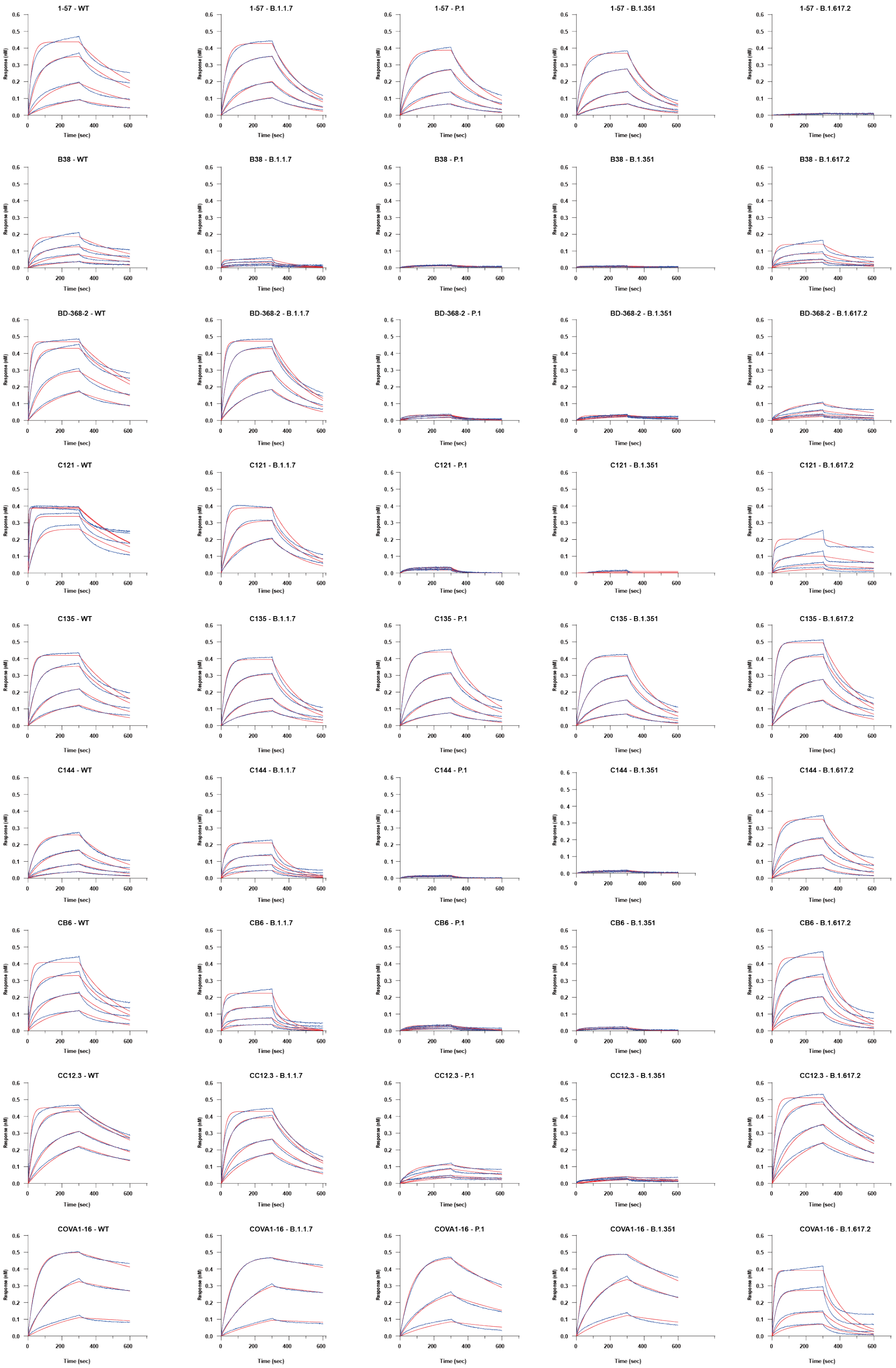
1-57 - WT
1-57 - B.1.1.7
1-57 - P.1
1-57 - B.1.351
1-57 - B.1.617.2
B38 - WT
B38 - B.1.1.7
B38 - P.1
B38 - B.1.351
B38 - B.1.617.2
BD-368-2 - WT
BD-368-2 - B.1.1.7
BD-368-2 - P.1
BD-368-2 - B.1.351
BD-368-2 - B.1.617.2
C121 - WT
C121 - B.1.1.7
C121 - P.1
C121 - B.1.351
C121 - B.1.617.2
C135 - WT
C135 - B.1.1.7
C135 - P.1
C135 - B.1.351
C135 - B.1.617.2
C144 - WT
C144 - B.1.1.7
C144 - P.1
C144 - B.1.351
C144 - B.1.617.2
CB6 - WT
CB6 - B.1.1.7
CB6 - P.1
CB6 - B.1.351
CB6 - B.1.617.2
CC12.3 - WT
CC12.3 - B.1.1.7
CC12.3 - P.1
CC12.3 - B.1.351
CC12.3 - B.1.617.2
COVA1-16 - WT
COVA1-16 - B.1.1.7
COVA1-16 - P.1
COVA1-16 - B.1.351
COVA1-16 - B.1.617.2
Response (nM)
Time (sec)

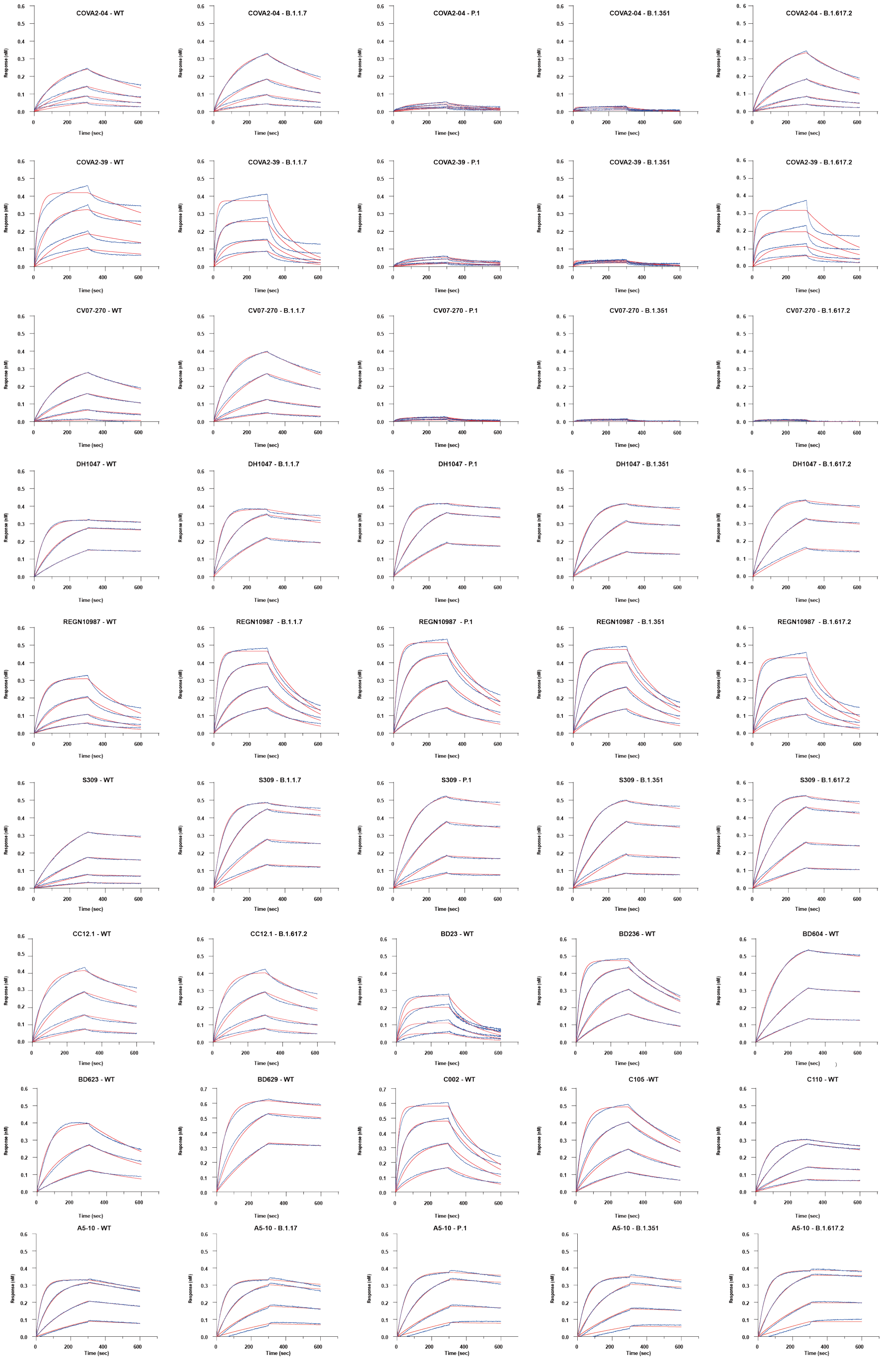

COVA2-04 - WT
COVA2-04 - B.1.1.7
COVA2-04 - P.1
COVA2-04 - B.1.351
COVA2-04 - B.1.617.2
COVA2-39 - WT
COVA2-39 - B.1.1.7
COVA2-39 - P.1
COVA2-39 - B.1.351
COVA2-39 - B.1.617.2
CV07-270 - WT
CV07-270 - B.1.1.7
CV07-270 - P.1
CV07-270 - B.1.351
CV07-270 - B.1.617.2
DH1047 - WT
DH1047 - B.1.1.7
DH1047 - P.1
DH1047 - B.1.351
DH1047 - B.1.617.2
REGN10987 - WT
REGN10987 - B.1.1.7
REGN10987 - P.1
REGN10987 - B.1.351
REGN10987 - B.1.617.2
S309 - WT
S309 - B.1.1.7
S309 - P.1
S309 - B.1.351
S309 - B.1.617.2
CC12.1 - WT
CC12.1 - B.1.617.2
BD23 - WT
BD236 - WT
BD604 - WT
BD623 - WT
BD629 - WT
C002 - WT
C105 -WT
C110 - WT
A5-10 - WT
A5-10 - B.1.17
A5-10 - P.1
A5-10 - B.1.351
A5-10 - B.1.617.2
Response (nM)
Time (sec)

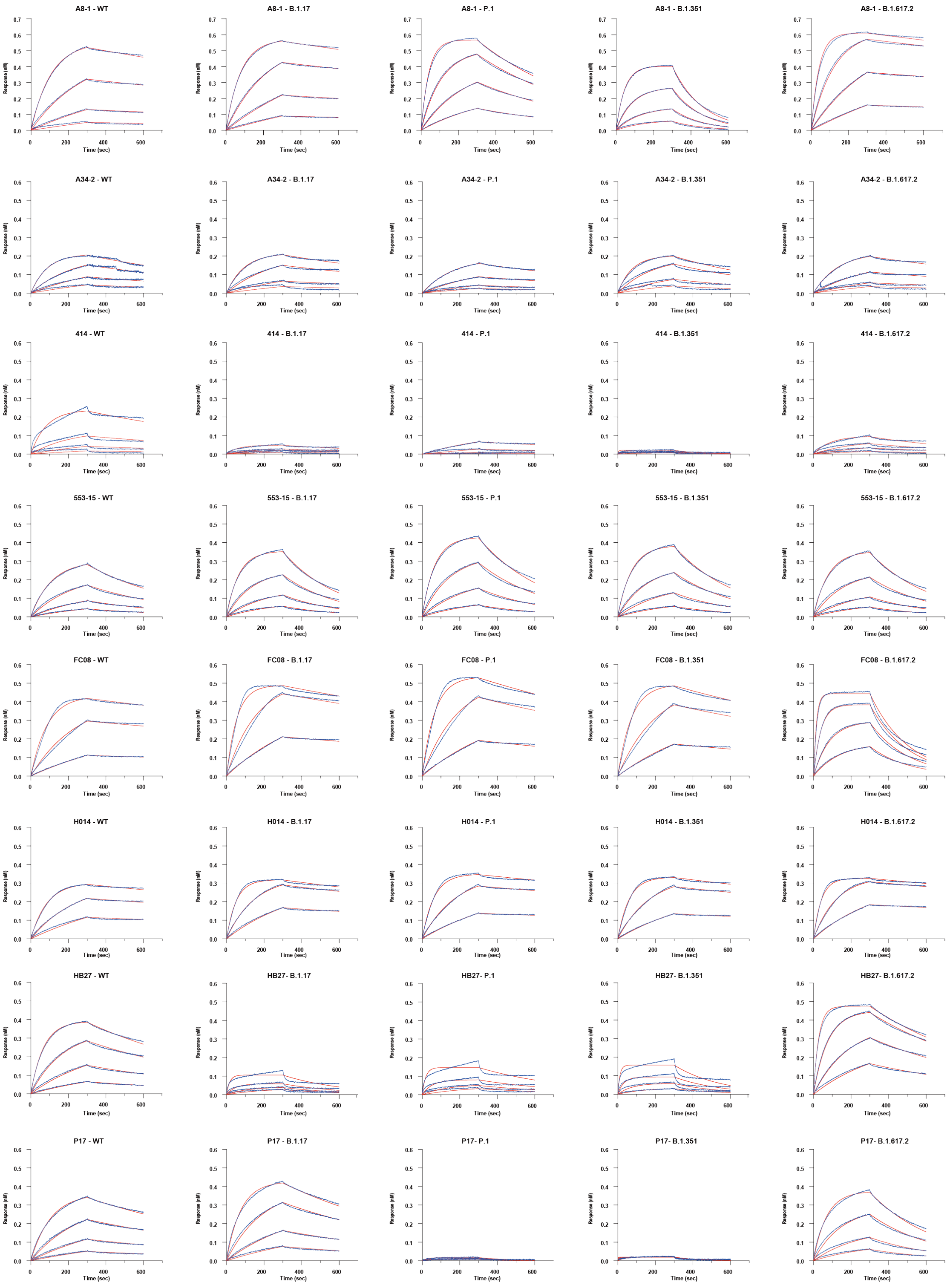
A8-1 - WT
A8-1 - B.1.17
A8-1 - P.1
A8-1 - B.1.351
A8-1 - B.1.617.2
A34-2 - WT
A34-2 - B.1.17
A34-2 - P.1
A34-2 - B.1.351
A34-2 - B.1.617.2
414 - WT
414 - B.1.17
414 - P.1
414 - B.1.351
414 - B.1.617.2
553-15 - WT
553-15 - B.1.17
553-15 - P.1
553-15 - B.1.351
553-15 - B.1.617.2
FC08 - WT
FC08 - B.1.17
FC08 - P.1
FC08 - B.1.351
FC08 - B.1.617.2
H014 - WT
H014 - B.1.17
H014 - P.1
H014 - B.1.351
H014 - B.1.617.2
HB27 - WT
HB27- B.1.17
HB27- P.1
HB27- B.1.351
HB27- B.1.617.2
P17 - WT
P17 - B.1.17
P17- P.1
P17- B.1.351
P17- B.1.617.2
Response (nM)
Time (sec)

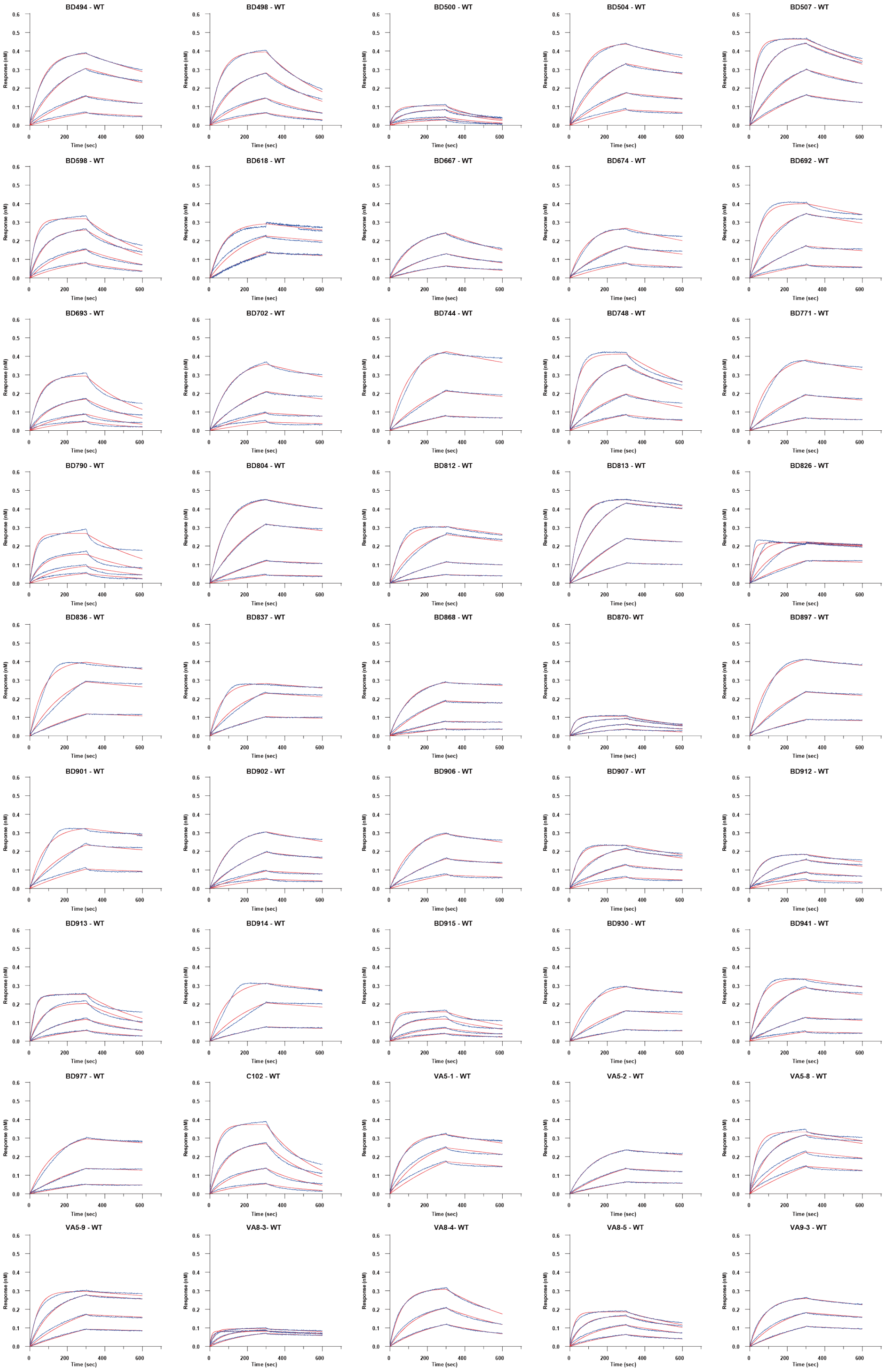
BD494 - WT
BD498 - WT
BD500 - WT
BD504 - WT
BD507 - WT
BD598 - WT
BD618 - WT
BD667 - WT
BD674 - WT
BD692 - WT
BD693 - WT
BD702 - WT
BD744 - WT
BD748 - WT
BD771 - WT
BD790 - WT
BD804 - WT
BD812 - WT
BD813 - WT
BD826 - WT
BD836 - WT
BD837 - WT
BD868 - WT
BD870- WT
BD897 - WT
BD901 - WT
BD902 - WT
BD906 - WT
BD907 - WT
BD912 - WT
BD913 - WT
BD914 - WT
BD915 - WT
BD930 - WT
BD941 - WT
BD977 - WT
C102 - WT
VA5-1 - WT
VA5-2 - WT
VA5-8 - WT
VA5-9 - WT
VA8-3- WT
VA8-4- WT
VA8-5 - WT
VA9-3 - WT
Response (nM)
Time (sec)

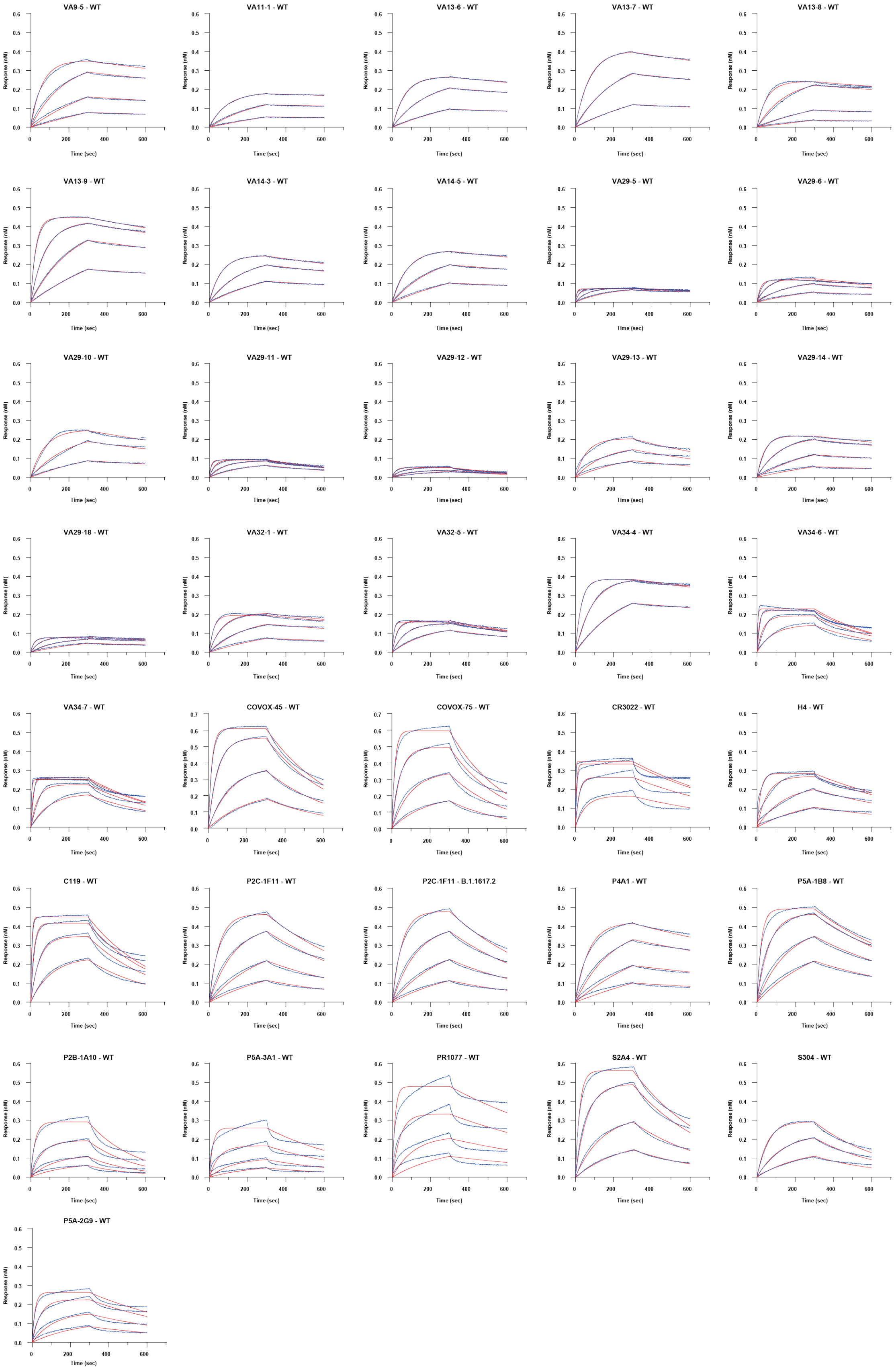

VA9-5 - WT
VA11-1 - WT
VA13-6 - WT
VA13-7 - WT
VA13-8 - WT
VA13-9 - WT
VA14-3 - WT
VA14-5 - WT
VA29-5 - WT
VA29-6 - WT
VA29-10 - WT
VA29-11 - WT
VA29-12 - WT
VA29-13 - WT
VA29-14 - WT
VA29-18 - WT
VA32-1 - WT
VA32-5 - WT
VA34-4 - WT
VA34-6 - WT
VA34-7 - WT
COVOX-45 - WT
COVOX-75 - WT
CR3022 - WT
H4 - WT
C119 - WT
P2C-1F11 - WT
P2C-1F11 - B.1.1617.2
P4A1 - WT
P5A-1B8 - WT
P2B-1A10 - WT
P5A-3A1 - WT
PR1077 - WT
S2A4 - WT
S304 - WT
P5A-2G9 - WT
Response (nM)
Time (sec)

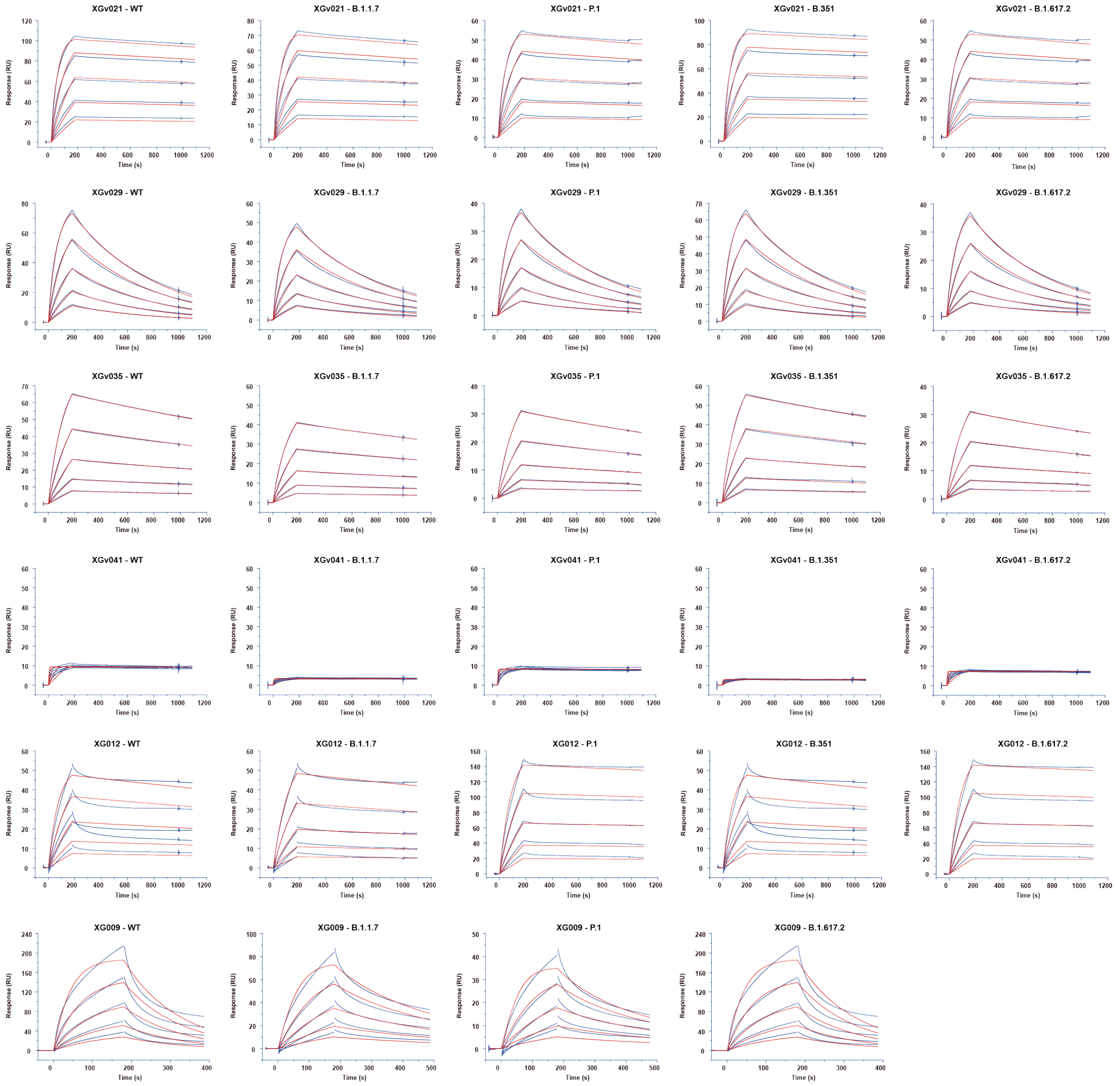

**Fig. S9 Sensorgrams for binding of all antibodies bound to SARS-CoV-2 at sub-nM levels.**

Binding kinetics of all antibodies against SARS-CoV-2 over variants were measured by biolayer interferometry (BLI). Y-axis represents the response. Blue solid lines represent the response curves and red solid lines represent the best fitting curve. The $K_D$ of the fitting is shown at Table S2.

**A**

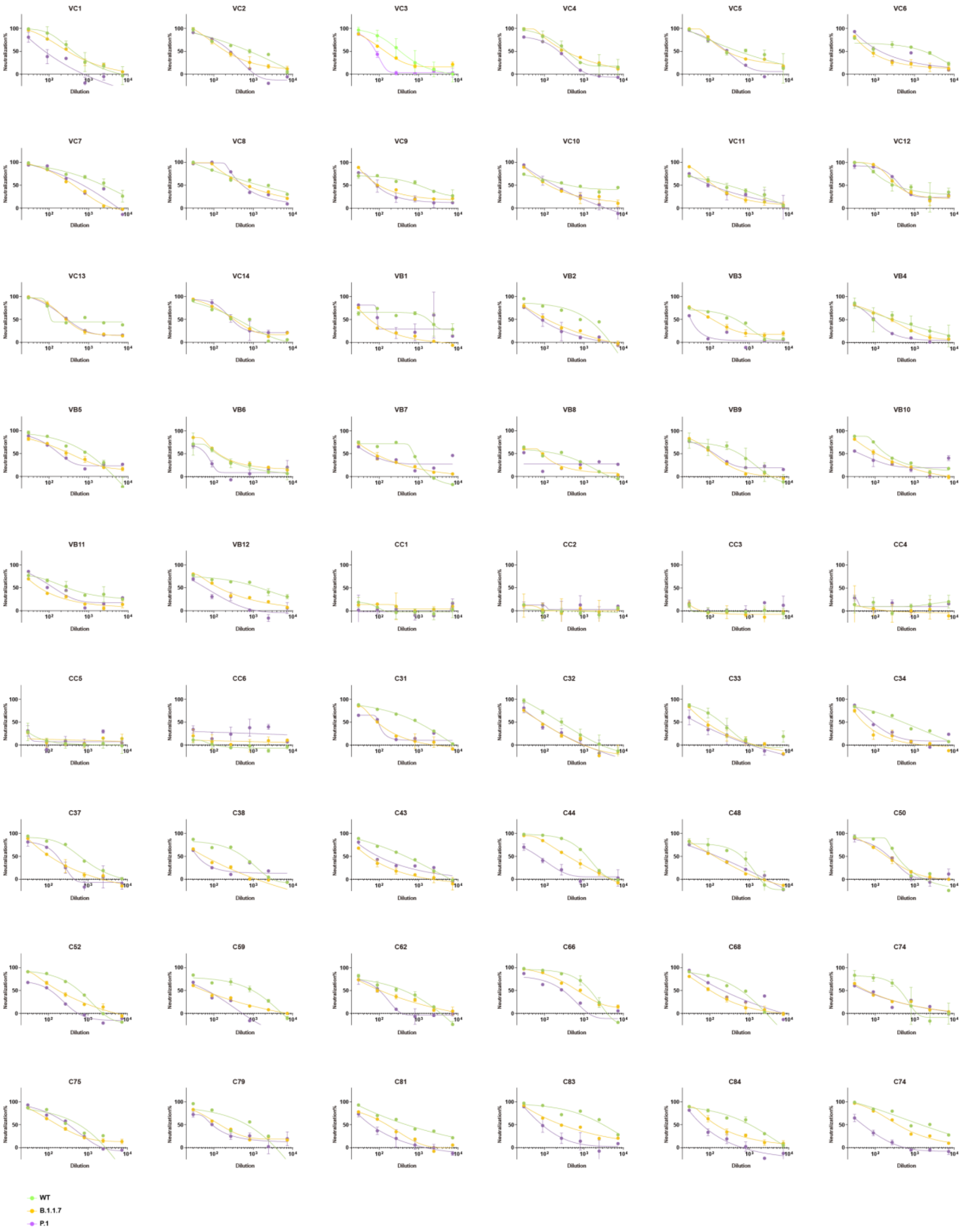
VC1
VC2
VC3
VC4
VC5
VC6
VC7
VC8
VC9
VC10
VC11
VC12
VC13
VC14
VB1
VB2
VB3
VB4
VB5
VB6
VB7
VB8
VB9
VB10
VB11
VB12
CC1
CC2
CC3
CC4
CC5
CC6
C31
C32
C33
C34
C37
C38
C43
C44
C48
C50
C52
C59
C62
C66
C68
C74
C75
C79
C81
C83
C84
C74
Neutralization%
Dilution
WT
B.1.1.7
P.1

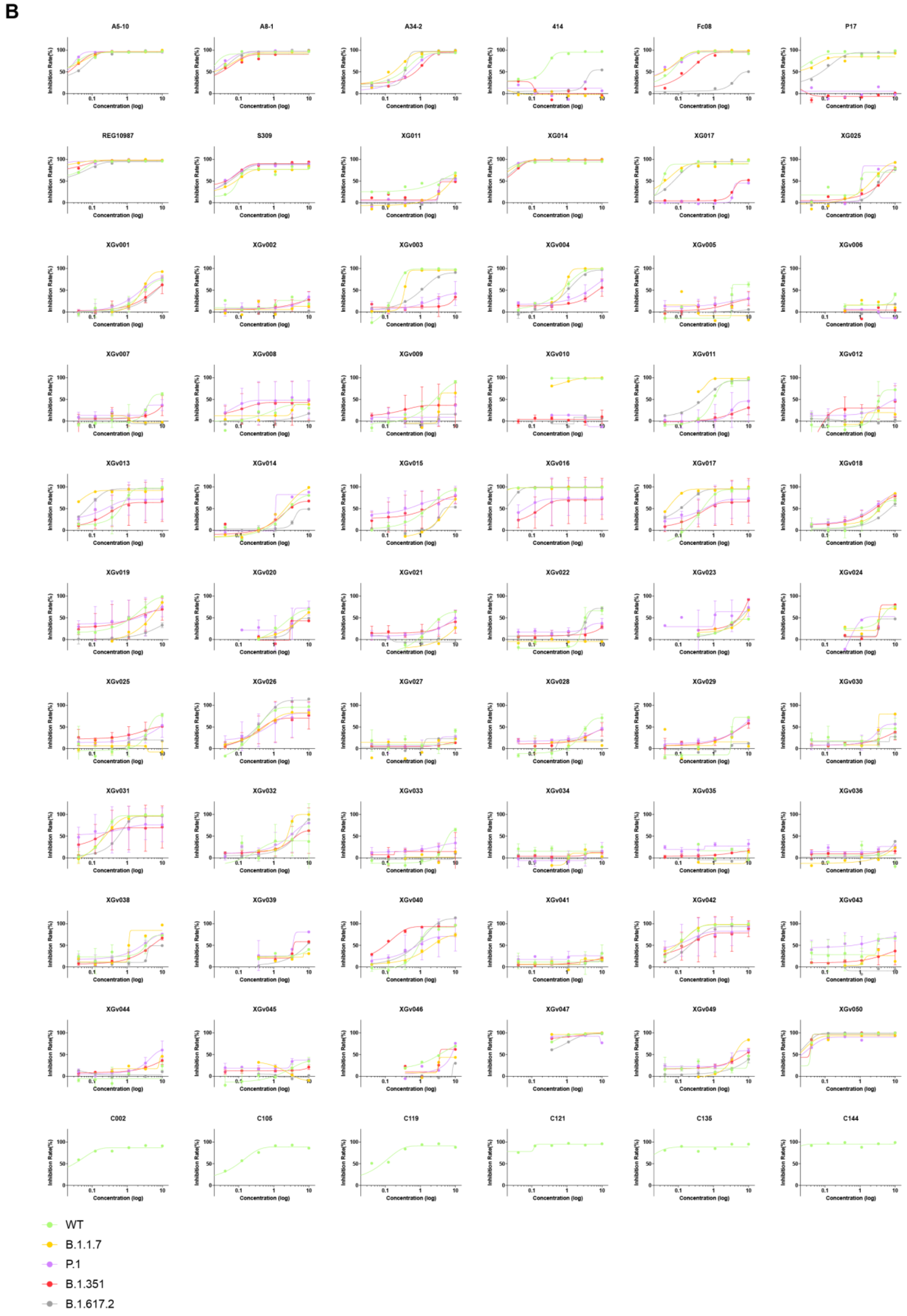

**Fig. S10**. **Neutralization profiles of plasma and NAbs against both the WT and variant pseudoviruses.**

(A) Neutralization assays of WT, B.1.1.7, B.1.351, P.1 and B.1.617.2 pseudoviruses using plasma from convalescents (C1-C22), 2-dose (VB1-VB12) and 3-dose (VC1-VC14) CoronaVac vaccinees, as well as healthy individuals (CC1-CC6).

(B) Neutralization curves of the XGv mAbs and other representative mAbs analyzed in Fig. 3E against pseudotyped viruses of both WT and VOCs. Values of each mAb are listed in Table S3.

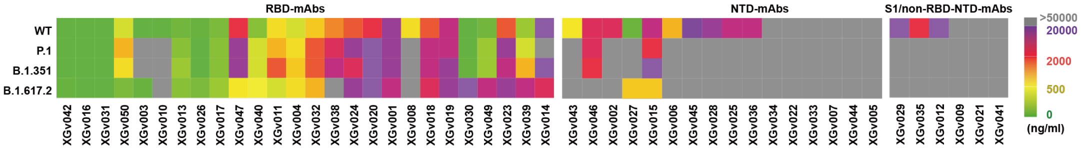

**Fig. S11 Neutralization of monoclonal antibodies against authentic virus.**
Neutralization activity of the 48 mAbs against pseudoviruses bearing the S protein of WT (green), B.1.351 (red), P.1 (purple), and B.1.617.2 (grey).

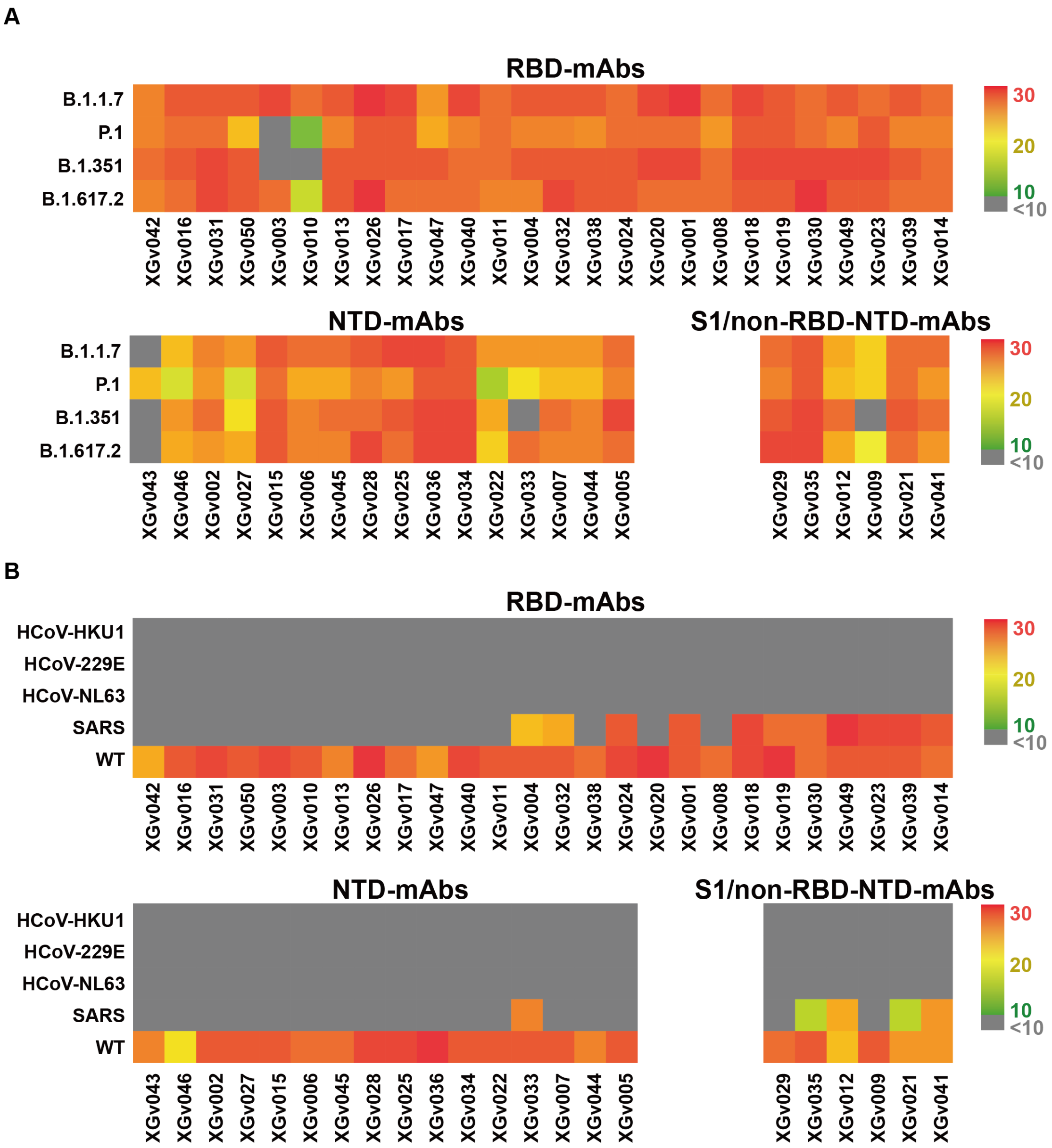

**Fig. S12 Antibody binding assay.**

Heat map shows the binding activities calculated by AUC of clonally related antibodies against SARS-CoV-2 over variants of concerns (A) and species (B).

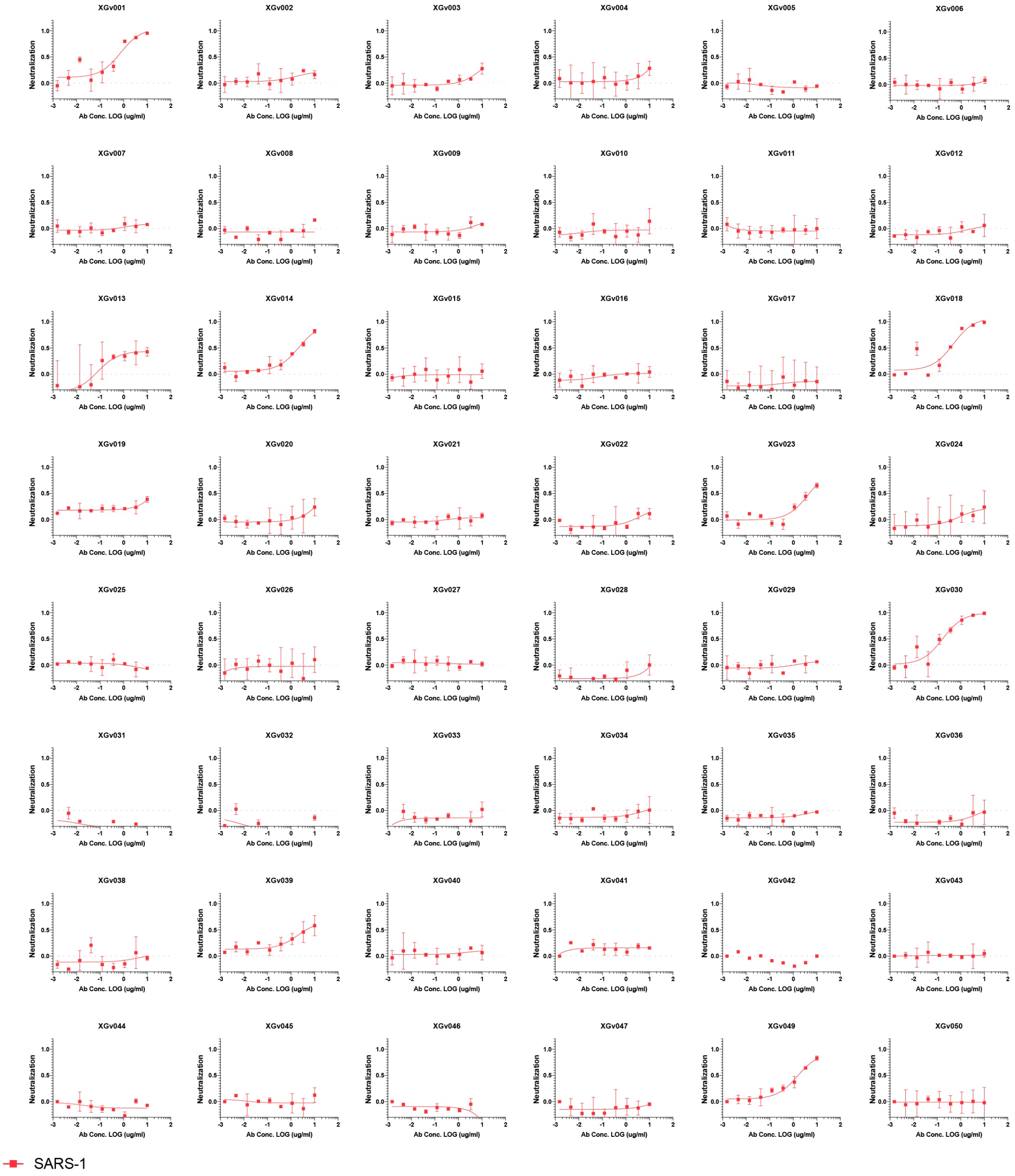

**Fig. S13 Neutralization activity of monoclonal antibodies from 3-dose inactivated vaccine immunized individuals against SARS-CoV typed pseudovirus.**

Data are mean ± s.e.m. of technical triplicates.

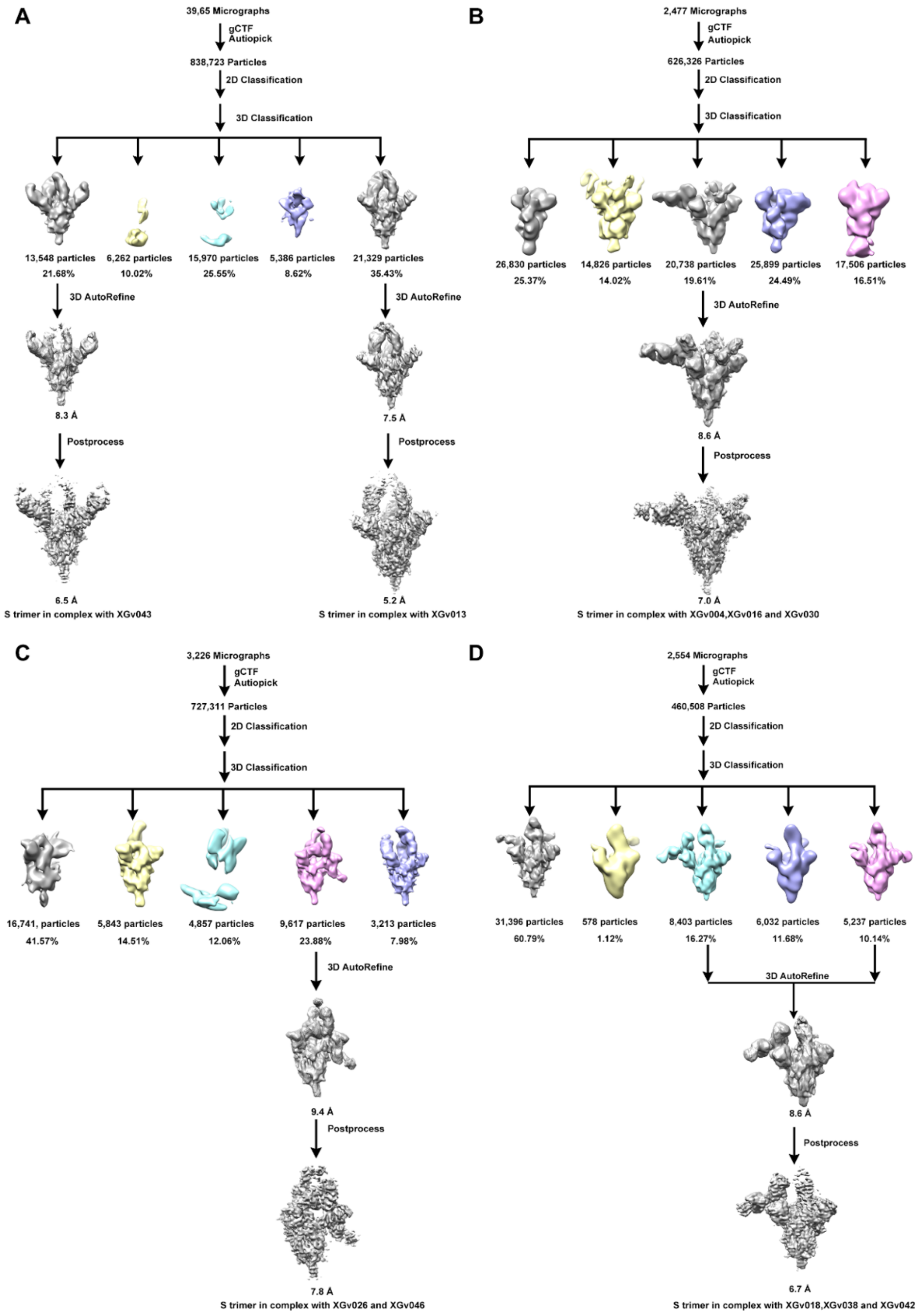

**Fig. S14 Flowchart for Cryo-EM data processing.**

Flowchart for SARS-CoV-2 S-XGv013-XGv043 (**A**) , S-XGv004-XGv016-XGv030 (**B**), S-XGv026-XGv046 (**C**) and S-XGv018-XGv038-XGv042 (**D**) Cryo-EM data processing are shown.

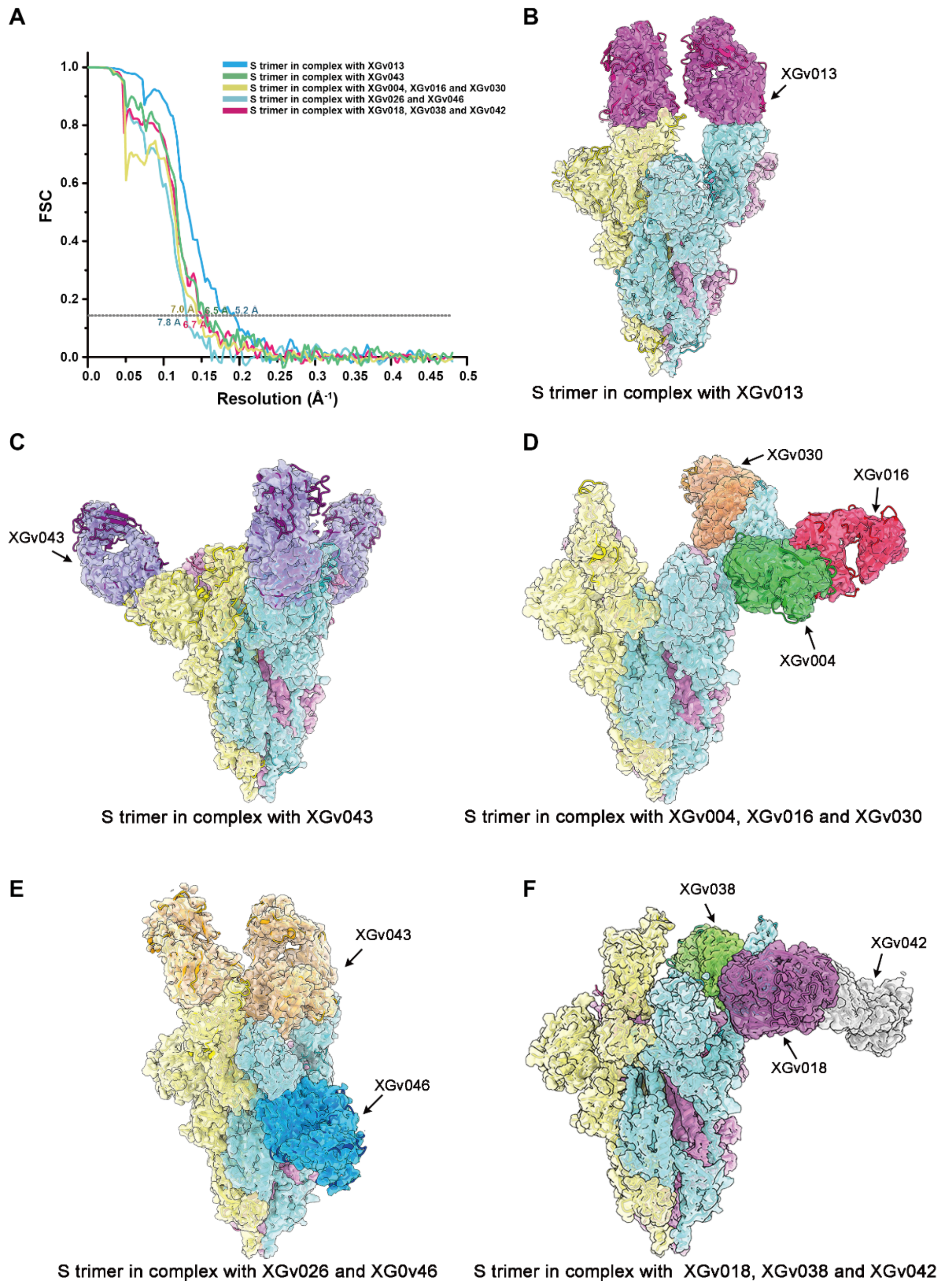

**Fig. S15 Evaluation of resolution and density maps with resulting atomic models of SARS-CoV-2 S trimer in complex with Fabs.**

(**A**)The gold-standard FSC curves of the final maps. (**B-F**) Cryo-EM maps of SARS-CoV-2 trimer in complex with Fabs described above. Each monomer of S trimer is colored by cyan, pink and yellow, respectively.

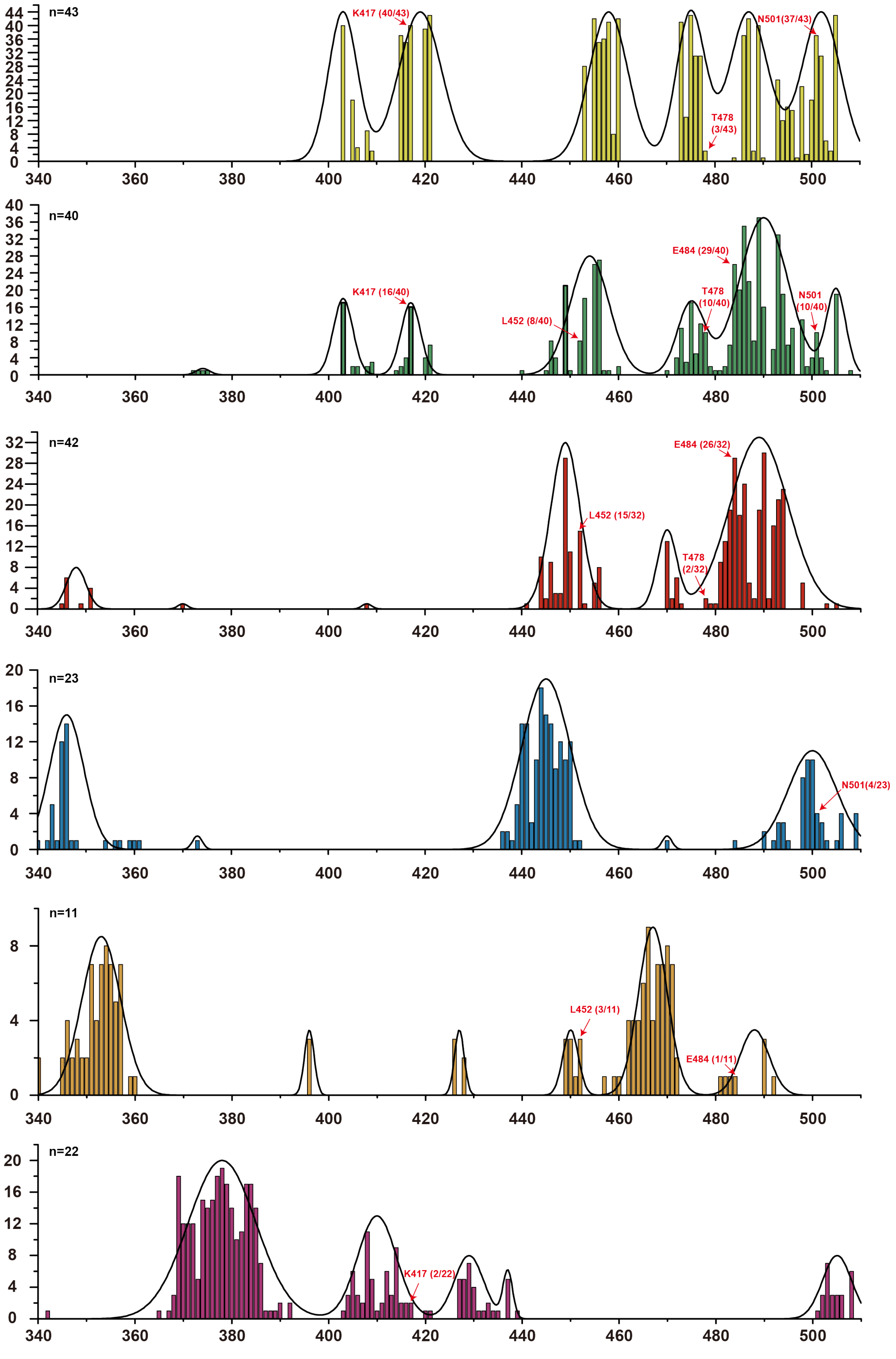

**Fig. S16 Distribution of the antigenic sites for the 6 antibody classes.**

Frequency of each epitope of six classes of antibodies is counted and displayed by histogram. Distribution for six antibody classes is shown by fitted curves. Color scheme is the same as in Fig. 3A.

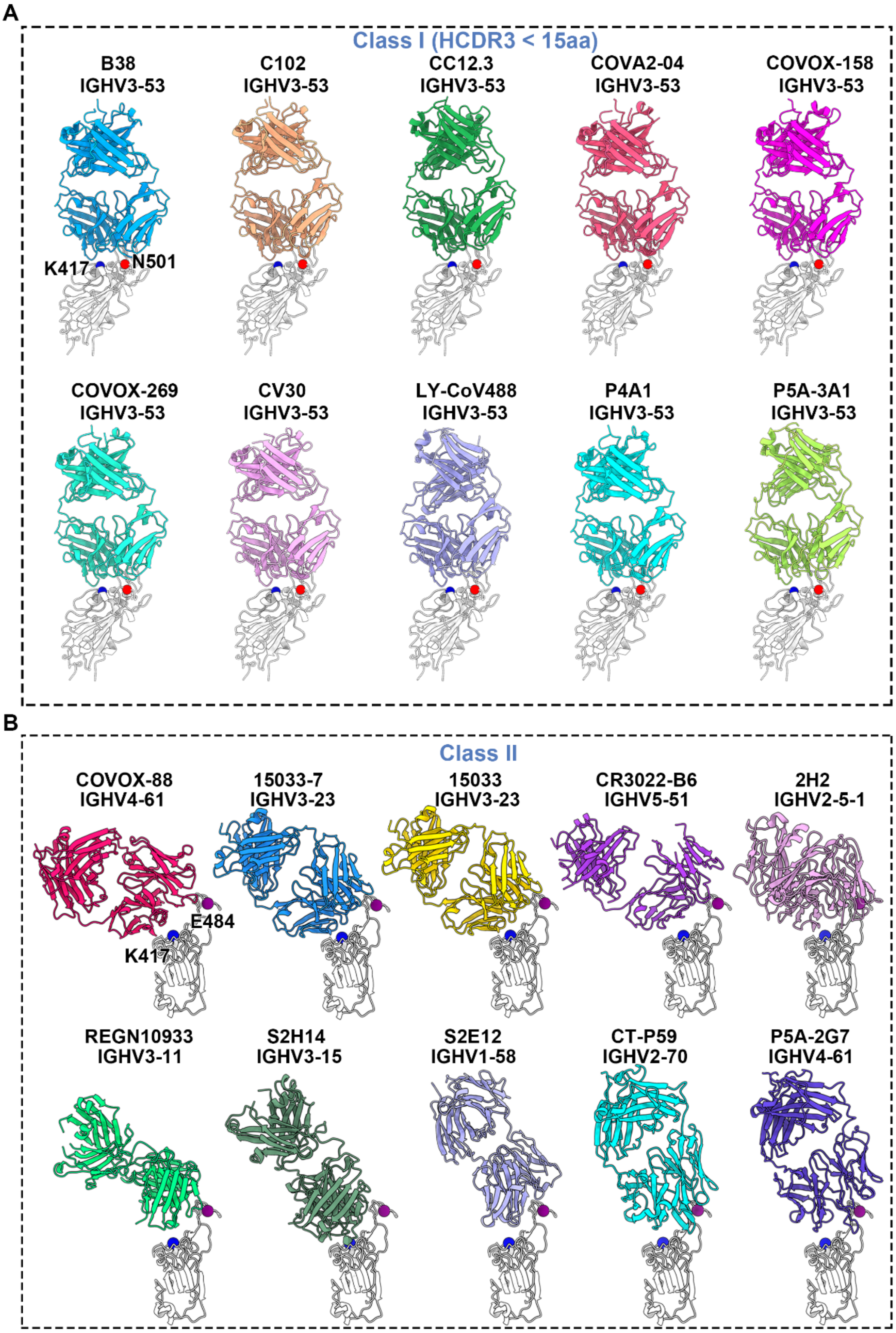

A
Class I (HCDR3 < 15aa)
B38
IGHV3-53
C102
IGHV3-53
CC12.3
IGHV3-53
COVA2-04
IGHV3-53
COVOX-158
IGHV3-53
K417
N501
COVOX-269
IGHV3-53
CV30
IGHV3-53
LY-CoV488
IGHV3-53
P4A1
IGHV3-53
P5A-3A1
IGHV3-53
B
Class II
COVOX-88
IGHV4-61
15033-7
IGHV3-23
15033
IGHV3-23
CR3022-B6
IGHV5-51
2H2
IGHV2-5-1
E484
K417
REGN10933
IGHV3-11
S2H14
IGHV3-15
S2E12
IGHV1-58
CT-P59
IGHV2-70
P5A-2G7
IGHV4-61

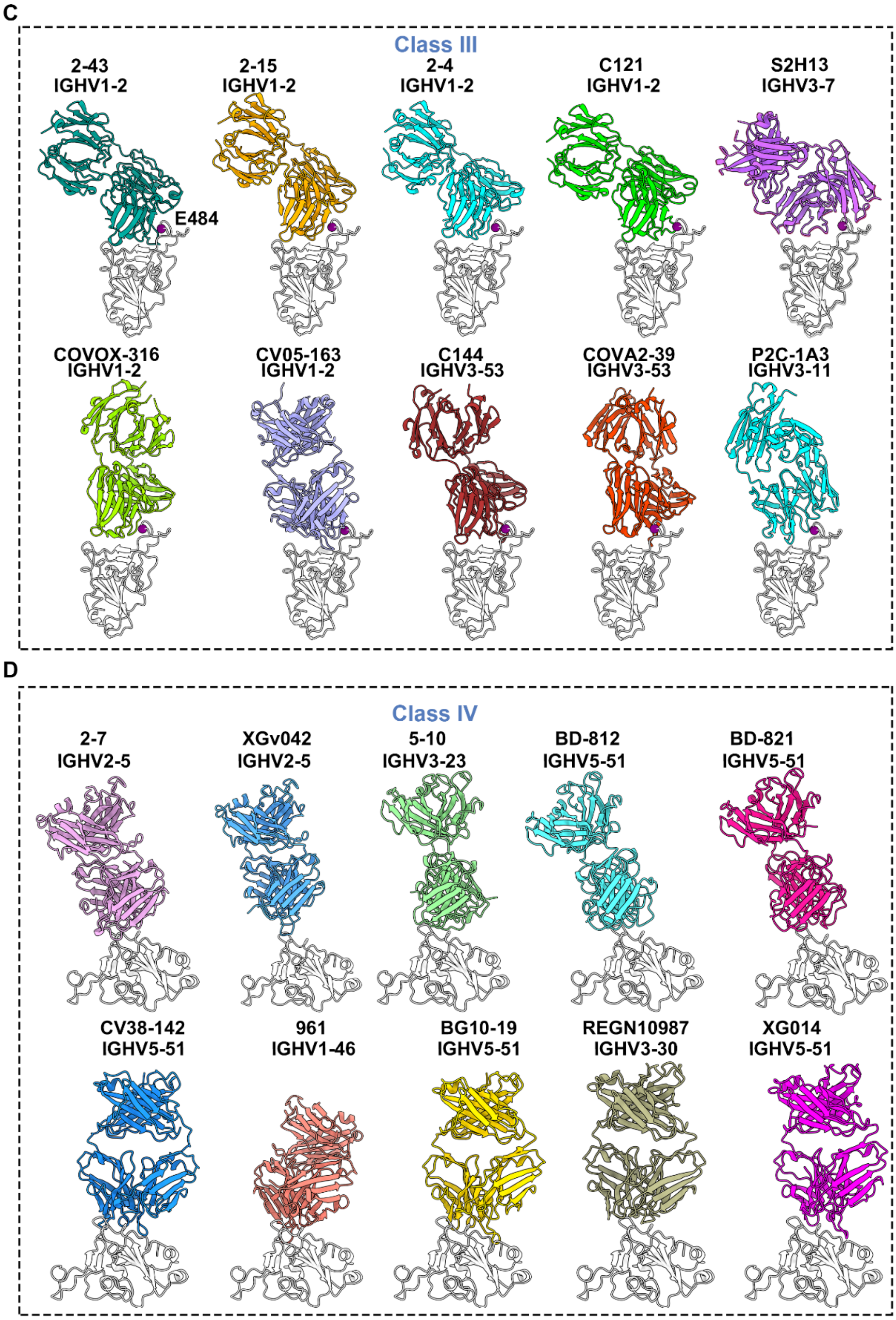
C
Class III
2-43
IGHV1-2
E484
2-15
IGHV1-2
2-4
IGHV1-2
C121
IGHV1-2
S2H13
IGHV3-7
COVOX-316
IGHV1-2
CV05-163
IGHV1-2
C144
IGHV3-53
COVA2-39
IGHV3-53
P2C-1A3
IGHV3-11
D
Class IV
2-7
IGHV2-5
XGv042
IGHV2-5
5-10
IGHV3-23
BD-812
IGHV5-51
BD-821
IGHV5-51
CV38-142
IGHV5-51
961
IGHV1-46
BG10-19
IGHV5-51
REGN10987
IGHV3-30
XG014
IGHV5-51

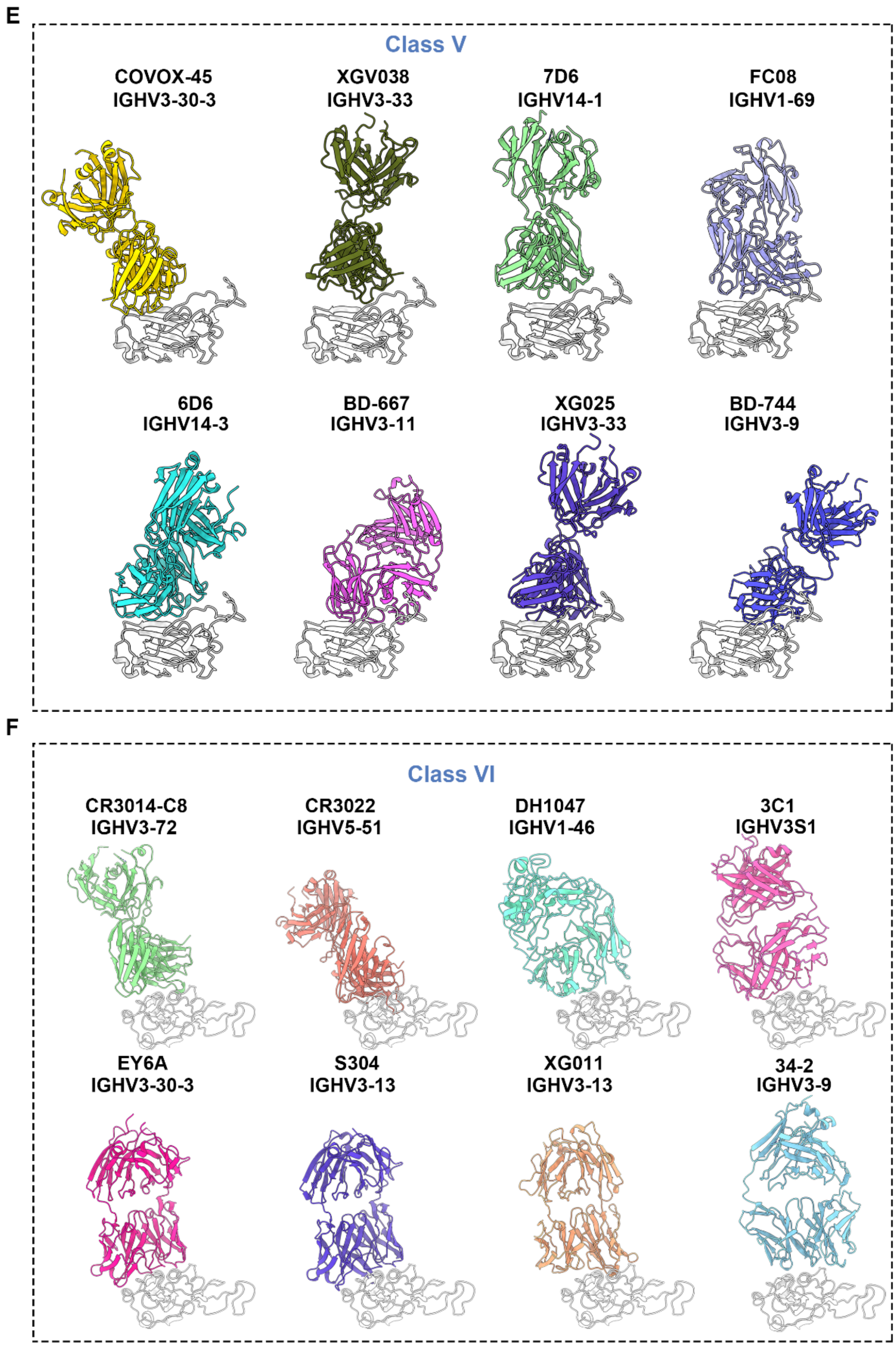

**Fig. S17 Structures of 6 classes of antibodies in complex with SARS-CoV-2 RBD.** Structures of SARS-CoV-2 RBD in complex with antibodies of six classes are shown (**A-F**) as cartoon. RBD is colored by white. Residues K417, E484 and N501 are shown as sphere and colored by blue, purple and red, respectively.

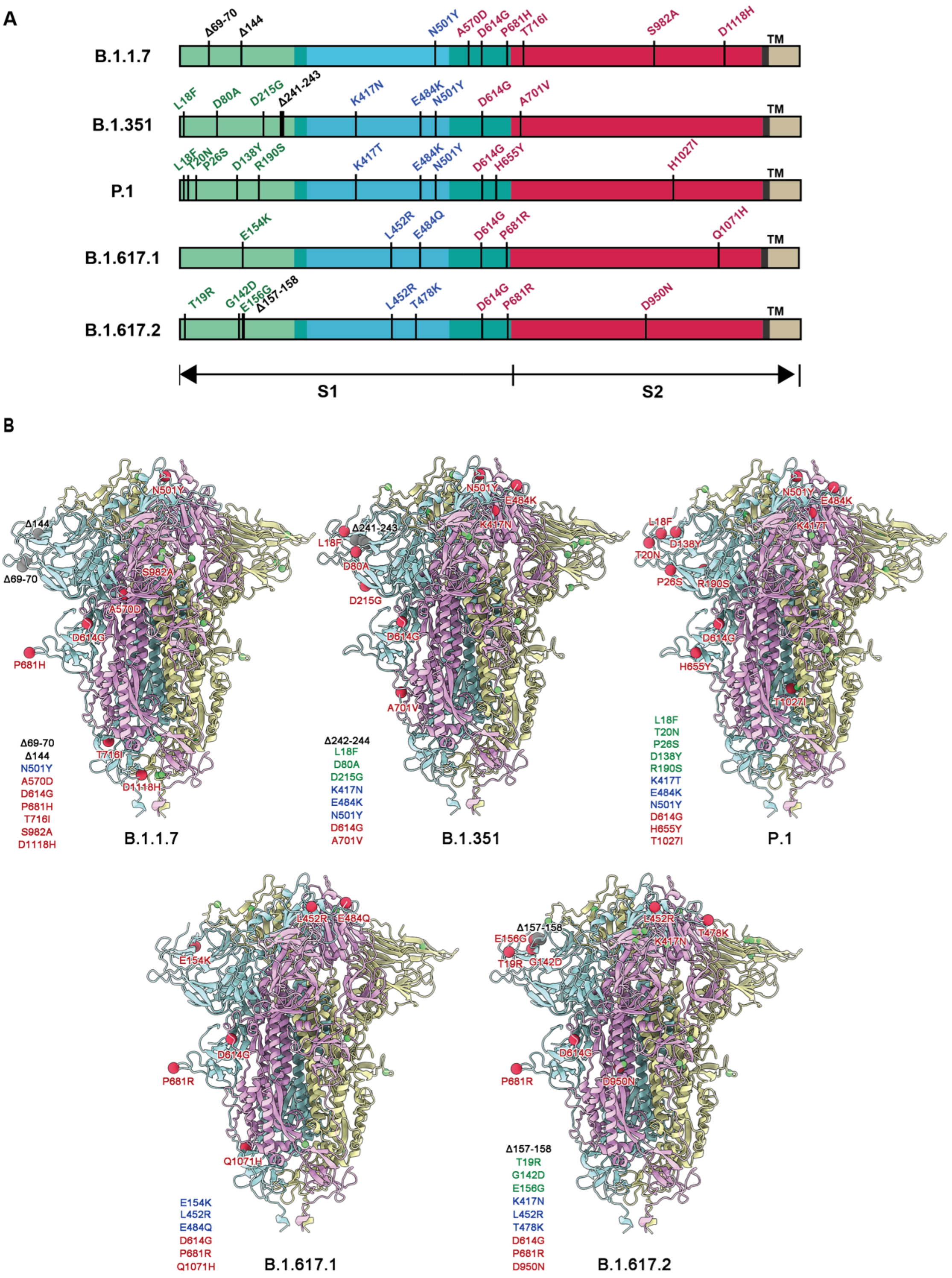

**Fig. S18 Mutation residues of SARS-CoV-2 variants.**

(**A**) Schematic diagram showing the mutation and deletion sites of SARS-CoV-2 variants (B.1.1.7, B.1.351, P.1, B.1.617.1 and B.1.617.2). (**B**) S trimers of variants are shown as cartoon and mutations and deletions are shown as spheres. Mutation sites are colored by red and deletion sites are colored by grey on one monomer (cyan), and both are colored by green on the other two monomers (pink and yellow).

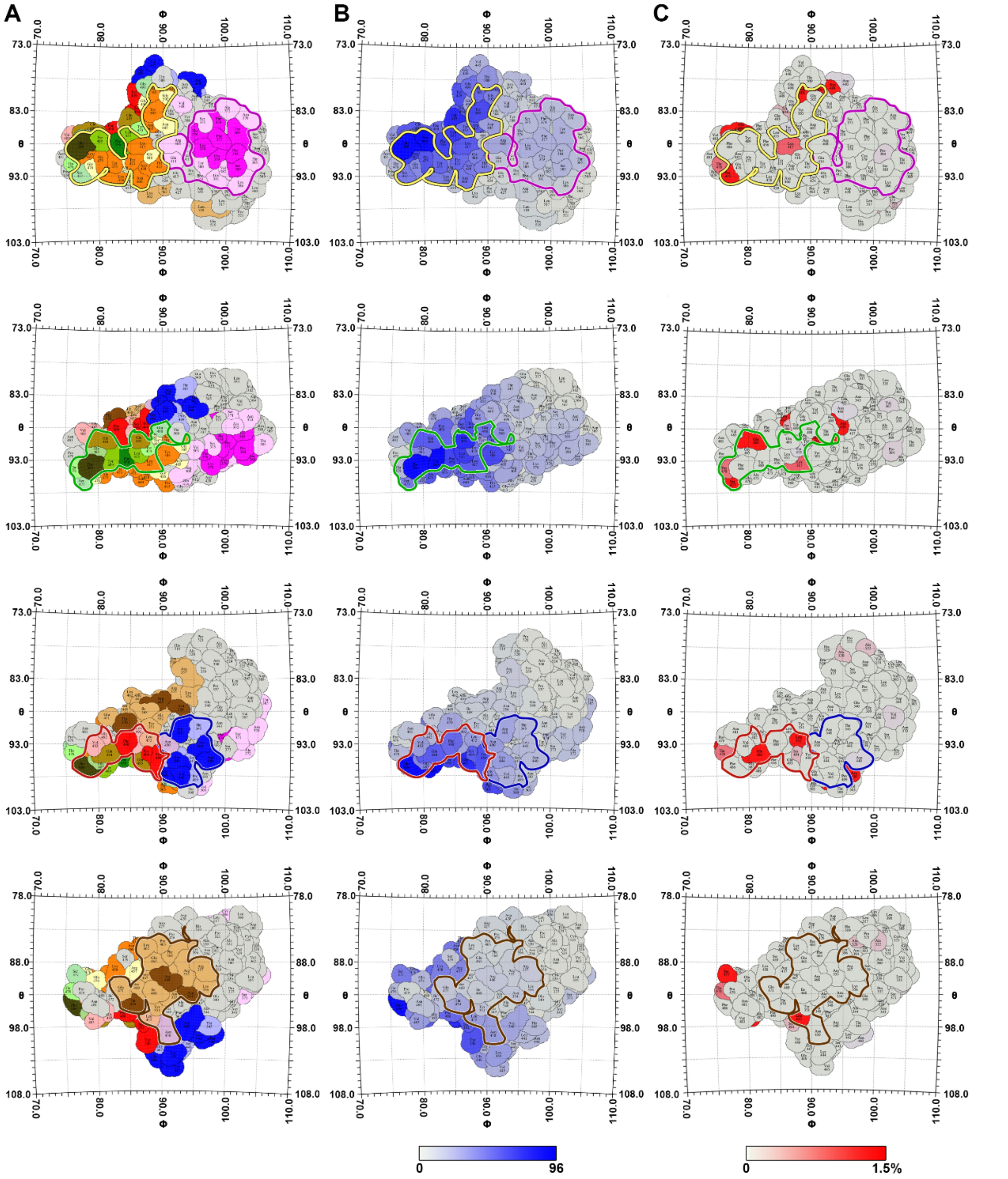

**Fig. S19 Structural landscape of footprints on RBD surface.**

A two-dimensional projection of the RBD surface was produced using RIVEM. Residues of RBD are colored in gray. (A) Residues of RBD belonging to antigenic patches (with targeting frequency >30%) recognized by six classes of NAbs are colored in the assigned color scheme (same to Fig. 3C), among which residues with “hot targeting frequency” (generally over 65%, but over 85% in class I) are shown in bright colors corresponding to the patches they belong to. Residues involved in two (such as Y489, L452) or three (such as F486) antigenic patches are shown in a mixed color. (B) Heat map of antibody binding number. Residues are colored by the frequency of recognition by 171 Nabs. (C) Heat map for circulating variants with mutations on the RBD. Mutation frequency for each residue was calculated based on the datasets from GISAID.

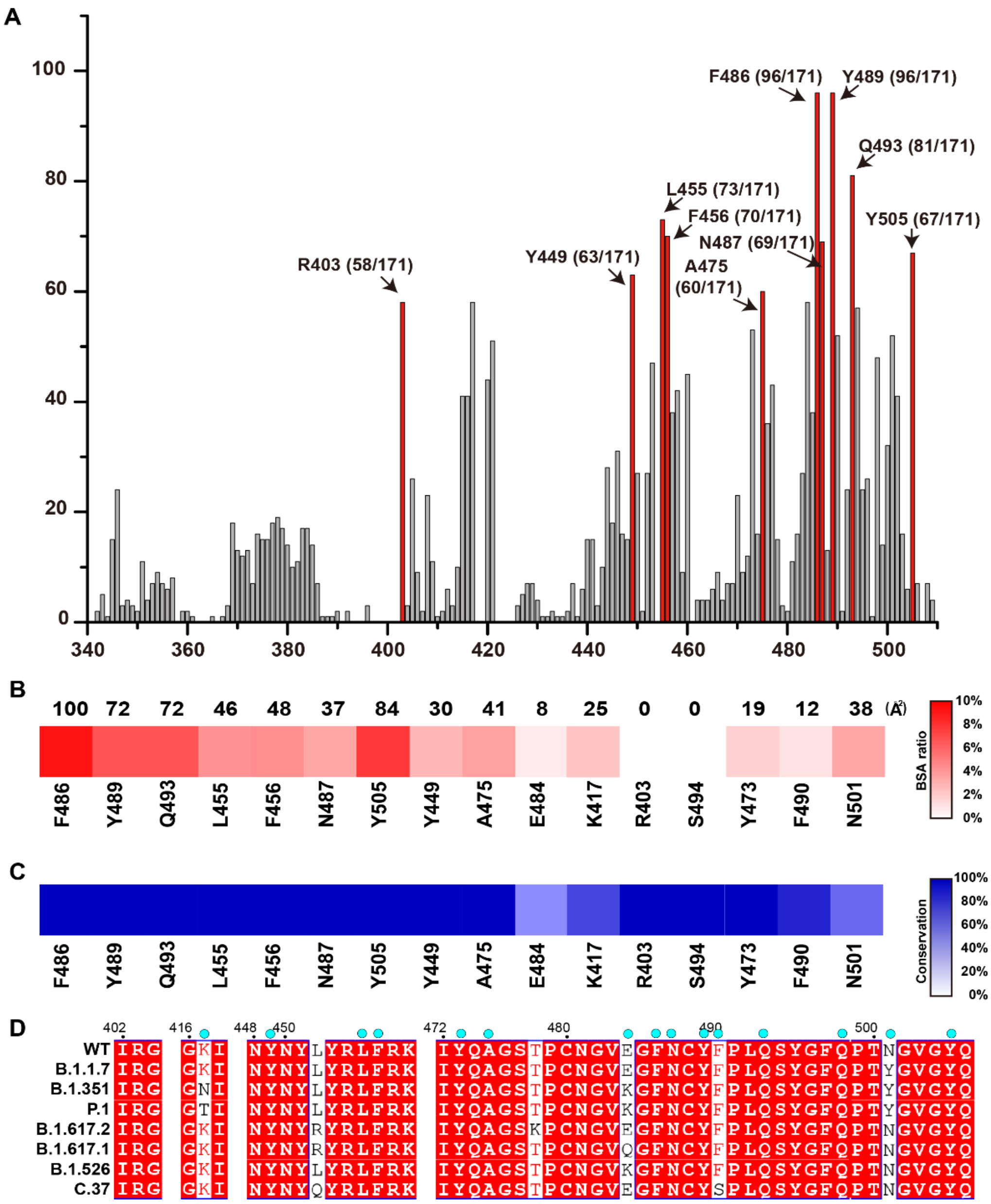

**Fig. S20 Immunogenic analysis of the residues on SARS-CoV-2 RBD.**

(A) Immunogenic analysis of the residues on SARS-CoV-2 RBD based on 171 Nabs. The vertical axis represents the number of Nabs that neutralize a certain site. Bars of the 10 hottest immunogenic residues in the histogram are filling with red. (B) Buried surface area (BSA) ratio upon binding to ACE2 of the 16 hottest immunogenic residues. BSA of these residues are listed above the chart. (C) Conservation of the 16 hottest immunogenic residues in eight SARS-CoV-2 strains (WT, B.1.1.7, B.1.351, P.1, B.1.617.2, B.1.617.1, B.1.526 and C.37). (D) Sequence alignment of SARS-CoV-2 WT and variant B.1.1.7, B.1.351, P.1, B.1.617.2, B.1.617.1, B.1.526 and C.37. The residues involved in direct interactions with ACE2 are marked with cyan balls.

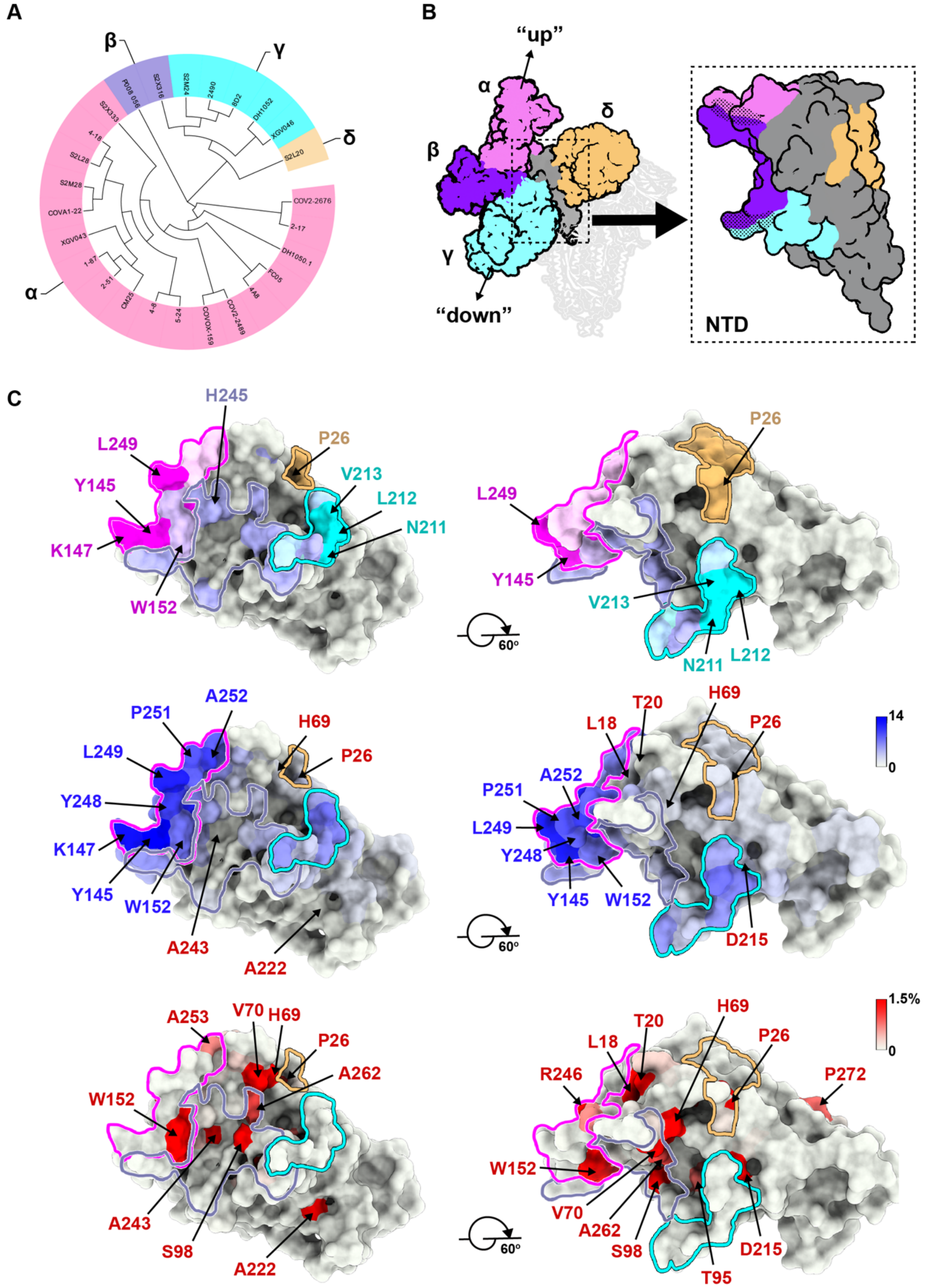

**Fig. S21 Structural landscape of NTD NAbs.**

(A) Structure-based antigenic clustering of SARS-CoV-2 NTD NAbs. A total of 26 NTD NAbs with available structures were classified into four clusters (α, β, γ and δ).

(B) Surface representative model of four types of NAbs bound to the NTD. Fab fragments of four representative antibody are shown in different colors and the NTD is colored in gray. Insets illustrate the antigenic patches targeted by four representative antibodies. Dashed dots indicate the overlaps between two adjacent antigenic patches. (C) Structural landscapes of the four classes of NTD NAbs (upper panel). Antigenic patches (with targeting frequency >30%) recognized by four classes of NAbs are outlined in the assigned color scheme (same to Fig. S21B), among which residues with "hot targeting frequency" (generally over 60%) are shown in bright colors corresponding to the patches they belong to. Residues involved in two (such as W152, Q183 and R214) neighboring antigenic patches are presented in a mixed color. Representative "hot" antigenic residues are labeled. Middle: hot map for antigenic residues on the NTD. Per residue frequency recognized by the 26 NAbs were calculated and shown. The key residues with substitutions in several VOCs are marked and labeled. Bottom: hot map for circulating variants with mutations on the NTD. Mutation frequency for each residue was calculated based on the datasets from GISAID.

**A**

| | | XGv026 | XGv017 | XGv013 | XGv040 | XGv050 | XGv003 | XGv031 | XGv010 | XGv011 | XGv042 | XGv016 | XGv047 | XGv038 | XGv008 | XGv030 | XGv020 | XGv014 | XGv004 | XGv032 | XGv024 | XGv019 | XGv023 | XGv039 | XGv001 | XGv049 | XGv018 |
|---|---|---|---|---|---|---|---|---|---|---|---|---|---|---|---|---|---|---|---|---|---|---|---|---|---|---|---|
| XGv026 | | 4 | 6 | 6 | 29 | 6 | 5 | 5 | 102 | 103 | 108 | 102 | 95 | 105 | 101 | 102 | 103 | 100 | 103 | 108 | 109 | 103 | 108 | 107 | 94 | 104 | 100 |
| XGv017 | | 5 | 6 | 7 | 33 | 6 | 5 | 5 | 101 | 101 | 101 | 100 | 99 | 101 | 100 | 101 | 95 | 102 | 102 | 101 | 102 | 100 | 103 | 102 | 95 | 105 | 100 |
| XGv013 | | 4 | 5 | 5 | 28 | 5 | 4 | 4 | 104 | 105 | 105 | 104 | 97 | 105 | 105 | 105 | 99 | 105 | 104 | 100 | 105 | 106 | 106 | 106 | 98 | 101 | 100 |
| XGv040 | | 4 | 4 | 3 | 6 | 3 | 4 | 3 | 11 | 104 | 102 | 103 | 98 | 101 | 101 | 101 | 98 | 103 | 105 | 102 | 103 | 103 | 103 | 102 | 97 | 110 | 98 |
| XGv050 | | 54 | 71 | 72 | 52 | 8 | 62 | 62 | 92 | 102 | 101 | 100 | 94 | 104 | 101 | 103 | 98 | 102 | 102 | 103 | 105 | 100 | 105 | 103 | 96 | 102 | 101 |
| XGv003 | | 24 | 29 | 36 | 89 | 15 | 6 | 6 | 98 | 97 | 102 | 102 | 63 | 102 | 102 | 102 | 96 | 101 | 101 | 102 | 102 | 101 | 104 | 103 | 95 | 105 | 100 |
| XGv031 | | 15 | 24 | 25 | 67 | 8 | 7 | 5 | 86 | 102 | 101 | 104 | 94 | 100 | 102 | 101 | 97 | 103 | 103 | 100 | 104 | 102 | 102 | 102 | 97 | 102 | 98 |
| XGv010 | | 12 | 13 | 18 | 29 | 5 | 5 | 5 | 8 | 5 | 11 | 38 | 4 | 13 | 48 | 5 | 106 | 111 | 110 | 110 | 111 | 110 | 110 | 110 | 104 | 108 | 97 |
| XGv011 | | 103 | 103 | 103 | 104 | 106 | 45 | 50 | 60 | 9 | 47 | 78 | 7 | 36 | 82 | 11 | 95 | 103 | 103 | 102 | 104 | 103 | 103 | 103 | 95 | 105 | 100 |
| XGv042 | | 102 | 102 | 98 | 103 | 100 | 105 | 103 | 22 | 8 | 6 | 4 | 6 | 102 | 104 | 102 | 99 | 98 | 103 | 103 | 102 | 104 | 103 | 103 | 98 | 102 | 96 |
| XGv016 | | 103 | 103 | 104 | 104 | 118 | 102 | 105 | 102 | 87 | 34 | 9 | 103 | 102 | 102 | 100 | 97 | 103 | 104 | 104 | 104 | 104 | 104 | 104 | 100 | 118 | 112 |
| XGv047 | | 101 | 107 | 108 | 101 | 103 | 43 | 37 | 58 | 17 | 42 | 107 | 6 | 30 | 91 | 15 | 91 | 105 | 109 | 102 | 102 | 107 | 101 | 102 | 102 | 103 | 99 |
| XGv038 | | 103 | 102 | 105 | 102 | 104 | 104 | 104 | 90 | 17 | 102 | 102 | 12 | 5 | 18 | 4 | 14 | 99 | 102 | 95 | 95 | 96 | 102 | 101 | 95 | 103 | 78 |
| XGv008 | | 98 | 103 | 104 | 100 | 114 | 104 | 100 | 22 | 7 | 98 | 103 | 8 | 4 | 6 | 3 | 6 | 13 | 95 | 63 | 70 | 93 | 88 | 82 | 94 | 104 | 72 |
| XGv030 | | 104 | 104 | 104 | 105 | 106 | 104 | 105 | 82 | 104 | 105 | 103 | 17 | 7 | 25 | 5 | 23 | 70 | 101 | 99 | 99 | 103 | 103 | 102 | 96 | 103 | 86 |
| XGv020 | | 105 | 105 | 107 | 110 | 119 | 107 | 110 | 104 | 87 | 111 | 106 | 27 | 6 | 29 | 5 | 9 | 60 | 106 | 97 | 96 | 103 | 106 | 105 | 101 | 114 | 76 |
| XGv014 | | 107 | 108 | 108 | 106 | 126 | 108 | 107 | 102 | 108 | 106 | 107 | 114 | 68 | 24 | 7 | 18 | 6 | 109 | 20 | 20 | 16 | 8 | 13 | 7 | 11 | 6 |
| XGv004 | | 104 | 104 | 105 | 104 | 109 | 105 | 106 | 104 | 105 | 104 | 104 | 105 | 103 | 102 | 103 | 98 | 103 | 7 | 10 | 19 | 13 | 7 | 7 | 6 | 13 | 5 |
| XGv032 | | 103 | 105 | 105 | 104 | 105 | 106 | 104 | 104 | 104 | 104 | 103 | 99 | 95 | 99 | 103 | 99 | 75 | 6 | 6 | 9 | 5 | 5 | 4 | 4 | 5 | 3 |
| XGv024 | | 104 | 105 | 105 | 104 | 106 | 104 | 103 | 103 | 103 | 106 | 104 | 101 | 94 | 102 | 102 | 98 | 89 | 17 | 13 | 7 | 6 | 4 | 5 | 5 | 5 | 4 |
| XGv019 | | 101 | 105 | 105 | 105 | 105 | 105 | 103 | 104 | 104 | 105 | 105 | 101 | 94 | 101 | 100 | 97 | 81 | 19 | 16 | 9 | 8 | 5 | 6 | 5 | 5 | 4 |
| XGv023 | | 91 | 106 | 106 | 103 | 112 | 107 | 99 | 95 | 95 | 102 | 105 | 108 | 86 | 83 | 96 | 88 | 69 | 22 | 20 | 10 | 9 | 6 | 7 | 6 | 13 | 6 |
| XGv039 | | 103 | 110 | 110 | 105 | 101 | 111 | 106 | 108 | 111 | 105 | 110 | 96 | 97 | 105 | 104 | 102 | 82 | 22 | 23 | 12 | 9 | 7 | 6 | 7 | 7 | 4 |
| XGv001 | | 103 | 104 | 105 | 104 | 110 | 104 | 104 | 102 | 105 | 104 | 104 | 105 | 95 | 102 | 102 | 97 | 84 | 29 | 28 | 18 | 14 | 7 | 9 | 6 | 8 | 6 |
| XGv049 | | 104 | 103 | 104 | 102 | 103 | 102 | 106 | 101 | 103 | 104 | 102 | 96 | 103 | 101 | 104 | 102 | 99 | 40 | 29 | 17 | 13 | 7 | 8 | 7 | 6 | 6 |
| XGv018 | | 108 | 102 | 102 | 104 | 102 | 102 | 109 | 101 | 101 | 107 | 102 | 97 | 104 | 101 | 108 | 99 | 100 | 57 | 44 | 26 | 21 | 9 | 11 | 10 | 8 | 8 |
| XG017 | | 4 | 4 | 4 | 8 | 4 | 4 | 4 | 87 | 105 | 106 | 105 | 105 | 116 | 102 | 119 | 118 | 106 | 106 | 122 | 122 | 107 | 123 | 121 | 103 | 110 | 108 |
| XG014 | | 118 | 104 | 102 | 102 | 110 | 104 | 120 | 46 | 6 | 8 | 6 | 7 | 94 | 97 | 121 | 118 | 60 | 100 | 115 | 119 | 103 | 122 | 115 | 93 | 112 | 116 |
| XG025 | | 108 | 101 | 95 | 76 | 99 | 109 | 122 | 15 | 5 | 99 | 107 | 5 | 4 | 5 | 4 | 6 | 16 | 96 | 62 | 67 | 86 | 104 | 92 | 65 | 96 | 92 |
| XG011 | | 118 | 107 | 107 | 94 | 126 | 108 | 125 | 99 | 99 | 100 | 110 | 126 | 96 | 81 | 120 | 116 | 42 | 7 | 7 | 6 | 5 | 5 | 5 | 4 | 5 | 5 |
| ACE2 | | 3 | 4 | 4 | 3 | 3 | 4 | 4 | 4 | 12 | 3 | 4 | 7 | 37 | 52 | 41 | 50 | 88 | 6 | 6 | 33 | 36 | 38 | 30 | 49 | 38 | 27 |

**B**

| | | XGv013 | P4A1 | XGv026 | XG017 | H4 | 414 | XGv003 | XGv031 | P17 | XG014 | XGv016 | XGv042 | XG025 | FC08 | XGv030 | XG011 | A34-2 | XGv004 | FC05 |
|---|---|---|---|---|---|---|---|---|---|---|---|---|---|---|---|---|---|---|---|---|
| BD906 | | 3 | 4 | 3 | 3 | 12 | 3 | 6 | 5 | 12 | 4 | 17 | 13 | 79 | 82 | 61 | 73 | 58 | 60 | 93 |
| BD912 | | 3 | 4 | 3 | 3 | 12 | 3 | 4 | 4 | 7 | 4 | 80 | 89 | 75 | 74 | 85 | 67 | 70 | 59 | 89 |
| BD837 | | 3 | 4 | 3 | 3 | 12 | 3 | 4 | 5 | 11 | 5 | 72 | 89 | 74 | 66 | 69 | 62 | 52 | 50 | 88 |
| BD930 | | 4 | 6 | 6 | 4 | 11 | 4 | 20 | 19 | 106 | 9 | 44 | 48 | 58 | 108 | 56 | 38 | 57 | 67 | 81 |
| VA5-1 | | 6 | 9 | 8 | 10 | 39 | 33 | 20 | 17 | 54 | 33 | 78 | 91 | 74 | 75 | 75 | 74 | 78 | 75 | 99 |
| VA5-2 | | 12 | 9 | 7 | 11 | 75 | 34 | 16 | 15 | 77 | 55 | 94 | 80 | 83 | 77 | 66 | 88 | 78 | 90 | 97 |
| VA5-9 | | 10 | 9 | 8 | 10 | 43 | 30 | 17 | 17 | 26 | 18 | 91 | 85 | 67 | 83 | 82 | 99 | 88 | 94 | 103 |
| VA8-3 | | 9 | 12 | 11 | 10 | 23 | 24 | 21 | 19 | 51 | 28 | 93 | 96 | 83 | 93 | 102 | 82 | 102 | 89 | 97 |
| VA8-4 | | 14 | 9 | 8 | 18 | 45 | 42 | 7 | 10 | 14 | 21 | 19 | 13 | 49 | 86 | 90 | 74 | 83 | 86 | 95 |
| VA8-5 | | 13 | 13 | 11 | 12 | 35 | 37 | 15 | 19 | 23 | 11 | 19 | 13 | 94 | 104 | 91 | 87 | 95 | 99 | 101 |
| VA9-3 | | 25 | 23 | 12 | 13 | 50 | 36 | 33 | 31 | 66 | 55 | 108 | 85 | 93 | 81 | 84 | 91 | 95 | 98 | 100 |
| VA9-5 | | 21 | 22 | 10 | 10 | 41 | 29 | 26 | 28 | 62 | 56 | 98 | 100 | 98 | 96 | 107 | 83 | 92 | 91 | 104 |
| VA13-6 | | 7 | 12 | 9 | 7 | 46 | 37 | 27 | 82 | 40 | 39 | 83 | 64 | 80 | 76 | 64 | 70 | 92 | 82 | 89 |
| VA13-7 | | 7 | 9 | 11 | 10 | 49 | 38 | 26 | 114 | 37 | 43 | 75 | 87 | 81 | 83 | 81 | 83 | 152 | 88 | 92 |
| VA13-9 | | 8 | 9 | 13 | 9 | 54 | 39 | 25 | 94 | 38 | 44 | 76 | 80 | 94 | 90 | 83 | 75 | 120 | 84 | 98 |
| VA14-3 | | 13 | 18 | 15 | 12 | 50 | 39 | 28 | 159 | 65 | 51 | 95 | 87 | 84 | 91 | 84 | 99 | 97 | 80 | 99 |
| VA14-5 | | 8 | 14 | 10 | 6 | 48 | 37 | 15 | 19 | 81 | 68 | 87 | 94 | 90 | 116 | 103 | 95 | 89 | 79 | 86 |
| VA29-18 | | 9 | 10 | 8 | 8 | 34 | 22 | 24 | 17 | 68 | 50 | 83 | 109 | 85 | 114 | 116 | 99 | 91 | 96 | 104 |
| VA32-1 | | 8 | 9 | 8 | 8 | 44 | 33 | 16 | 16 | 69 | 48 | 108 | 89 | 55 | 87 | 105 | 74 | 93 | 89 | 94 |
| VA32-5 | | 16 | 18 | 13 | 19 | 48 | 35 | 20 | 18 | 50 | 20 | 66 | 52 | 105 | 116 | 93 | 81 | 82 | 81 | 96 |
| VA34-6 | | 14 | 14 | 11 | 14 | 48 | 45 | 14 | 20 | 52 | 24 | 91 | 73 | 129 | 129 | 120 | 116 | 97 | 108 | 95 |
| XG038 | | 16 | 25 | 21 | 16 | 54 | 51 | 36 | 30 | 39 | 22 | 90 | 39 | 77 | 94 | 92 | 106 | 89 | 94 | 88 |
| VA34-7 | | 35 | 33 | 20 | 25 | 50 | 43 | 22 | 24 | 60 | 23 | 56 | 58 | 88 | 111 | 95 | 96 | 83 | 102 | 85 |
| VA5-8 | | 9 | 8 | 7 | 6 | 40 | 31 | 13 | 14 | 39 | 33 | 93 | 109 | 128 | 107 | 107 | 101 | 98 | 100 | 107 |
| BD30-503 | | 7 | 16 | 12 | 6 | 6 | 3 | 51 | 49 | 45 | 14 | 68 | 80 | 88 | 106 | 88 | 93 | 99 | 88 | 91 |
| BD494 | | 20 | 29 | 24 | 19 | 6 | 3 | 86 | 69 | 102 | 49 | 79 | 90 | 83 | 106 | 84 | 95 | 64 | 69 | 79 |
| BD507 | | 28 | 38 | 30 | 25 | 6 | 4 | 85 | 77 | 93 | 55 | 83 | 86 | 80 | 92 | 80 | 85 | 80 | 87 | 89 |
| XG008 | | 35 | 36 | 30 | 22 | 52 | 28 | 45 | 33 | 62 | 61 | 131 | 86 | 81 | 89 | 87 | 87 | 87 | 97 | 93 |
| BD30-508 | | 36 | 46 | 46 | 24 | 5 | 4 | 78 | 80 | 73 | 45 | 80 | 106 | 78 | 108 | 99 | 92 | 126 | 81 | 90 |
| VA29-11 | | 54 | 57 | 46 | 48 | 62 | 67 | 57 | 57 | 28 | 17 | 88 | 80 | 89 | 104 | 114 | 101 | 95 | 81 | 96 |

| | | XGv013 | P4A1 | XGv026 | XG017 | H4 | 414 | XGv003 | XGv031 | P17 | XG014 | XGv016 | XGv042 | XG025 | FC08 | XGv030 | XG011 | A34-2 | XGv004 | FC05 |
|---|---|---|---|---|---|---|---|---|---|---|---|---|---|---|---|---|---|---|---|---|
| BD836 | | 3 | 3 | 3 | 3 | 12 | 3 | 4 | 3 | 6 | 4 | 56 | 67 | 72 | 67 | 63 | 51 | 71 | 32 | 93 |
| BD674 | | 5 | 6 | 15 | 5 | 11 | 4 | 23 | 16 | 42 | 5 | 70 | 37 | 49 | 50 | 72 | 45 | 33 | 46 | 78 |
| BD790 | | 31 | 24 | 26 | 25 | 12 | 3 | 75 | 57 | 76 | 20 | 46 | 52 | 95 | 85 | 82 | 79 | 50 | 82 | 100 |
| BD915 | | 19 | 36 | 28 | 28 | 10 | 5 | 67 | 51 | 68 | 43 | 73 | 55 | 43 | 66 | 76 | 58 | 82 | 51 | 82 |
| BD913 | | 49 | 48 | 57 | 63 | 12 | 3 | 86 | 70 | 86 | 26 | 42 | 60 | 10 | 53 | 76 | 73 | 69 | 78 | 93 |
| BD748 | | 45 | 63 | 56 | 42 | 10 | 4 | 53 | 68 | 96 | 12 | 54 | 25 | 5 | 17 | 46 | 45 | 50 | 37 | 81 |
| BD693 | | 39 | 56 | 49 | 44 | 12 | 4 | 54 | 62 | 99 | 25 | 81 | 36 | 55 | 64 | 70 | 53 | 59 | 51 | 80 |
| BD30-515 | | 7 | 15 | 11 | 8 | 6 | 3 | 41 | 44 | 63 | 35 | 77 | 41 | 64 | 103 | 73 | 86 | 99 | 81 | 93 |
| BD30-613 | | 27 | 30 | 30 | 27 | 5 | 4 | 82 | 89 | 80 | 43 | 78 | 72 | 66 | 118 | 91 | 100 | 106 | 84 | 97 |
| BD30-616 | | 9 | 18 | 14 | 9 | 6 | 3 | 45 | 52 | 64 | 24 | 94 | 87 | 75 | 108 | 67 | 82 | 100 | 80 | 93 |
| VA29-13 | | 20 | 22 | 17 | 11 | 31 | 14 | 31 | 33 | 46 | 18 | 27 | 27 | 73 | 103 | 89 | 104 | 99 | 85 | 105 |
| BD504 | | 16 | 22 | 16 | 12 | 6 | 3 | 71 | 54 | 72 | 31 | 100 | 89 | 78 | 91 | 84 | 93 | 85 | 91 | 105 |
| BD618 | | 6 | 12 | 11 | 8 | 9 | 3 | 45 | 33 | 85 | 31 | 85 | 91 | 78 | 103 | 94 | 93 | 93 | 94 | 97 |
| VA29-12 | | 55 | 57 | 48 | 63 | 32 | 61 | 57 | 54 | 70 | 64 | 53 | 54 | 81 | 113 | 94 | 91 | 78 | 78 | 101 |
| XG003 | | 95 | 86 | 92 | 78 | 46 | 23 | 69 | 81 | 108 | 39 | 74 | 47 | 107 | 95 | 77 | 91 | 83 | 101 | 83 |
| XG004 | | 74 | 74 | 91 | 83 | 21 | 28 | 117 | 52 | 46 | 9 | 99 | 64 | 143 | 140 | 42 | 67 | 55 | 80 | 94 |
| XG009 | | 49 | 48 | 39 | 36 | 18 | 36 | 57 | 31 | 44 | 60 | 71 | 59 | 119 | 102 | 81 | 94 | 91 | 94 | 96 |
| XG043 | | 44 | 47 | 48 | 41 | 15 | 50 | 47 | 40 | 42 | 44 | 50 | 43 | 74 | 87 | 72 | 76 | 97 | 82 | 99 |
| BD498 | | 54 | 51 | 54 | 37 | 6 | 5 | 127 | 89 | 97 | 67 | 80 | 88 | 71 | 119 | 95 | 87 | 86 | 88 | 87 |

| | | XGv013 | P4A1 | XGv026 | XG017 | H4 | 414 | XGv003 | XGv031 | P17 | XG014 | XGv016 | XGv042 | XG025 | FC08 | XGv030 | XG011 | A34-2 | XGv004 | FC05 |
|---|---|---|---|---|---|---|---|---|---|---|---|---|---|---|---|---|---|---|---|---|
| VA29-5 | | 11 | 28 | 11 | 24 | 40 | 27 | 7 | 8 | 10 | 11 | 11 | 11 | 72 | 105 | 97 | 79 | 76 | 81 | 91 |
| VA29-6 | | 16 | 14 | 11 | 28 | 52 | 41 | 7 | 7 | 12 | 19 | 16 | 13 | 75 | 92 | 95 | 87 | 85 | 80 | 87 |
| VA29-14 | | 13 | 12 | 11 | 25 | 51 | 42 | 5 | 7 | 9 | 16 | 14 | 10 | 75 | 90 | 94 | 103 | 97 | 102 | 98 |
| VA34-4 | | 16 | 15 | 12 | 38 | 55 | 44 | 6 | 5 | 26 | 33 | 106 | 90 | 119 | 114 | 104 | 101 | 91 | 80 | 92 |
| VA29-10-A | | 52 | 61 | 50 | 81 | 68 | 47 | 12 | 12 | 18 | 32 | 21 | 12 | 79 | 94 | 107 | 95 | 94 | 93 | 95 |
| BD897 | | 3 | 19 | 6 | 59 | 11 | 3 | 5 | 5 | 6 | 4 | 6 | 5 | 71 | 72 | 68 | 62 | 27 | 57 | 89 |
| BD826 | | 61 | 81 | 55 | 61 | 14 | 4 | 49 | 77 | 14 | 4 | 5 | 7 | 4 | 8 | 52 | 56 | 61 | 53 | 77 |
| BD914 | | 65 | 70 | 70 | 71 | 12 | 3 | 69 | 73 | 22 | 5 | 5 | 7 | 7 | 33 | 96 | 75 | 71 | 52 | 93 |
| BD901 | | 52 | 58 | 37 | 51 | 11 | 3 | 7 | 57 | 8 | 4 | 4 | 4 | 4 | 8 | 70 | 70 | 52 | 46 | 95 |
| BD692 | | 64 | 68 | 62 | 65 | 11 | 4 | 65 | 74 | 49 | 3 | 8 | 5 | 3 | 10 | 56 | 55 | 63 | 49 | 76 |
| BD941 | | 100 | 94 | 74 | 80 | 11 | 4 | 76 | 120 | 52 | 5 | 10 | 9 | 4 | 13 | 69 | 72 | 72 | 65 | 85 |
| VA13-8 | | 19 | 52 | 12 | 69 | 29 | 41 | 9 | 39 | 10 | 12 | 12 | 9 | 98 | 74 | 74 | 70 | 124 | 95 | 97 |
| XG036 | | 86 | 75 | 76 | 84 | 53 | 42 | 52 | 59 | 34 | 25 | 34 | 34 | 14 | 24 | 105 | 83 | 83 | 81 | 96 |
| XG031 | | 46 | 47 | 82 | 90 | 30 | 24 | 101 | 86 | 84 | 39 | 48 | 41 | 102 | 93 | 62 | 68 | 87 | 96 | 91 |
| XG005 | | 108 | 103 | 75 | 79 | 58 | 48 | 85 | 76 | 99 | 33 | 37 | 43 | 96 | 116 | 70 | 104 | 94 | 98 | 99 |
| XG013 | | 88 | 92 | 90 | 90 | 54 | 40 | 89 | 82 | 112 | 49 | 60 | 56 | 103 | 95 | 75 | 94 | 94 | 106 | 89 |
| XG016 | | 92 | 101 | 86 | 99 | 57 | 26 | 90 | 87 | 103 | 30 | 54 | 46 | 105 | 98 | 83 | 87 | 102 | 103 | 94 |
| VA12-11-B | | 102 | 93 | 101 | 84 | 109 | 98 | 101 | 94 | 102 | 92 | 98 | 116 | 100 | 80 | 86 | 98 | 110 | 112 | 106 |
| BD977 | | 93 | 100 | 73 | 98 | 76 | 90 | 81 | 79 | 161 | 41 | 75 | 62 | 78 | 74 | 7 | 12 | 5 | 7 | 84 |
| BD902 | | 44 | 66 | 101 | 70 | 54 | 72 | 66 | 89 | 80 | 87 | 63 | 96 | 77 | 66 | 68 | 39 | 22 | 29 | 89 |
| BD702 | | 56 | 82 | 55 | 84 | 67 | 75 | 76 | 99 | | | | | | | | | 21 | 22 | 89 |
| 553-15 | | 95 | 82 | 60 | 117 | 66 | 76 | 81 | 114 | 129 | 82 | 113 | 125 | 52 | 127 | 79 | 80 | 67 | 70 | 81 |
| VA11-1 | | 82 | 93 | 88 | 91 | 136 | 104 | 91 | 90 | 104 | 99 | 102 | 108 | 83 | 102 | 57 | 22 | 21 | 26 | 97 |
| XG026 | | 70 | 73 | 86 | 84 | 62 | 68 | 72 | 82 | 79 | 79 | 76 | 71 | 78 | 69 | 40 | 35 | 29 | 44 | 88 |

**Fig. S22 Data sheets of ELISA assay of NAbs neutralizing SARS-CoV-2 RBD.** Data of ELISA assay of different NAbs are listed in (A) and (B). NAbs corresponding to Class I-VI are filled with yellow, green, red, blue, brown and magenta colors, respectively. Values in the tables are filled with black (<25), grey (25~50), silver (50~70) and white (>70).

### Table S1. Information of recombinant antibodies

| Antibody ID | IGH V gene | SHM | Binding Affinity - $K_D$ (nM) | | | | |
|---|---|---|---|---|---|---|---|
| | | | WT | B.1.1.7 | P.1 | B.1.351 | B.1.617.2 |
| XGv001 | IGHV3-21 | 5.08% | 0.17 | 0.23 | 0.18 | 0.24 | 0.30 |
| XGv002 | IGHV3-33 | 6.76% | 4.15 | 4.42 | 7.18 | 5.80 | 3.91 |
| XGv003 | IGHV1-69 | 7.12% | 0.10 | 0.21 | NS | NS | 0.71 |
| XGv004 | IGHV1-18 | 3.41% | 0.14 | 0.10 | 0.11 | 0.16 | 0.20 |
| XGv005 | IGHV1-69-2 | 7.48% | 3.17 | 3.69 | 28.03 | 1.25 | 7.94 |
| XGv006 | IGHV1-18 | 8.53% | 3.68 | 3.06 | 9.33 | 6.65 | 3.86 |
| XGv007 | IGHV4-61 | 3.01% | 1.70 | 4.22 | 5.34 | 5.82 | 4.50 |
| XGv008 | IGHV4-39 | 2.68% | 0.47 | 0.45 | 0.30 | 0.34 | 0.28 |
| XGv009 | IGHV3-30 | 4.05% | 1.30 | 9.85 | 9.63 | NS | 22.60 |
| XGv010 | IGHV3-74 | 0.68% | 1.83 | 0.52 | NS | NS | 33.50 |
| XGv011 | IGHV3-30 | 6.94% | 0.10 | 0.50 | 0.64 | 3.93 | 0.66 |
| XGv012 | IGHV4-39 | 6.10% | 0.25 | 4.51 | 2.90 | 3.10 | 1.16 |
| XGv013 | IGHV3-53 | 3.75% | 0.26 | 1.22 | 5.58 | 9.74 | 0.29 |
| XGv014 | IGHV3-9 | 4.76% | 0.37 | 0.40 | 0.45 | 0.26 | 0.39 |
| XGv015 | IGHV3-30 | 4.73% | 21.01 | 19.93 | 20.39 | 31.41 | 21.67 |
| XGv016 | IGHV1-46 | 8.14% | 0.16 | 0.26 | 0.17 | 0.34 | 0.34 |
| XGv017 | IGHV3-53 | 5.12% | 0.31 | 0.31 | 1.21 | 8.43 | 0.26 |
| XGv018 | IGHV3-33 | 3.38% | 0.21 | 0.62 | 1.67 | 0.79 | 0.89 |
| XGv019 | IGHV3-13 | 6.85% | 0.59 | 0.24 | 0.84 | 0.43 | 0.48 |
| XGv020 | IGHV3-30 | 4.41% | 0.29 | 0.67 | 0.19 | 0.24 | 0.17 |
| XGv021 | IGHV3-30 | 5.41% | 0.26 | 0.35 | 0.40 | 0.18 | 0.41 |
| XGv022 | IGHV3-30 | 4.78% | 25.60 | 27.40 | 13.70 | 15.80 | 20.70 |
| XGv023 | IGHV3-13 | 5.48% | 0.21 | 0.62 | 1.67 | 0.79 | 0.89 |
| XGv024 | IGHV3-13 | 2.05% | 0.76 | 0.51 | 1.08 | 0.67 | 0.58 |
| XGv025 | IGHV3-30-3 | 4.05% | 1.22 | 2.20 | 3.74 | 4.46 | 3.02 |
| XGv026 | IGHV3-53 | 6.83% | 0.23 | 0.33 | 0.69 | 0.81 | 0.15 |
| XGv027 | IGHV4-34 | 7.67% | 4.16 | 7.78 | 6.20 | 13.73 | 6.64 |
| XGv028 | IGHV2-5 | 5.03% | 0.68 | 1.11 | NS | 1.78 | NS |
| XGv029 | IGHV7-4-1 | 2.70% | 14.00 | 13.60 | 16.10 | 13.50 | 17.30 |
| XGv030 | IGHV3-48 | 3.73% | 1.30 | 1.83 | 2.12 | 1.85 | 1.68 |
| XGv031 | IGHV1-69 | 2.72% | 0.26 | 0.16 | 0.50 | 0.47 | 0.48 |
| XGv032 | IGHV3-13 | 5.14% | 0.16 | 0.16 | 0.11 | 0.09 | 0.09 |
| XGv033 | IGHV4-4 | 5.48% | 1.86 | 2.43 | 4.10 | 4.40 | 3.24 |
| XGv034 | IGHV3-48 | 5.74% | 4.13 | 7.19 | 9.17 | 11.63 | 6.42 |
| XGv035 | IGHV7-4-1 | 3.38% | 3.98 | 3.76 | 5.01 | 3.45 | 5.22 |
| XGv036 | IGHV1-18 | 8.50% | 0.63 | 0.99 | 1.82 | 1.51 | 1.69 |
| XGv038 | IGHV3-33 | 3.04% | 0.43 | 0.59 | 0.33 | 0.28 | 0.29 |
| XGv039 | IGHV3-13 | 5.82% | 1.00 | 0.75 | 1.36 | 1.54 | 0.66 |
| XGv040 | IGHV3-53 | 2.73% | 0.91 | 0.68 | 11.56 | 18.03 | 1.10 |
| XGv041 | IGHV3-30-3 | 4.08% | NS | NS | NS | NS | NS |
| XGv042 | IGHV2-5 | 4.71% | 0.16 | 0.17 | 0.29 | 0.16 | 0.27 |
| XGv043 | IGHV4-39 | 7.59% | 9.79 | NS | 6.39 | NS | NS |
| XGv044 | IGHV3-23 | 6.10% | 2.17 | 13.93 | 17.71 | 23.15 | 22.84 |
| XGv045 | IGHV4-61 | 5.74% | 0.40 | 0.94 | 1.29 | 1.41 | 0.92 |
| XGv046 | IGHV4-34 | 9.31% | 0.53 | 1.10 | 0.95 | 2.03 | 1.24 |
| XGv047 | IGHV1-69 | 5.86% | 0.10 | 0.08 | 0.17 | 0.32 | 0.79 |
| XGv049 | IGHV3-13 | 6.48% | 0.39 | 0.17 | 0.58 | 0.40 | 0.18 |
| XGv050 | IGHV3-13 | 4.79% | 0.26 | 0.19 | 0.37 | 0.60 | 2.87 |
| XG011 | IGHV3-13 | 4.11% | 2.74 | 1.74 | 3.40 | 2.50 | 2.19 |
| XG014 | IGHV5-51 | 2.36% | 1.78 | 1.13 | 1.90 | 1.30 | 1.14 |
| XG017 | IGHV3-66 | 2.40% | 1.52 | 1.28 | NS | NS | 0.75 |
| XG025 | IGHV3-33 | 3.05% | 1.60 | 0.77 | 1.36 | 1.41 | 0.64 |
| H014 | N/A | N/A | 0.11 | 0.25 | 0.21 | 0.26 | 0.14 |
| HB27 | N/A | N/A | 0.99 | 1.88 | 1.32 | 1.57 | 1.37 |
| FC08 | IGHV1-69 | 0.33% | 0.10 | 0.25 | 0.46 | 0.49 | 1.32 |
| P17 | N/A | N/A | 3.97 | 9.66 | NS | NS | 25.00 |
| 414 | IGHV1-46 | 0.00% | 0.92 | NS | NS | NS | 5.99 |
| A8-1 | IGHV3-53 | 1.71% | 0.51 | 0.86 | 2.45 | 12.62 | 0.34 |
| S309 | IGHV1-18 | 2.78% | 0.75 | 0.55 | 1.01 | 0.93 | 0.60 |
| REGN10987 | IGHV3-30 | 1.37% | 1.91 | 3.14 | 2.84 | 3.43 | 4.16 |
| 553 | IGHV3-7 | 3.12% | 3.06 | 9.07 | 7.49 | 8.93 | 10.48 |
| 1-57 | IGHV3-72 | 0.70% | 5.86 | 16.22 | 23.82 | 24.04 | NS |
| A34-2 | IGHV3-9 | 3.09% | 0.36 | 0.31 | 2.08 | 1.45 | 0.41 |
| A5-10 | IGHV3-23 | 2.04% | 1.78 | 1.96 | 1.43 | 1.99 | 0.33 |
| B38 | IGHV3-53 | 1.00% | 21.89 | 14.03 | NS | NS | 25.80 |
| BD-368-2 | IGHV3-23 | 7.98% | 2.62 | 6.67 | NS | NS | 31.60 |
| C121 | IGHV1-2 | 1.02% | 1.49 | 4.39 | NS | NS | 1.98 |
| C135 | IGHV3-30 | 2.05% | 5.96 | 15.52 | 17.09 | 21.20 | 8.72 |
| C144 | IGHV3-53 | 1.37% | 21.87 | 62.99 | NS | NS | 51.93 |
| CB6 | IGHV3-66 | 1.00% | 17.25 | 51.64 | NS | NS | 46.18 |
| CC12.3 | IGHV3-53 | N/A | 7.70 | 21.26 | NS | NS | 12.14 |
| COVA1-16 | IGHV1-46 | 0.34% | 1.02 | 0.95 | 4.01 | 2.10 | 8.71 |
| COVA2-04 | IGHV4-4 | 1.10% | 33.13 | 30.49 | NS | NS | 29.48 |

| | | | | | | | |
|---|---|---|---|---|---|---|---|
| COVA2-39 | IGHV3-53 | 1.10% | 2.69 | 7.62 | NS | NS | 3.62 |
| CV07-270 | IGHV3-11 | 0.68% | 22.87 | 40.06 | NS | NS | NS |
| DH1047 | IGHV1-46 | N/A | 0.21 | 0.60 | 0.80 | 0.73 | 0.49 |
| CC12.1 | IGHV3-53 | N/A | 8.85 | - | - | - | 9.76 |
| COVOX-150 | IGHV3-53 | 3.16% | 1.40 | - | - | - | 1.79 |
| COVOX-222 | IGHV3-53 | 2.46% | 1.22 | - | - | - | 1.48 |
| COVOX-269 | IGHV3-53 | 4.21% | 3.88 | - | - | - | 4.64 |
| COVOX-40 | IGHV3-53 | 1.75% | 0.89 | - | - | - | 1.12 |
| P2C-1F11 | IGHV3-11 | 1.75% | 9.75 | - | - | - | 10.87 |
| BD-236 | IGHV3-53 | 3.10% | 15.80 | - | - | - | - |
| BD-604 | IGHV3-53 | 4.00% | 0.31 | - | - | - | - |
| BD-623 | IGHV3-66 | 2.75% | 1.30 | - | - | - | - |
| BD-629 | IGHV3-53 | 1.70% | 0.93 | - | - | - | - |
| BD23 | IGHV7-4-1 | 0.00% | 34.99 | - | - | - | - |
| BD30-494 | IGHV3-53 | 1.02% | 7.04 | - | - | - | - |
| BD30-498 | IGHV3-53 | 1.71% | 16.37 | - | - | - | - |
| BD30-503 | IGHV3-53 | 2.39% | 1.81 | - | - | - | - |
| BD30-504 | IGHV1-NL1 | 7.67% | 4.32 | - | - | - | - |
| BD30-505 | IGHV3-53 | N/A | 4.01 | - | - | - | - |
| BD30-507 | IGHV3-53 | 1.71% | 2.82 | - | - | - | - |
| BD30-508 | IGHV3-53 | 1.71% | 10.65 | - | - | - | - |
| BD30-515 | IGHV3-66 | 2.05% | 1.36 | - | - | - | - |
| BD30-605 | IGHV3-53 | 0.40% | 15.12 | - | - | - | - |
| BD30-613 | IGHV3-66 | 0.00% | 6.88 | - | - | - | - |
| BD30-616 | IGHV3-66 | 1.03% | 3.48 | - | - | - | - |
| BD30-618 | IGHV3-66 | 4.79% | 1.00 | - | - | - | - |
| BD667 | IGHV3-11 | 7.12% | 2.56 | - | - | - | - |
| BD674 | IGHV4-4 | 4.39% | 0.34 | - | - | - | - |
| BD692 | IGHV3-48 | 1.01% | 0.29 | - | - | - | - |
| BD693 | IGHV3-15 | 2.68% | 0.22 | - | - | - | - |
| BD702 | IGHV3-48 | 2.38% | 0.85 | - | - | - | - |
| BD744 | IGHV3-9 | 4.05% | 0.21 | - | - | - | - |
| BD748 | IGHV3-48 | 4.41% | 0.73 | - | - | - | - |
| BD771 | IGHV3-15 | 4.65% | 0.21 | - | - | - | - |
| BD790 | IGHV1-2 | 4.75% | 0.75 | - | - | - | - |
| BD804 | IGHV3-21 | 3.04% | 0.37 | - | - | - | - |
| BD812 | IGHV5-51 | 5.90% | 0.32 | - | - | - | - |
| BD813 | IGHV3-53 | 3.44% | 0.42 | - | - | - | - |
| BD826 | IGHV4-4 | 7.46% | 0.04 | - | - | - | - |
| BD836 | IGHV1-58 | 6.14% | 0.09 | - | - | - | - |
| BD837 | IGHV1-58 | 5.80% | 0.07 | - | - | - | - |
| BD868 | IGHV3-53 | 7.85% | 0.24 | - | - | - | - |
| BD870 | IGHV3-48 | 9.15% | 0.52 | - | - | - | - |
| BD897 | IGHV1-69 | 9.15% | 0.11 | - | - | - | - |
| BD901 | IGHV1-69 | 10.14% | 0.12 | - | - | - | - |
| BD902 | IGHV3-33 | 5.07% | 0.74 | - | - | - | - |
| BD906 | IGHV4-39 | 9.70% | 0.25 | - | - | - | - |
| BD907 | IGHV7-4-1 | 4.05% | 0.39 | - | - | - | - |
| BD912 | IGHV3-21 | 6.25% | 0.50 | - | - | - | - |
| BD913 | IGHV1-69 | 1.69% | 0.71 | - | - | - | - |
| BD914 | IGHV1-69 | 4.05% | 0.14 | - | - | - | - |
| BD915 | IGHV4-4 | 2.41% | 0.53 | - | - | - | - |
| BD930 | IGHV4-39 | 3.68% | 0.17 | - | - | - | - |
| BD941 | IGHV4-31 | 4.70% | 0.28 | - | - | - | - |
| BD977 | IGHV3-30 | 4.73% | 0.13 | - | - | - | - |
| BG1-22 | IGHV3-66 | N/A | 21.79 | - | - | - | - |
| C002 | IGHV3-30 | 0.34% | 19.66 | - | - | - | - |
| C102 | IGHV3-53 | 0.68% | 14.27 | - | - | - | - |
| C105 | IGHV3-53 | 0.34% | 23.72 | - | - | - | - |
| C110 | IGHV5-51 | 0.34% | 0.90 | - | - | - | - |
| C119 | IGHV1-46 | 0.34% | 5.04 | - | - | - | - |
| C1A-B12 | IGHV3-53 | 1.40% | 3.89 | - | - | - | - |
| C1A-B3 | IGHV3-53 | 2.11% | 16.96 | - | - | - | - |
| C1A-C2 | IGHV3-53 | 1.40% | 15.39 | - | - | - | - |
| C1A-F10 | IGHV3-53 | 2.11% | 17.69 | - | - | - | - |
| COVOX-45 | IGHV1-33 | 1.39% | 5.25 | - | - | - | - |
| COVOX-75 | IGHV3-30 | 4.86% | 17.06 | - | - | - | - |
| CV30 | IGHV3-53 | N/A | 11.31 | - | - | - | - |
| H4 | IGHV1-2 | 0.00% | 8.56 | - | - | - | - |
| LY-CoV481 | IGHV3-53 | N/A | 7.76 | - | - | - | - |
| LY-CoV488 | IGHV3-53 | N/A | 35.83 | - | - | - | - |
| P2B-1A10 | IGHV3-53 | 0.35% | 29.84 | - | - | - | - |
| P4A1 | IGHV3-53 | N/A | 13.58 | - | - | - | - |
| P5A-1B8 | IGHV3-53 | 1.40% | 10.57 | - | - | - | - |
| P5A-2G9 | IGHV3-33 | 0.00% | 7.52 | - | - | - | - |
| P5A-3A1 | IGHV3-53 | 0.00% | 11.28 | - | - | - | - |
| REGN10933 | IGHV3-11 | 1.35% | 1.39 | - | - | - | - |

| | | | | | | | |
|---|---|---|---|---|---|---|---|
| S2A4 | IGHV3-7 | N/A | 5.60 | - | - | - | - |
| S304 | IGHV3-13 | 2.11% | 1.99 | - | - | - | - |
| VA11-1_H | IGHV3-30 | 1.36% | 0.52 | - | - | - | - |
| VA13-6_H | IGHV3-66 | 0.34% | 0.82 | - | - | - | - |
| VA13-7_H | IGHV3-66 | 0.34% | 0.98 | - | - | - | - |
| VA13-8_H | IGHV1-69 | 2.71% | 0.21 | - | - | - | - |
| VA13-9_H | IGHV3-66 | 0.34% | 1.14 | - | - | - | - |
| VA14-3_H | IGHV3-66 | 1.37% | 1.28 | - | - | - | - |
| VA14-5_H | IGHV3-53 | 3.41% | 1.10 | - | - | - | - |
| VA29-10-A_H | IGHV1-69 | 3.07% | 0.42 | - | - | - | - |
| VA29-11_H | IGHV3-53 | 1.73% | 1.75 | - | - | - | - |
| VA29-12_H | IGHV1-69 | 3.05% | 4.94 | - | - | - | - |
| VA29-13_H | IGHV2-5 | 1.00% | 2.80 | - | - | - | - |
| VA29-14_H | IGHV1-69 | 5.14% | 0.40 | - | - | - | - |
| VA29-18_H | IGHV3-53 | 2.73% | 0.99 | - | - | - | - |
| VA29-5_H | IGHV1-69 | 3.04% | 4.22 | - | - | - | - |
| VA29-6_H | IGHV1-69 | 3.08% | 0.35 | - | - | - | - |
| VA32-1_H | IGHV3-66 | 4.78% | 1.28 | - | - | - | - |
| VA32-5_H | IGHV1-69 | 3.05% | 1.02 | - | - | - | - |
| VA34-4_H | IGHV3-53 | 2.08% | 0.32 | - | - | - | - |
| VA34-6_H | IGHV4-34 | 3.41% | 1.04 | - | - | - | - |
| VA34-7_H | IGHV4-34 | 3.41% | 0.97 | - | - | - | - |
| VA5-1_H | IGHV3-66 | 1.71% | 3.87 | - | - | - | - |
| VA5-2_H | IGHV3-66 | 1.71% | 0.58 | - | - | - | - |
| VA5-8_H | IGHV3-53 | 3.07% | 5.49 | - | - | - | - |
| VA5-9_H | IGHV3-66 | 1.37% | 1.15 | - | - | - | - |
| VA8-3_H | IGHV3-66 | 4.10% | 0.48 | - | - | - | - |
| VA8-4_H | IGHV3-49 | 3.32% | 4.30 | - | - | - | - |
| VA8-5_H | IGHV3-49 | 4.32% | 3.68 | - | - | - | - |
| VA9-3_H | IGHV3-53 | 1.71% | 1.21 | - | - | - | - |
| VA9-5_H | IGHV3-53 | 3.07% | 2.33 | - | - | - | - |
| XG003 | IGHV2-70 | 5.69% | 2.29 | - | - | - | - |
| XG004 | IGHV2-70 | 0.00% | 1.19 | - | - | - | - |
| XG005 | IGHV2-5 | 1.69% | 0.67 | - | - | - | - |
| XG008 | IGHV1-69 | 2.36% | 0.41 | - | - | - | - |
| XG009 | IGHV1-8 | 1.01% | 25.28 | - | - | - | - |
| XG013 | IGHV2-70 | 2.68% | 0.50 | - | - | - | - |
| XG016 | IGHV2-5 | 4.38% | 2.35 | - | - | - | - |
| XG031 | IGHV2-5 | 3.68% | 1.24 | - | - | - | - |
| XG036 | IGHV1-69 | 5.78% | 1.71 | - | - | - | - |
| XG038 | IGHV1-18 | 4.39% | 1.42 | - | - | - | - |
| XG043 | IGHV3-20 | 0.34% | 7.39 | - | - | - | - |

### Table S2. Neutralizing titers of recombinant antibodies

| Antibody ID | Neutralizing Titer - $IC_{50}$ (ng/mL) | | | | | | | | | |
|---|---|---|---|---|---|---|---|---|---|---|
| | Pseudovirus | | | | | | Authentic virus | | | |
| | WT | B.1.1.7 | P.1 | B.1.351 | B.1.617.2 | SARS-CoV | WT | P.1 | B.1.351 | B.1.617.2 |
| XGv001 | 736 | 519 | 458 | 792 | 815 | 115 | 16667 | 16667 | 16667 | 12500 |
| XGv002 | 1201 | >2000 | >2000 | >2000 | >2000 | >2000 | 8333 | >50000 | >50000 | >50000 |
| XGv003 | 5 | 7 | >2000 | >2000 | 20 | >2000 | 24 | >50000 | >50000 | 33 |
| XGv004 | 118 | 131 | 30 | 72 | 144 | >2000 | 391 | 1042 | 1563 | 1042 |
| XGv005 | >2000 | >2000 | >2000 | >2000 | >2000 | >2000 | >50000 | >50000 | >50000 | >50000 |
| XGv006 | 1563 | >2000 | >2000 | >2000 | >2000 | >2000 | 2083 | >50000 | >50000 | >50000 |
| XGv007 | >2000 | >2000 | >2000 | >2000 | >2000 | >2000 | 50000 | >50000 | >50000 | >50000 |
| XGv008 | 1073 | 955 | 1436 | 1769 | 2000 | >2000 | 1042 | >50000 | >50000 | >50000 |
| XGv009 | >2000 | >2000 | >2000 | >2000 | >2000 | >2000 | 50000 | >50000 | >50000 | >50000 |
| XGv010 | 6 | 6 | >2000 | >2000 | >2000 | >2000 | 24 | >50000 | >50000 | >50000 |
| XGv011 | 25 | 37 | 56 | 178 | 9 | >2000 | 1042 | 2083 | 4167 | 391 |
| XGv012 | 2000 | >2000 | >2000 | >2000 | >2000 | >2000 | 33333 | >50000 | >50000 | >50000 |
| XGv013 | 10 | 10 | 2 | 5 | 12 | >2000 | 65 | 130 | 195 | 130 |
| XGv014 | 1199 | 745 | 414 | 1021 | 1022 | 486 | 33333 | 50000 | 33333 | 8333 |
| XGv015 | 1406 | 1421 | 291 | 590 | 992 | >2000 | 12500 | 6250 | 33333 | 1563 |
| XGv016 | 1 | 1 | 1 | 2 | 4 | >2000 | 24 | 24 | 24 | 24 |
| XGv017 | 16 | 13 | 4 | 6 | 16 | >2000 | 98 | 130 | 130 | 195 |
| XGv018 | 839 | 618 | 389 | 449 | 756 | 73 | 4167 | 12500 | 12500 | 8333 |
| XGv019 | 847 | 893 | 801 | 747 | 1104 | >2000 | 12500 | 12500 | 16667 | 16667 |
| XGv020 | 723 | 1132 | 1365 | 1531 | 1459 | >2000 | 6250 | 33333 | 33333 | 33333 |
| XGv021 | >2000 | >2000 | >2000 | >2000 | >2000 | >2000 | >50000 | >50000 | >50000 | >50000 |
| XGv022 | >2000 | >2000 | >2000 | >2000 | >2000 | >2000 | >50000 | >50000 | >50000 | >50000 |
| XGv023 | 1126 | 961 | 959 | 614 | 957 | 741 | 4167 | 16667 | 12500 | 16667 |
| XGv024 | 603 | 702 | 955 | 1028 | 1459 | >2000 | 4167 | 16667 | 8333 | 16667 |
| XGv025 | 2000 | >2000 | >2000 | >2000 | >2000 | >2000 | 12500 | >50000 | >50000 | >50000 |
| XGv026 | 10 | 13 | 3 | 5 | 9 | >2000 | 33 | 24 | 24 | 98 |
| XGv027 | 1259 | 2083 | 2083 | 2083 | 2083 | >2000 | 49 | >50000 | >50000 | 1563 |
| XGv028 | 2000 | >2000 | >2000 | >2000 | >2000 | >2000 | 16667 | >50000 | >50000 | 50000 |
| XGv029 | 2000 | >2000 | >2000 | >2000 | >2000 | >2000 | 33333 | >50000 | >50000 | >50000 |
| XGv030 | 850 | 809 | 865 | 1125 | 1101 | 41 | 24 | 24 | 33 | 33333 |
| XGv031 | 4 | 5 | 1 | 2 | 11 | >2000 | 24 | 24 | 24 | 24 |
| XGv032 | 194 | 350 | 243 | 348 | 439 | >2000 | 1563 | 4167 | 4167 | 2083 |
| XGv033 | >2000 | >2000 | >2000 | >2000 | >2000 | >2000 | >50000 | >50000 | >50000 | >50000 |
| XGv034 | >2000 | >2000 | >2000 | >2000 | >2000 | >2000 | >50000 | >50000 | >50000 | >50000 |
| XGv035 | 2000 | >2000 | >2000 | >2000 | >2000 | >2000 | 6250 | >50000 | >50000 | >50000 |
| XGv036 | 2000 | >2000 | >2000 | >2000 | >2000 | >2000 | 12500 | >50000 | >50000 | >50000 |
| XGv038 | 283 | 220 | 334 | 764 | 833 | >2000 | 3125 | 8333 | 12500 | >50000 |
| XGv039 | 1165 | 1695 | 690 | 958 | 1317 | 1250 | 195 | 391 | 2083 | 12500 |
| XGv040 | 21 | 40 | 6 | 15 | 16 | >2000 | 130 | 391 | 391 | 521 |
| XGv041 | >2000 | >2000 | >2000 | >2000 | >2000 | >2000 | >50000 | >50000 | >50000 | >50000 |
| XGv042 | 2 | 2 | 1 | 2 | 5 | >2000 | 24 | 24 | 24 | 33 |
| XGv043 | 181 | >2000 | >2000 | 2083 | >2000 | >2000 | 781 | >50000 | >50000 | >50000 |
| XGv044 | >2000 | >2000 | >2000 | >2000 | >2000 | >2000 | >50000 | >50000 | >50000 | >50000 |
| XGv045 | 2000 | >2000 | >2000 | >2000 | >2000 | >2000 | 25000 | >50000 | >50000 | >50000 |
| XGv046 | 586 | 1447 | 1158 | 1159 | 2000 | >2000 | 8333 | 8333 | 6250 | >50000 |
| XGv047 | 20 | 12 | 4 | 9 | 56 | >2000 | 6250 | 16667 | 16667 | 781 |
| XGv049 | 911 | 937 | 578 | 634 | 1168 | 552 | 65 | 195 | 195 | 12500 |
| XGv050 | 5 | 5 | 6 | 3 | 3 | >2000 | 391 | 2083 | 1042 | 98 |
| XG011 | 1832 | 1735 | 1236 | 1145 | 1614 | | - | - | - | - |
| XG014 | 2 | 2 | 2 | 3 | 3 | | - | - | - | - |
| XG017 | 9 | 10 | 1966 | 1248 | 16 | | - | - | - | - |
| XG025 | 163 | 442 | 255 | 640 | 820 | | - | - | - | - |
| H014 | 86 | 212 | 152 | 246 | 143 | | - | - | - | - |
| HB27 | 1 | 309 | 48 | 130 | 1 | | - | - | - | - |
| FC08 | 10 | 5 | 6 | 28 | 1207 | | - | - | - | - |
| P17 | 3 | 6 | >2000 | >2000 | 13 | | - | - | - | - |
| 414 | 18 | >2000 | >2000 | >2000 | 1124 | | - | - | - | - |
| A8-1 | 6 | 4 | 4 | 7 | 5 | | - | - | - | - |
| S309 | 30 | 23 | 12 | 7 | 12 | | - | - | - | - |
| REGN10987 | 2 | 1 | 1 | 1 | 3 | | - | - | - | - |

**Table S3. Data collection, processing and refinement**

| Protein | Spike in complex with XGv013 | Spike in complex with XGv043 | Spike in complex with XGv004, XGv016 and XGv030 | Spike in complex with XGv026 and XGv046 | Spike in complex with XGv018, XGv038 and XGv042 |
|---|---|---|---|---|---|
| Voltage (kV) | 300 | 300 | 300 | 300 | 300 |
| Detector | K2 | K2 | K2 | K2 | K2 |
| Pixel size (Å) | 1.04 | 1.04 | 1.04 | 1.04 | 1.04 |
| Electron dose ($e^-$/$Å^2$) | 60 | 60 | 60 | 60 | 60 |
| Defocus range (μm) | 1.5-2.7 | 1.5-2.7 | 1.5-2.7 | 1.5-2.7 | 1.5-2.7 |
| Final particles | 21,329 | 13,548 | 20,738 | 9,617 | 13,637 |
| Final resolution (Å) | 5.2 | 6.5 | 7.0 | 7.8 | 6.7 |
